## Supplementary Material for "Cohort study of cardiovascular safety of different COVID-19 vaccination doses among 46 million English adults"

### Contents – Supplementary Tables

### Supplementary Tables

#### Supplementary Table 1: Characterization of the population under study – dose 2 cohort[1]

| | | Those receiving dose 1 and eligible for dose 2 analysis | Vaccination status during dose 2 follow-up | | | |
|---|---|---|---|---|---|---|
| | | | Remains unvaccinated | ChAdOx1 | BNT-162b2 | mRNA-1273 |
| **N (%)** | All | 37251370 | 1540465 (4.14%) | 18868545 (50.7%) | 15876210 (42.6%) | 966150 (2.59%) |
| **Sex (%)** | Male | 17889600 (48.0%) | 790760 (51.3%) | 9204605 (48.8%) | 7368365 (46.4%) | 525870 (54.4%) |
| | Female | 19361775 (52.0%) | 749710 (48.7%) | 9663940 (51.2%) | 8507845 (53.6%) | 440280 (45.6%) |
| **Age in years (%)** | 18-29 | 5550870 (14.9%) | 519295 (33.7%) | 875685 (4.64%) | 3776100 (23.8%) | 379790 (39.3%) |
| | 30-39 | 5999670 (16.1%) | 405280 (26.3%) | 1413835 (7.49%) | 3821875 (24.1%) | 358680 (37.1%) |
| | 40-49 | 6070765 (16.3%) | 248650 (16.1%) | 4011380 (21.3%) | 1611990 (10.2%) | 198745 (20.6%) |
| | 50-59 | 6875320 (18.5%) | 153380 (9.96%) | 5221950 (27.7%) | 1476100 (9.30%) | 23890 (2.47%) |
| | 60-69 | 5531095 (14.8%) | 77465 (5.03%) | 3841270 (20.4%) | 1608570 (10.1%) | 3790 (0.39%) |
| | 70-79 | 4518565 (12.1%) | 53345 (3.46%) | 2657470 (14.1%) | 1806765 (11.4%) | 985 (0.10%) |
| | 80-89 | 2197495 (5.90%) | 52050 (3.38%) | 640635 (3.40%) | 1504580 (9.48%) | 230 (0.02%) |
| | 90+ | 507585 (1.36%) | 30990 (2.01%) | 206320 (1.09%) | 270235 (1.70%) | 40 (0.00%) |
| **Ethnicity (%)** | Asian | 2884950 (7.74%) | 184470 (12.0%) | 1274095 (6.75%) | 1359280 (8.56%) | 67105 (6.95%) |
| | Black | 1023165 (2.75%) | 104660 (6.79%) | 465195 (2.47%) | 427080 (2.69%) | 26230 (2.71%) |
| | Mixed | 468275 (1.26%) | 35795 (2.32%) | 188815 (1.00%) | 224100 (1.41%) | 19565 (2.03%) |
| | Other | 821445 (2.21%) | 48970 (3.18%) | 351410 (1.86%) | 388005 (2.44%) | 33060 (3.42%) |
| | White | 31195550 (83.7%) | 1130285 (73.4%) | 16168500 (85.7%) | 13111455 (82.6%) | 785310 (81.3%) |
| | Unknown | 618965 (1.66%) | 25025 (1.62%) | 308525 (1.64%) | 260965 (1.64%) | 24450 (2.53%) |
| | Missing | 239020 (0.64%) | 11265 (0.73%) | 112000 (0.59%) | 105325 (0.66%) | 10430 (1.08%) |
| **Index of multiple deprivation (%) (lowest: most deprived; highest: last deprived).** | 1-2 | 6449620 (17.3%) | 467955 (30.4%) | 3143120 (16.7%) | 2686540 (16.9%) | 152005 (15.7%) |
| | 3-4 | 7051385 (18.9%) | 355100 (23.1%) | 3470440 (18.4%) | 3037825 (19.1%) | 188020 (19.5%) |
| | 5-6 | 7396310 (19.9%) | 271710 (17.6%) | 3764675 (19.9%) | 3171410 (20.0%) | 188515 (19.5%) |
| | 7-8 | 7509945 (20.2%) | 216265 (14.0%) | 3924635 (20.8%) | 3183135 (20.1%) | 185910 (19.2%) |
| | 9-10 | 7498525 (20.1%) | 166605 (10.8%) | 4024335 (21.3%) | 3114460 (19.6%) | 193125 (20.0%) |
| | Missing | 1345585 (3.61%) | 62830 (4.08%) | 541340 (2.87%) | 682840 (4.30%) | 58575 (6.06%) |
| **Smoking status (%)** | Current smoker | 5538470 (14.9%) | 448330 (29.1%) | 2833035 (15.0%) | 2104945 (13.3%) | 152160 (15.8%) |
| | Former smoker | 9241950 (24.8%) | 266870 (17.3%) | 5189935 (27.5%) | 3633300 (22.9%) | 151845 (15.7%) |
| | Never smoker | 21263525 (57.1%) | 712255 (46.2%) | 10586335 (56.1%) | 9385040 (59.1%) | 579895 (60.0%) |
| | Missing | 1207430 (3.24%) | 113010 (7.34%) | 259240 (1.37%) | 752930 (4.74%) | 82250 (8.51%) |
| **Medical history (%)** | AMI[2] | 1094110 (2.94%) | 34585 (2.25%) | 558100 (2.96%) | 500685 (3.15%) | 740 (0.08%) |
| | Diabetes | 3693320 (9.91%) | 102485 (6.65%) | 1989370 (10.5%) | 1593155 (10.0%) | 8310 (0.86%) |
| | Depression | 8243490 (22.1%) | 418265 (27.1%) | 4486845 (23.8%) | 3185615 (20.1%) | 152765 (15.8%) |
| | Obesity | 4100690 (11.0%) | 145970 (9.48%) | 2287020 (12.1%) | 1626170 (10.2%) | 41530 (4.30%) |
| | Cancer | 6574195 (17.6%) | 218480 (14.2%) | 3345820 (17.7%) | 2884915 (18.2%) | 124980 (12.9%) |
| | COPD[3] | 1450725 (3.89%) | 52580 (3.41%) | 776770 (4.12%) | 619830 (3.90%) | 1545 (0.16%) |
| | Liver disease | 199480 (0.54%) | 11895 (0.77%) | 114345 (0.61%) | 72665 (0.46%) | 575 (0.06%) |
| | Chronic kidney disease | 2706535 (7.27%) | 94820 (6.16%) | 1324560 (7.02%) | 1282125 (8.08%) | 5030 (0.52%) |
| | Dementia | 476765 (1.28%) | 34425 (2.23%) | 255770 (1.36%) | 186405 (1.17%) | 165 (0.02%) |
| | All stroke | 783585 (2.10%) | 29400 (1.91%) | 414900 (2.20%) | 338485 (2.13%) | 800 (0.08%) |
| | All VTE[4] | 595795 (1.60%) | 28100 (1.82%) | 325375 (1.72%) | 240650 (1.52%) | 1670 (0.17%) |
| | Thrombophilia | 39750 (0.11%) | 2160 (0.14%) | 20155 (0.11%) | 16950 (0.11%) | 485 (0.05%) |
| **Major surgery in the last year (%)** | | 3438335 (9.23%) | 3438335 (9.23%) | 181420 (11.8%) | 1635875 (8.67%) | 1564930 (9.86%) |
| **Number of unique diseases in the last year (%)** | 0 | 29317000 (78.7%) | 1212325 (78.7%) | 14790460 (78.4%) | 12453645 (78.4%) | 860570 (89.1%) |
| | 1-5 | 7813905 (21.0%) | 319400 (20.7%) | 4012770 (21.3%) | 3376330 (21.3%) | 105405 (10.9%) |
| | 6+ | 120470 (0.32%) | 8750 (0.57%) | 65310 (0.35%) | 46235 (0.29%) | 175 (0.02%) |
| **Region (%)** | Northwest | 4707675 (12.6%) | 229505 (14.9%) | 2371585 (12.6%) | 1995750 (12.6%) | 110835 (11.5%) |
| | Southeast | 5674780 (15.2%) | 191450 (12.4%) | 2979705 (15.8%) | 2322200 (14.6%) | 181425 (18.8%) |
| | London | 5090990 (13.7%) | 262050 (17.0%) | 2312295 (12.2%) | 2354265 (14.8%) | 162380 (16.8%) |
| | East of England | 3439430 (9.23%) | 127030 (8.25%) | 1810815 (9.60%) | 1416010 (8.92%) | 85575 (8.86%) |
| | Southwest | 3167570 (8.50%) | 102225 (6.64%) | 1605125 (8.51%) | 1368590 (8.62%) | 91630 (9.48%) |
| | Yorkshire and the Humber | 3781095 (10.2%) | 142590 (9.26%) | 1841720 (9.76%) | 1678690 (10.6%) | 118095 (12.2%) |
| | East Midlands | 3577685 (9.60%) | 154570 (10.0%) | 1872735 (9.93%) | 1444470 (9.10%) | 105910 (11.0%) |
| | West Midlands | 2679810 (7.19%) | 96145 (6.24%) | 1420480 (7.53%) | 1152785 (7.26%) | 10400 (1.08%) |
| | Northeast | 3688215 (9.90%) | 171180 (11.1%) | 1906780 (10.1%) | 1544495 (9.73%) | 65760 (6.81%) |
| | Missing | 1444125 (3.88%) | 63720 (4.14%) | 747310 (3.96%) | 598955 (3.77%) | 34140 (3.53%) |
| **Prior COVID-19 at index date** | | 2496910 (6.7%) | 2496910 (6.70%) | 204985 (13.3%) | 1138400 (6.03%) | 1073310 (6.76%) |
| **Medications taken in the last 3 months** | Antiplatelet | 2342070 (6.29%) | 55160 (3.58%) | 1193905 (6.33%) | 1090955 (6.87%) | 2050 (0.21%) |
| | Blood pressure lowering | 8100550 (21.8%) | 151080 (9.81%) | 4486165 (23.8%) | 3449035 (21.7%) | 14270 (1.48%) |
| | Lipid lowering | 6475850 (17.4%) | 106190 (6.89%) | 3538865 (18.8%) | 2825350 (17.8%) | 5445 (0.56%) |
| | Anticoagulant | 1263445 (3.39%) | 37395 (2.43%) | 608995 (3.23%) | 616390 (3.88%) | 665 (0.07%) |
| | COPD[3] | 498145 (1.34%) | 19255 (1.25%) | 132030 (0.70%) | 319500 (2.01%) | 27360 (2.83%) |
| | HRT[5] | 513585 (1.38%) | 8425 (0.55%) | 348300 (1.85%) | 153415 (0.97%) | 3445 (0.36%) |
| **Clinically vulnerable status** | Neither | 27913540 (74.9%) | 1256045 (81.5%) | 13831795 (73.3%) | 11878650 (74.8%) | 947050 (98.0%) |
| | Vulnerable | 7373420 (19.8%) | 216525 (14.1%) | 3976360 (21.1%) | 3163620 (19.9%) | 16915 (1.75%) |
| | Extremely vulnerable | 1964415 (5.27%) | 67900 (4.41%) | 1060390 (5.62%) | 833940 (5.25%) | 2185 (0.23%) |

[1] All counts rounded to the nearest 5 and counts less than 10 displayed as '10'
[2] Acute myocardial infarction
[3] Combined oral contraceptives
[4] Venous Thromboembolic Event
[5] Hormone replacement therapy

Supplementary Table 2: Characterization of the population under study – booster cohort[1]

| | | Those receiving dose 2 and eligible for booster analysis | Vaccination status during booster follow-up | | | |
|---|---|---|---|---|---|---|
| | | | Remains unvaccinated | ChAdOx1 | BNT-162b2 | mRNA-1273 |
| **N (%)** | All | 35854085 | 27723925 (77.3%) | 3745 (0.01%) | 5600360 (15.6%) | 2526055 (7.05%) |
| **Sex (%)** | Male | 17169775 (47.9%) | 12991955 (46.9%) | 1290 (34.5%) | 2833710 (16.5%) | 1342820 (53.2%) |
| | Female | 18684315 (52.1%) | 14731975 (53.1%) | 2455 (65.5%) | 2766650 (14.8%) | 1183235 (46.8%) |
| **Age in years (%)** | 18-29 | 5045225 (14.1%) | 3216800 (11.6%) | 180 (4.81%) | 1226665 (24.3%) | 601580 (23.8%) |
| | 30-39 | 5641140 (15.7%) | 3428150 (12.4%) | 300 (8.01%) | 1473775 (26.1%) | 738915 (29.2%) |
| | 40-49 | 5844900 (16.3%) | 3815820 (13.8%) | 765 (20.4%) | 1319555 (22.6%) | 708760 (28.1%) |
| | 50-59 | 6741645 (18.8%) | 5650605 (20.4%) | 950 (25.4%) | 751325 (11.1%) | 338765 (13.4%) |
| | 60-69 | 5473820 (15.3%) | 4867890 (17.6%) | 770 (20.6%) | 477750 (8.73%) | 127410 (5.04%) |
| | 70-79 | 4471255 (12.5%) | 4205415 (15.2%) | 450 (12.0%) | 257065 (5.75%) | 8325 (0.33%) |
| | 80-89 | 2134695 (5.95%) | 2054905 (7.41%) | 235 (6.28%) | 77640 (3.64%) | 1915 (0.08%) |
| | 90+ | 501410 (1.40%) | 484340 (1.75%) | 95 (2.54%) | 16590 (3.31%) | 385 (0.02%) |
| **Ethnicity (%)** | Asian | 2722170 (7.59%) | 2117865 (7.64%) | 180 (4.81%) | 440605 (16.2%) | 163520 (6.47%) |
| | Black | 927395 (2.59%) | 784285 (2.83%) | 205 (5.47%) | 106585 (11.5%) | 36320 (1.44%) |
| | Mixed | 435310 (1.21%) | 320770 (1.16%) | 50 (1.34%) | 79925 (18.4%) | 34565 (1.37%) |
| | Other | 776290 (2.17%) | 552145 (1.99%) | 70 (1.87%) | 156485 (20.2%) | 67590 (2.68%) |
| | White | 30168350 (84.1%) | 23366285 (84.3%) | 3190 (85.2%) | 4655380 (15.4%) | 2143495 (84.9%) |
| | Unknown | 595945 (1.66%) | 422255 (1.52%) | 30 (0.80%) | 115675 (19.4%) | 57985 (2.30%) |
| | Missing | 228620 (0.64%) | 160310 (0.58%) | 15 (0.40%) | 45715 (20%) | 22580 (0.89%) |
| **Index of multiple deprivation (%) (lowest: most deprived; highest: last deprived).** | 1-2 | 6052805 (16.9%) | 4959800 (17.9%) | 625 (16.7%) | 781095 (12.9%) | 311285 (12.3%) |
| | 3-4 | 6764950 (18.9%) | 5309215 (19.1%) | 810 (21.6%) | 1007875 (14.9%) | 447050 (17.7%) |
| | 5-6 | 7190900 (20.1%) | 5556055 (20.0%) | 755 (20.2%) | 1125485 (15.7%) | 508605 (20.1%) |
| | 7-8 | 7354000 (20.5%) | 5618345 (20.3%) | 715 (19.1%) | 1183555 (16.1%) | 551385 (21.8%) |
| | 9-10 | 7383905 (20.6%) | 5499520 (19.8%) | 760 (20.3%) | 1285170 (17.4%) | 598455 (23.7%) |
| | Missing | 1107525 (3.09%) | 780995 (2.82%) | 75 (2.00%) | 217180 (19.6%) | 109275 (4.33%) |
| **Smoking status (%)** | Current smoker | 5111905 (14.3%) | 4070290 (14.7%) | 520 (13.9%) | 725560 (14.2%) | 315535 (12.5%) |
| | Former smoker | 9034425 (25.2%) | 7401730 (26.7%) | 1020 (27.2%) | 1157605 (12.8%) | 474070 (18.8%) |
| | Never smoker | 20645195 (57.6%) | 15571450 (56.2%) | 2165 (57.8%) | 3464350 (16.8%) | 1607230 (63.6%) |
| | Missing | 1062560 (2.96%) | 680455 (2.45%) | 40 (1.07%) | 252845 (23.8%) | 129220 (5.12%) |
| **Medical history (%)** | AMI[2] | 1080570 (3.01%) | 1010395 (3.64%) | 175 (4.67%) | 62640 (5.8%) | 7360 (0.29%) |
| | Diabetes | 3708200 (10.3%) | 3404425 (12.3%) | 570 (15.2%) | 256510 (6.92%) | 46695 (1.85%) |
| | Depression | 7939005 (22.1%) | 6435965 (23.2%) | 1200 (32.0%) | 1063075 (13.4%) | 438765 (17.4%) |
| | Obesity | 4044025 (11.3%) | 3469750 (12.5%) | 665 (17.8%) | 423890 (10.5%) | 149720 (5.93%) |
| | Cancer | 6774645 (18.9%) | 5340045 (19.3%) | 960 (25.6%) | 1007575 (14.9%) | 426065 (16.9%) |
| | COPD[3] | 1419540 (3.96%) | 1325105 (4.78%) | 315 (8.41%) | 84390 (5.94%) | 9730 (0.39%) |
| | Liver disease | 193475 (0.54%) | 173765 (0.63%) | 60 (1.60%) | 16605 (8.58%) | 3045 (0.12%) |
| | Chronic kidney disease | 2676310 (7.46%) | 2483740 (8.96%) | 570 (15.2%) | 168215 (6.29%) | 23785 (0.94%) |
| | Dementia | 466130 (1.30%) | 440500 (1.59%) | 130 (3.47%) | 24350 (5.22%) | 1150 (0.05%) |
| | All stroke | 771180 (2.15%) | 718630 (2.59%) | 175 (4.67%) | 46355 (6.01%) | 6020 (0.24%) |
| | All VTE[4] | 582620 (1.62%) | 530050 (1.91%) | 165 (4.41%) | 43880 (7.53%) | 8525 (0.34%) |
| | Thrombophilia | 38600 (0.11%) | 31925 (0.12%) | 15 (0.40%) | 5125 (13.3%) | 1535 (0.06%) |
| **Major surgery in the last year (%)** | | 3363130 (9.38%) | 3363130 (9.38%) | 2759995 (9.96%) | 720 (19.2%) | 446625 (13.3%) |
| **Number of unique diseases in the last year (%)** | 0 | 28248440 (78.8%) | 21291805 (76.8%) | 2430 (64.9%) | 4731060 (16.7%) | 2223145 (88.0%) |
| | 1-5 | 7488270 (20.9%) | 6324715 (22.8%) | 1270 (33.9%) | 860415 (11.5%) | 301870 (11.9%) |
| | 6+ | 117375 (0.33%) | 107410 (0.39%) | 40 (1.07%) | 8890 (7.57%) | 1035 (0.04%) |
| **Region (%)** | Northwest | 4518830 (12.6%) | 3631650 (13.1%) | 645 (17.2%) | 636445 (14.1%) | 250090 (9.90%) |
| | Southeast | 5527960 (15.4%) | 4181075 (15.1%) | 550 (14.7%) | 1045550 (18.9%) | 300785 (11.9%) |
| | London | 4878535 (13.6%) | 3589390 (12.9%) | 945 (25.2%) | 902705 (18.5%) | 385495 (15.3%) |
| | East of England | 3348135 (9.34%) | 2579335 (9.30%) | 230 (6.14%) | 485170 (14.5%) | 283400 (11.2%) |
| | Southwest | 3094010 (8.63%) | 2355545 (8.50%) | 295 (7.88%) | 481300 (15.6%) | 256870 (10.2%) |
| | Yorkshire and the Humber | 3484770 (9.72%) | 2631800 (9.49%) | 240 (6.41%) | 551330 (15.8%) | 301400 (11.9%) |
| | East Midlands | 3454285 (9.63%) | 2747485 (9.91%) | 225 (6.01%) | 467455 (13.5%) | 239120 (9.47%) |
| | West Midlands | 2608560 (7.28%) | 2103480 (7.59%) | 150 (4.01%) | 343980 (13.2%) | 160950 (6.37%) |
| | Northeast | 3547885 (9.90%) | 2820950 (10.2%) | 280 (7.48%) | 534875 (15.1%) | 191780 (7.59%) |
| | Missing | 1391110 (3.88%) | 1083205 (3.91%) | 185 (4.94%) | 151555 (10.9%) | 156165 (6.18%) |
| **Prior COVID-19 at index date** | | 2792305 (7.79%) | 2792305 (7.79%) | 2070735 (7.47%) | 340 (9.08%) | 494320 (17.7%) |
| **Medications taken in the last 3 months** | Antiplatelet | 2309730 (6.44%) | 2154415 (7.77%) | 345 (9.21%) | 138925 (6.01%) | 16045 (0.64%) |
| | Blood pressure lowering | 8040690 (22.4%) | 7340975 (26.5%) | 1075 (28.7%) | 576125 (7.17%) | 122515 (4.85%) |
| | Lipid lowering | 6461885 (18.0%) | 5958370 (21.5%) | 770 (20.6%) | 433580 (6.71%) | 69165 (2.74%) |
| | Anticoagulant | 1261465 (3.52%) | 1182670 (4.27%) | 220 (5.87%) | 71550 (5.67%) | 7025 (0.28%) |
| | COCP[3] | 468320 (1.31%) | 274460 (0.99%) | 30 (0.80%) | 129735 (27.7%) | 64095 (2.54%) |
| | HRT[5] | 536220 (1.50%) | 440870 (1.59%) | 135 (3.60%) | 67670 (12.6%) | 27545 (1.09%) |
| **Clinically vulnerable status** | Neither | 26773975 (74.7%) | 19342955 (69.8%) | 2275 (60.8%) | 464250 (6.47%) | 2429060 (96.2%) |
| | Vulnerable | 7178100 (20.0%) | 6623400 (23.9%) | 780 (20.8%) | 136425 (7.17%) | 89670 (3.55%) |
| | Extremely vulnerable | 1902015 (5.30%) | 1757580 (6.34%) | 685 (18.3%) | 4999685 (18.7%) | 7325 (0.29%) |

Supplementary Table 3: Definitions of the after-vaccination period and the corresponding period before or without vaccination in eligible individuals. Each comparison group included only individuals eligible to receive the vaccine brand and dose under consideration.

| | Analysis | Post-vaccination | No vaccination |
|---|---|---|---|
| Dose 1 | ChAdOx1 | Post first dose of ChAdOx1 | Unvaccinated or prior to the first dose of any vaccine |
| | BNT-162b2 | Post first dose of BNT-162b2 | |
| | mRNA-1273 | Post first dose of mRNA-1273 | |
| Dose 2 | ChAdOx1 | Post second dose of ChAdOx1, following a first dose of ChAdOx1 | Between first dose of ChAdOx1 and second dose vaccination of any brand |
| | BNT-162b2 | Post second dose of BNT-162b2, following a first dose of BNT-162b2 | Between first dose of BNT-162b2 and second dose vaccination of any brand |
| | mRNA-1273 | Post second dose of mRNA-1273, following a first dose of mRNA-1273 | Between first dose of mRNA-1273 and second dose vaccination of any brand |
| Booster | Any booster dose (vs. ChAdOx1 primary course) | Post any mRNA booster vaccination (BNT-162b2 or mRNA-1273), following a primary course of ChAdOx1 | Between second dose of ChAdOx1 and booster dose vaccination, following a primary course of ChAdOx1 |
| | Any booster dose (vs. BNT-162b2 primary course) | Post any mRNA booster vaccination (BNT-162b2 or mRNA-1273), following a primary course of BNT-162b2 | Between second dose of ChAdOx1 and booster dose vaccination, following a primary course of BNT-162b2 |
| | Any booster dose (vs. mRNA primary course) | Post any mRNA booster vaccination (BNT-162b2 or mRNA-1273), following a primary course of either BNT-162b2 or mRNA-1273 | Between second dose of ChAdOx1 and booster dose vaccination, following a primary course of either BNT-162b2 or mRNA-1273 |
| | Any booster dose (vs. Any primary course) | Post any mRNA booster vaccination (BNT-162b2 or mRNA-1273), following a primary course of one of BNT-162b2 or mRNA-1273 or ChAdOx1 | Between second dose of ChAdOx1 and booster dose vaccination, following a primary course of one of BNT-162b2 or mRNA-1273 or ChAdOx1 |
| | BNT-162b2 booster dose (vs. ChAdOx1 primary course) | Post BNT-162b2 booster vaccination, following a primary course of ChAdOx1 | Between second dose of ChAdOx1 and booster dose vaccination, following a primary course of ChAdOx1 |
| | BNT-162b2 booster dose (vs. BNT-162b2 primary course) | Post BNT-162b2 booster vaccination, following a primary course of BNT-162b2 | Between second dose of BNT-162b2 and booster dose vaccination, following a primary course of BNT-162b2 |
| | BNT-162b2 booster dose (vs. mRNA primary course) | Post BNT-162b2 booster vaccination, following a primary course of either BNT-162b2 or mRNA-1273 | Between second dose of either BNT-162b2 or mRNA-1273 and booster dose vaccination, following a primary course of either BNT-162b2 or mRNA-1273 |
| | BNT-162b2 booster dose (vs. Any primary course) | Post BNT-162b2 booster vaccination, following a primary course of one of BNT-162b2 or mRNA-1273 or ChAdOx1 | Between second dose of one of BNT-162b2 or mRNA-1273 or ChAdOx1 and booster dose vaccination, following a primary course of one of BNT-162b2 or mRNA-1273 or ChAdOx1 |
| | mRNA-1273 booster dose (vs. ChAdOx1 primary course) | Post mRNA-1273 booster vaccination, following a primary course of ChAdOx1 | Between second dose of ChAdOx1 and booster dose vaccination, following a primary course of ChAdOx1 |
| | mRNA-1273 booster dose (vs. BNT-162b2 primary course) | Post mRNA-1273 booster vaccination, following a primary course of BNT-162b2 | Between second dose of BNT-162b2 and booster dose vaccination, following a primary course of BNT-162b2 |
| | mRNA-1273 booster dose (vs. mRNA primary course) | Post mRNA-1273 booster vaccination, following a primary course of either BNT-162b2 or mRNA-1273 | Between second dose of either BNT-162b2 or mRNA-1273 and booster dose vaccination, following a primary course of either BNT-162b2 or mRNA-1273 |
| | mRNA-1273 booster dose (vs Any primary course) | Post mRNA-1273 booster vaccination, following a primary course of one of BNT-162b2 or mRNA-1273 or ChAdOx1 | Between second dose of one of BNT-162b2 or mRNA-1273 or ChAdOx1 and booster dose vaccination, following a primary course of one of BNT-162b2 or mRNA-1273 or ChAdOx1 |

Supplementary Table 4: Number of events, person-years and incidence rates for individual arterial and venous thrombotic events, and other cardiovascular events, for first and second vaccinations.

| Vaccination | Event | Events/100K person-years | | Incidence rate | |
|---|---|---|---|---|---|
| **Dose 1** | | **No vaccination** | **After first dose** | **No vaccination** | **After first dose** |
| **ChAdOx1** | AMI[2] | 37915/205.00 | 37675/ 95.87 | 184.95 | 392.97 |
| | Ischaemic stroke | 36720/205.00 | 35080/ 95.84 | 179.12 | 366.01 |
| | Pulmonary embolism | 11835/205.00 | 9695/ 95.81 | 57.73 | 101.19 |
| | Lower limb DVT[6] | 9075/205.00 | 7590/95.78 | 44.27 | 79.24 |
| | ICVT[7] | 370/205.00 | 290/96.28 | 1.80 | 3.01 |
| | Portal vein thrombosis | 185/205.00 | 175/95.79 | 0.90 | 1.83 |
| | Any thrombocytopenia | 1885/205.00 | 1760/95.98 | 9.20 | 18.34 |
| | SAH and HS[8] | 5235/205.00 | 4790/95.88 | 25.54 | 49.96 |
| | Mesenteric thrombus | 1515/205.00 | 1675/96.40 | 7.39 | 17.38 |
| | Myocarditis | 590/205.00 | 300/97.15 | 2.88 | 3.09 |
| | Pericarditis | 455/205.00 | 315/96.08 | 2.22 | 3.28 |
| | Composite arterial | 75655/205.00 | 73330/95.63 | 369.05 | 766.83 |
| | Composite venous | 21230/205.00 | 17525/96.09 | 103.56 | 182.39 |
| **BNT-162b2** | AMI[2] | 37915/205.00 | 28060/81.03 | 184.95 | 346.28 |
| | Ischaemic stroke | 36720/205.00 | 28535/81.06 | 179.12 | 352.04 |
| | Pulmonary embolism | 11835/205.00 | 6820/81.21 | 57.73 | 83.98 |
| | Lower limb DVT[6] | 9075/205.00 | 4895/81.37 | 44.27 | 60.16 |
| | ICVT[7] | 370/205.00 | 160/80.53 | 1.80 | 1.99 |
| | Portal vein thrombosis | 185/205.00 | 110/79.18 | 0.90 | 1.39 |
| | Any thrombocytopenia | 1885/205.00 | 1215/81.20 | 9.20 | 14.96 |
| | SAH and HS[8] | 5235/205.00 | 3575/81.60 | 25.54 | 43.81 |
| | Mesenteric thrombus | 1515/205.00 | 1310/81.12 | 7.39 | 16.15 |
| | Myocarditis | 590/205.00 | 315/81.44 | 2.88 | 3.87 |
| | Pericarditis | 455/205.00 | 270/78.92 | 2.22 | 3.42 |
| | Composite arterial | 75655/205.00 | 57050/80.99 | 369.05 | 704.38 |
| | Composite venous | 21230/205.00 | 11860/81.00 | 103.56 | 146.42 |
| **mRNA-1273** | AMI[2] | 37915/205.00 | 145/5.20 | 184.95 | 27.9 |
| | Ischaemic stroke | 36720/205.00 | 105/5.05 | 179.12 | 20.81 |
| | Pulmonary embolism | 11835/205.00 | 90/5.04 | 57.73 | 17.86 |
| | Lower limb DVT[6] | 9075/205.00 | 95/5.15 | 44.27 | 18.43 |
| | ICVT[7] | 370/205.00 | 10/5.09 | 1.80 | 1.97 |
| | Portal vein thrombosis | 185/205.00 | 10/4.33 | 0.90 | 2.31 |
| | Any thrombocytopenia | 1885/205.00 | 25/4.91 | 9.20 | 5.09 |
| | SAH and HS[8] | 5235/205.00 | 30/5.15 | 25.54 | 5.82 |
| | Mesenteric thrombus | 1515/205.00 | 10/5.03 | 7.39 | 1.99 |
| | Myocarditis | 590/205.00 | 50/4.91 | 2.88 | 10.19 |
| | Pericarditis | 455/205.00 | 20/5.26 | 2.22 | 3.8 |
| | Composite arterial | 75655/205.00 | 255/5.11 | 369.05 | 49.89 |
| | Composite venous | 21230/205.00 | 190/5.10 | 103.56 | 37.25 |
| **Doses 1 and 2** | | **After first dose** | **After second dose** | **After first dose** | **After second dose** |
| **ChAdOx1** | AMI[2] | 19475/41.29 | 36210/93.48 | 471.69 | 387.35 |
| | Ischaemic stroke | 19440/41.25 | 33805/93.53 | 471.26 | 361.44 |
| | Pulmonary embolism | 4785/41.26 | 9000/93.63 | 115.98 | 96.12 |
| | Lower limb DVT[6] | 3730/41.28 | 6975/93.64 | 90.35 | 74.49 |
| | ICVT[7] | 190/41.29 | 170/93.60 | 4.6 | 1.82 |
| | Portal vein thrombosis | 85/41.48 | 145/93.49 | 2.05 | 1.55 |
| | Any thrombocytopenia | 1130/41.16 | 1400/93.77 | 27.45 | 14.93 |
| | SAH and HS[8] | 2385/41.31 | 4360/93.64 | 57.74 | 46.56 |
| | Mesenteric thrombus | 765/41.24 | 1555/93.64 | 18.55 | 16.61 |
| | Myocarditis | 145/41.23 | 285/93.75 | 3.52 | 3.04 |
| | Pericarditis | 160/41.70 | 280/93.36 | 3.84 | 3 |
| | Composite arterial | 39425/41.23 | 70485/93.36 | 956.17 | 755.01 |
| | Composite venous | 8685/41.27 | 16120/93.60 | 210.46 | 172.22 |
| **BNT-162b2** | AMI[2] | 13125/33.65 | 27295/72.57 | 390 | 376.12 |
| | Ischaemic stroke | 13575/33.62 | 27930/72.61 | 403.81 | 384.67 |
| | Pulmonary embolism | 3070/33.64 | 6545/72.75 | 91.26 | 89.96 |
| | Lower limb DVT[6] | 2115/33.62 | 4725/72.64 | 62.9 | 65.04 |
| | ICVT[7] | 70/33.60 | 145/73.05 | 2.08 | 1.98 |
| | Portal vein thrombosis | 50/33.67 | 105/72.40 | 1.48 | 1.45 |
| | Any thrombocytopenia | 675/33.70 | 1125/72.62 | 20.03 | 15.49 |
| | SAH and HS[8] | 1515/33.67 | 3440/72.72 | 45 | 47.31 |
| | Mesenteric thrombus | 565/33.70 | 1225/72.67 | 16.76 | 16.86 |
| | Myocarditis | 120/33.82 | 280/72.58 | 3.55 | 3.86 |
| | Pericarditis | 100/33.54 | 255/72.52 | 2.98 | 3.52 |
| | Composite arterial | 26975/33.63 | 55620/72.44 | 802.22 | 767.8 |
| | Composite venous | 5260/33.67 | 11400/72.69 | 156.22 | 156.83 |
| **mRNA-1273** | AMI[2] | 85/2.17 | 110/4.02 | 39.17 | 27.34 |
| | Ischaemic stroke | 70/2.16 | 90/4.06 | 32.39 | 22.14 |
| | Pulmonary embolism | 40/2.15 | 70/4.09 | 18.6 | 17.1 |
| | Lower limb DVT[6] | 50/2.17 | 60/4.03 | 23.04 | 14.88 |
| | ICVT[7] | 10/2.20 | 10/4.01 | 4.54 | 2.5 |
| | Portal vein thrombosis | 10/2.25 | 10/4.05 | 4.44 | 2.47 |
| | Any thrombocytopenia | 20/2.17 | 15/4.11 | 9.21 | 3.65 |
| | SAH and HS[8] | 10/2.29 | 25/3.99 | 4.37 | 6.27 |
| | Mesenteric thrombus | 10/2.22 | 10/4.19 | 4.51 | 2.39 |
| | Myocarditis | 15/2.13 | 45/4.07 | 7.04 | 11.05 |
| | Pericarditis | 10/2.19 | 15/4.07 | 4.57 | 3.68 |
| | Composite arterial | 160/2.17 | 200/4.06 | 73.66 | 49.24 |
| | Composite venous | 90/2.18 | 135/4.06 | 41.37 | 33.29 |

Supplementary Table 5: Number of events, person-years and incidence rates for individual arterial and venous thrombotic events, and other cardiovascular events, for booster vaccination with BNT-162b2.

| Vaccination | Event | Events/100K person-years | | Incidence rate | |
|---|---|---|---|---|---|
| | | Primary course only | After booster vaccination | Primary course only | After booster vaccination |
| **Primary course** | | | | | |
| **Any** | AMI[2] | 77890/182.46 | 19640/ 38.88 | 426.9 | 505.18 |
| | Ischaemic stroke | 78120/182.46 | 20050/ 38.92 | 428.16 | 515.15 |
| | Pulmonary embolism | 18580/182.66 | 4710/ 39.05 | 101.72 | 120.61 |
| | Lower limb DVT[6] | 13605/182.59 | 3105/ 39.05 | 74.51 | 79.51 |
| | ICVT[7] | 360/182.19 | 65/ 38.47 | 1.98 | 1.69 |
| | Portal vein thrombosis | 300/183.01 | 50/39.51 | 1.64 | 1.27 |
| | Any thrombocytopenia | 3340/182.60 | 680/ 39.10 | 18.29 | 17.39 |
| | SAH and HS[8] | 9110/182.67 | 2455/ 38.97 | 49.87 | 62.99 |
| | Mesenteric thrombus | 3300/182.62 | 845/ 39.49 | 18.07 | 21.4 |
| | Myocarditis | 675/182.54 | 120/ 39.15 | 3.7 | 3.06 |
| | Pericarditis | 615/182.84 | 110/ 39.39 | 3.36 | 2.79 |
| | Composite arterial | 157235/182.21 | 39735/ 38.72 | 862.93 | 1026.14 |
| | Composite venous | 32545/182.54 | 7855/ 38.97 | 178.29 | 201.54 |
| **BNT-162b2/mRNA-1273** | AMI[2] | 33945/80.40 | 10435/19.11 | 422.22 | 546 |
| | Ischaemic stroke | 35745/80.39 | 10985/19.11 | 444.63 | 574.85 |
| | Pulmonary embolism | 8010/80.43 | 2410/19.13 | 99.58 | 125.97 |
| | Lower limb DVT[6] | 5660/80.46 | 1550/19.23 | 70.34 | 80.59 |
| | ICVT[7] | 165/80.98 | 35/19.70 | 2.04 | 1.78 |
| | Portal vein thrombosis | 125/80.42 | 20/19.27 | 1.55 | 1.04 |
| | Any thrombocytopenia | 1535/80.42 | 340/19.18 | 19.09 | 17.72 |
| | SAH and HS[8] | 4115/80.43 | 1370/19.14 | 51.16 | 71.58 |
| | Mesenteric thrombus | 1505/80.57 | 485/19.22 | 18.68 | 25.23 |
| | Myocarditis | 345/80.19 | 60/19.17 | 4.3 | 3.13 |
| | Pericarditis | 295/80.22 | 55/19.26 | 3.68 | 2.86 |
| | Composite arterial | 70215/80.31 | 21425/19.04 | 874.28 | 1125.34 |
| | Composite venous | 13830/80.45 | 3975/19.15 | 171.91 | 207.62 |
| **ChAdOx1** | AMI[2] | 43945/102.06 | 9200/ 19.79 | 430.59 | 464.92 |
| | Ischaemic stroke | 42375/102.05 | 9065/ 19.78 | 415.23 | 458.31 |
| | Pulmonary embolism | 10570/102.23 | 2300/ 19.81 | 103.4 | 116.08 |
| | Lower limb DVT[6] | 7945/102.12 | 1555/ 19.82 | 77.8 | 78.44 |
| | ICVT[7] | 195/101.89 | 30/ 19.53 | 1.91 | 1.54 |
| | Portal vein thrombosis | 175/102.33 | 30/ 19.48 | 1.71 | 1.54 |
| | Any thrombocytopenia | 1800/102.15 | 340/ 19.81 | 17.62 | 17.16 |
| | SAH and HS[8] | 4990/102.14 | 1085/ 19.83 | 48.85 | 54.71 |
| | Mesenteric thrombus | 1795/102.18 | 360/ 19.82 | 17.57 | 18.16 |
| | Myocarditis | 330/102.26 | 60/ 20.15 | 3.23 | 2.98 |
| | Pericarditis | 320/102.42 | 55/ 19.94 | 3.12 | 2.76 |
| | Composite arterial | 87020/101.93 | 18310/ 19.76 | 853.73 | 926.59 |
| | Composite venous | 18715/102.13 | 3880/ 19.81 | 183.25 | 195.83 |
| **BNT-162b2** | AMI[2] | 33835/76.71 | 10430/18.87 | 441.1 | 552.7 |
| | Ischaemic stroke | 35650/76.69 | 10980/18.83 | 464.85 | 583.27 |
| | Pulmonary embolism | 7945/76.84 | 2405/18.92 | 103.39 | 127.08 |
| | Lower limb DVT[6] | 5600/76.76 | 1545/18.91 | 72.96 | 81.7 |
| | ICVT[7] | 160/76.41 | 35/18.65 | 2.09 | 1.88 |
| | Portal vein thrombosis | 120/76.40 | 20/18.89 | 1.57 | 1.06 |
| | Any thrombocytopenia | 1520/76.94 | 340/18.95 | 19.75 | 17.94 |
| | SAH and HS[8] | 4090/76.62 | 1365/18.89 | 53.38 | 72.27 |
| | Mesenteric thrombus | 1505/76.80 | 485/19.09 | 19.6 | 25.41 |
| | Myocarditis | 300/77.04 | 60/19.10 | 3.89 | 3.14 |
| | Pericarditis | 280/76.49 | 55/18.67 | 3.66 | 2.95 |
| | Composite arterial | 70005/76.60 | 21415/18.78 | 913.88 | 1140.44 |
| | Composite venous | 13700/76.75 | 3965/18.91 | 178.51 | 209.65 |

[6] Deep venous thrombosis
[7] Intracranial venous thrombosis
[8] Subarachnoid haemorrhage and haemorrhagic stroke

Supplementary Table 6: Number of events, person-years and incidence rates for individual arterial and venous thrombotic events, and other cardiovascular events, for booster vaccination with mRNA-1273.

| Vaccination | Event | Events/100K person-years | | Incidence rate | |
|---|---|---|---|---|---|
| | | Primary course only | After booster vaccination | Primary course only | After booster vaccination |
| **Primary course** | | | | | |
| **Any** | AMI[2] | 77890/182.41 | 1830/7.47 | 427.01 | 244.92 |
| | Ischaemic stroke | 78120/182.41 | 1345/7.46 | 428.26 | 180.32 |
| | Pulmonary embolism | 18580/182.60 | 430/7.47 | 101.75 | 57.55 |
| | Lower limb DVT[6] | 13605/182.66 | 320/7.51 | 74.48 | 42.59 |
| | ICVT[7] | 360/183.47 | 10/7.01 | 1.96 | 1.43 |
| | Portal vein thrombosis | 300/183.23 | 10/7.50 | 1.64 | 1.33 |
| | Any thrombocytopenia | 3340/182.65 | 50/7.41 | 18.29 | 6.75 |
| | SAH and HS[8] | 9110/182.86 | 150/7.42 | 49.82 | 20.21 |
| | Mesenteric thrombus | 3300/182.68 | 55/7.48 | 18.06 | 7.35 |
| | Myocarditis | 675/182.13 | 20/7.35 | 3.71 | 2.72 |
| | Pericarditis | 615/182.71 | 10/7.51 | 3.37 | 1.33 |
| | Composite arterial | 157235/182.22 | 3200/7.46 | 862.88 | 429.07 |
| | Composite venous | 32545/182.56 | 760/7.47 | 178.27 | 101.79 |
| **BNT-162b2/mRNA-1273** | AMI[2] | 33945/80.39 | 475/ 2.25 | 422.25 | 210.88 |
| | Ischaemic stroke | 35745/80.45 | 385/ 2.22 | 444.29 | 173.12 |
| | Pulmonary embolism | 8010/80.47 | 115/ 2.27 | 99.54 | 50.64 |
| | Lower limb DVT[6] | 5660/80.38 | 70/ 2.24 | 70.42 | 31.28 |
| | ICVT[7] | 165/80.45 | 10/ 2.23 | 2.05 | 4.48 |
| | Portal vein thrombosis | 125/79.95 | 10/ 2.26 | 1.56 | 4.43 |
| | Any thrombocytopenia | 1535/80.60 | 15/ 2.22 | 19.04 | 6.77 |
| | SAH and HS[8] | 4115/80.55 | 40/ 2.23 | 51.09 | 17.94 |
| | Mesenteric thrombus | 1505/80.45 | 20/ 2.24 | 18.71 | 8.92 |
| | Myocarditis | 345/80.61 | 10/ 2.20 | 4.28 | 4.54 |
| | Pericarditis | 295/80.23 | 10/ 2.28 | 3.68 | 4.39 |
| | Composite arterial | 70215/80.29 | 870/ 2.23 | 874.52 | 389.78 |
| | Composite venous | 13830/80.45 | 185/ 2.24 | 171.91 | 82.76 |
| **ChAdOx1** | AMI[2] | 43945/102.03 | 1355/5.23 | 430.71 | 258.97 |
| | Ischaemic stroke | 42375/102.03 | 960/5.24 | 415.31 | 183.38 |
| | Pulmonary embolism | 10570/102.13 | 310/5.21 | 103.49 | 59.49 |
| | Lower limb DVT[6] | 7945/102.13 | 250/5.22 | 77.79 | 47.86 |
| | ICVT[7] | 195/101.99 | 10/5.23 | 1.91 | 1.91 |
| | Portal vein thrombosis | 175/102.06 | 10/5.31 | 1.71 | 1.88 |
| | Any thrombocytopenia | 1800/102.04 | 35/5.16 | 17.64 | 6.78 |
| | SAH and HS[8] | 4990/102.24 | 110/5.19 | 48.81 | 21.18 |
| | Mesenteric thrombus | 1795/102.21 | 35/5.23 | 17.56 | 6.69 |
| | Myocarditis | 330/102.45 | 15/5.11 | 3.22 | 2.93 |
| | Pericarditis | 320/102.16 | 10/5.24 | 3.13 | 1.91 |
| | Composite arterial | 87020/101.95 | 2325/5.21 | 853.59 | 446.61 |
| | Composite venous | 18715/102.12 | 575/5.24 | 183.27 | 109.82 |
| **BNT-162b2** | AMI[2] | 33835/76.67 | 470/ 2.02 | 441.28 | 233.01 |
| | Ischaemic stroke | 35650/76.71 | 380/ 2.02 | 464.71 | 188.46 |
| | Pulmonary embolism | 7945/76.75 | 110/ 2.03 | 103.52 | 54.2 |
| | Lower limb DVT[6] | 5600/76.69 | 65/ 2.03 | 73.03 | 31.96 |
| | ICVT[7] | 160/77.25 | 10/ 1.95 | 2.07 | 5.13 |
| | Portal vein thrombosis | 120/77.51 | 10/ 1.91 | 1.55 | 5.23 |
| | Any thrombocytopenia | 1520/76.91 | 15/ 2.02 | 19.76 | 7.44 |
| | SAH and HS[8] | 4090/76.69 | 40/ 2.01 | 53.33 | 19.91 |
| | Mesenteric thrombus | 1505/76.69 | 20/ 2.05 | 19.62 | 9.77 |
| | Myocarditis | 300/76.49 | 10/ 1.96 | 3.92 | 5.1 |
| | Pericarditis | 280/76.54 | 10/ 1.91 | 3.66 | 5.23 |
| | Composite arterial | 70005/76.59 | 860/ 2.02 | 914 | 426.6 |
| | Composite venous | 13700/76.80 | 180/2.04 | 178.38 | 88.15 |

Supplementary Table 7: Number of events, person-years and incidence rates for individual arterial and venous thrombotic events, and other cardiovascular events, for booster vaccination with BNT-162b2/ mRNA-1273.

| Vaccination | Event | Events/100K person-years | | Incidence rate | |
|---|---|---|---|---|---|
| | | Primary course only | After booster vaccination | Primary course only | After booster vaccination |
| **Primary course** | | | | | |
| **Any** | AMI[2] | 77890/182.46 | 21470/ 46.36 | 426.89 | 463.11 |
| | Ischaemic stroke | 78120/182.43 | 21400/ 46.34 | 428.21 | 461.82 |
| | Pulmonary embolism | 18580/182.75 | 5135/ 46.51 | 101.67 | 101.41 |
| | Lower limb DVT[6] | 13605/182.56 | 3425/ 46.43 | 74.52 | 73.77 |
| | ICVT[7] | 360/182.76 | 75/ 46.59 | 1.97 | 1.61 |
| | Portal vein thrombosis | 300/183.14 | 60/ 46.34 | 1.64 | 1.29 |
| | Any thrombocytopenia | 3340/182.74 | 735/ 46.32 | 18.28 | 15.87 |
| | SAH and HS[8] | 9110/182.47 | 2605/ 46.48 | 49.93 | 56.04 |
| | Mesenteric thrombus | 3300/182.58 | 900/ 46.49 | 18.07 | 19.36 |
| | Myocarditis | 675/182.65 | 145/ 46.56 | 3.7 | 3.11 |
| | Pericarditis | 615/182.49 | 125/ 46.32 | 3.37 | 2.7 |
| | Composite arterial | 157235/182.21 | 42935/ 46.20 | 862.93 | 929.36 |
| | Composite venous | 32545/182.58 | 8615/ 46.37 | 178.25 | 185.79 |
| **BNT-162b2/mRNA-1273** | AMI[2] | 33945/80.41 | 10915/21.37 | 422.15 | 510.79 |
| | Ischaemic stroke | 35745/80.43 | 11370/21.33 | 444.45 | 532.94 |
| | Pulmonary embolism | 8010/80.55 | 2525/21.45 | 99.45 | 117.73 |
| | Lower limb DVT[6] | 5660/80.52 | 1615/21.38 | 70.29 | 75.56 |
| | ICVT[7] | 165/80.54 | 40/21.93 | 2.05 | 1.82 |
| | Portal vein thrombosis | 125/80.52 | 25/21.40 | 1.55 | 1.17 |
| | Any thrombocytopenia | 1535/80.59 | 355/21.31 | 19.05 | 16.66 |
| | SAH and HS[8] | 4115/80.56 | 1405/21.44 | 51.08 | 65.53 |
| | Mesenteric thrombus | 1505/80.60 | 505/21.54 | 18.67 | 23.44 |
| | Myocarditis | 345/80.88 | 70/21.78 | 4.27 | 3.21 |
| | Pericarditis | 295/80.13 | 60/21.37 | 3.68 | 2.81 |
| | Composite arterial | 70215/80.27 | 22300/21.25 | 874.73 | 1049.33 |
| | Composite venous | 13830/80.46 | 4165/21.39 | 171.88 | 194.72 |
| **ChAdOx1** | AMI[2] | 43945/102.06 | 10555/ 25.01 | 430.57 | 422.06 |
| | Ischaemic stroke | 42375/102.08 | 10030/ 25.00 | 415.12 | 401.21 |
| | Pulmonary embolism | 10570/102.15 | 2610/ 25.09 | 103.47 | 104.01 |
| | Lower limb DVT[6] | 7945/102.09 | 1805/ 25.07 | 77.82 | 71.99 |
| | ICVT[7] | 195/102.22 | 40/ 25.37 | 1.91 | 1.58 |
| | Portal vein thrombosis | 175/102.11 | 35/ 25.80 | 1.71 | 1.36 |
| | Any thrombocytopenia | 1800/102.22 | 380/ 25.10 | 17.61 | 15.14 |
| | SAH and HS[8] | 4990/102.17 | 1195/ 25.15 | 48.84 | 47.51 |
| | Mesenteric thrombus | 1795/102.08 | 395/ 25.10 | 17.58 | 15.74 |
| | Myocarditis | 330/102.35 | 75/ 25.11 | 3.22 | 2.99 |
| | Pericarditis | 320/102.15 | 65/ 25.06 | 3.13 | 2.59 |
| | Composite arterial | 87020/101.95 | 20635/ 24.94 | 853.54 | 827.32 |
| | Composite venous | 18715/102.08 | 4455/ 25.08 | 183.33 | 177.61 |
| **BNT-162b2** | AMI[2] | 33835/76.69 | 10900/20.86 | 441.19 | 522.5 |
| | Ischaemic stroke | 35650/76.64 | 11360/20.88 | 465.17 | 544.02 |
| | Pulmonary embolism | 7945/76.83 | 2515/20.93 | 103.4 | 120.15 |
| | Lower limb DVT[6] | 5600/76.68 | 1610/20.80 | 73.03 | 77.41 |
| | ICVT[7] | 160/76.07 | 40/20.92 | 2.1 | 1.91 |
| | Portal vein thrombosis | 120/76.52 | 25/21.39 | 1.57 | 1.17 |
| | Any thrombocytopenia | 1520/76.61 | 355/20.93 | 19.84 | 16.96 |
| | SAH and HS[8] | 4090/76.70 | 1405/20.90 | 53.32 | 67.24 |
| | Mesenteric thrombus | 1505/76.67 | 505/20.89 | 19.63 | 24.18 |
| | Myocarditis | 300/76.28 | 65/21.07 | 3.93 | 3.09 |
| | Pericarditis | 280/76.51 | 60/20.81 | 3.66 | 2.88 |
| | Composite arterial | 70005/76.59 | 22275/20.79 | 913.99 | 1071.28 |
| | Composite venous | 13700/76.73 | 4145/20.96 | 178.54 | 197.8 |

Supplementary Table 8: Hazard ratios and 95% CIs for individual arterial and venous thrombotic events, and other cardiovascular events, by time since first dose vaccination with ChAdOx1.

| Outcome | | Age and Sex adjusted HR (95% CI) | | | | | | Fully Adjusted HR (95% CI) | | | | | |
|---|---|---|---|---|---|---|---|---|---|---|---|---|---|
| | | Week 1 | Week 2 | Weeks 3-4 | Weeks 5-12 | Weeks 13-24 | Weeks 25-26 | Week 1 | Week 2 | Weeks 3-4 | Weeks 5-12 | Weeks 13-24 | Weeks 25-26 |
| **Arterial** | Composite arterial | 0.90 (0.87,0.94) | 1.06 (1.02,1.10) | 1.09 (1.06,1.12) | 1.17 (1.15,1.19) | 1.26 (1.23,1.29) | 1.32 (1.28,1.37) | 0.84 (0.80,0.87) | 0.97 (0.93,1.01) | 0.97 (0.94,1.00) | 1.00 (0.97,1.02) | 0.99 (0.97,1.02) | 0.98 (0.95,1.02) |
| | AMI[2] | 0.94 (0.89,0.99) | 1.03 (0.98,1.09) | 1.11 (1.06,1.16) | 1.22 (1.18,1.25) | 1.31 (1.27,1.35) | 1.41 (1.34,1.48) | 0.87 (0.82,0.92) | 0.95 (0.89,1.00) | 0.99 (0.95,1.04) | 1.03 (1.00,1.06) | 1.02 (0.98,1.05) | 1.02 (0.97,1.08) |
| | Ischaemic stroke | 0.88 (0.83,0.93) | 1.06 (1.00,1.12) | 1.06 (1.01,1.10) | 1.12 (1.09,1.16) | 1.22 (1.18,1.27) | 1.27 (1.20,1.34) | 0.82 (0.77,0.87) | 0.97 (0.92,1.03) | 0.95 (0.91,0.99) | 0.97 (0.93,1.00) | 0.99 (0.95,1.03) | 0.97 (0.91,1.03) |
| **Venous** | Composite venous | 0.87 (0.80,0.94) | 1.11 (1.03,1.20) | 1.09 (1.03,1.15) | 1.06 (1.02,1.10) | 1.13 (1.09,1.18) | 1.05 (0.98,1.13) | 0.80 (0.74,0.87) | 1.02 (0.94,1.09) | 0.98 (0.93,1.04) | 0.92 (0.88,0.96) | 0.94 (0.90,0.98) | 0.84 (0.78,0.90) |
| | Lower limb DVT[6] | 0.89 (0.78,1.01) | 1.09 (0.97,1.23) | 1.14 (1.05,1.25) | 1.10 (1.03,1.16) | 1.11 (1.05,1.18) | 1.00 (0.90,1.11) | 0.82 (0.72,0.94) | 1.00 (0.89,1.13) | 1.04 (0.95,1.13) | 0.96 (0.90,1.02) | 0.93 (0.87,0.99) | 0.81 (0.73,0.90) |
| | Pulmonary embolism | 0.84 (0.76,0.93) | 1.03 (0.93,1.13) | 1.02 (0.94,1.10) | 1.03 (0.98,1.09) | 1.16 (1.09,1.22) | 1.10 (1.00,1.21) | 0.78 (0.70,0.86) | 0.94 (0.85,1.03) | 0.91 (0.85,0.99) | 0.90 (0.85,0.95) | 0.97 (0.91,1.02) | 0.88 (0.80,0.97) |
| | ICVT[7] | 1.39 (0.70,2.76) | 6.08 (4.21,8.79) | 4.09 (2.96,5.66) | 1.37 (1.00,1.87) | 1.49 (1.06,2.09) | 1.72 (0.94,3.14) | 1.37 (0.69,2.72) | 5.92 (4.07,8.63) | 3.91 (2.81,5.44) | 1.25 (0.91,1.73) | 1.34 (0.95,1.90) | 1.60 (0.88,2.91) |
| | Mesenteric thrombus | 0.64 (0.46,0.87) | 1.09 (0.84,1.40) | 1.07 (0.87,1.31) | 1.15 (0.99,1.33) | 1.38 (1.18,1.62) | 1.54 (1.21,1.97) | 0.56 (0.41,0.78) | 0.94 (0.72,1.22) | 0.91 (0.74,1.12) | 0.90 (0.77,1.06) | 0.98 (0.83,1.17) | 0.99 (0.77,1.29) |
| | Portal vein thrombosis | 0.86 (0.34,2.17) | 1.16 (0.52,2.57) | 1.06 (0.57,1.97) | 1.21 (0.82,1.79) | 1.51 (0.98,2.33) | 1.96 (0.96,4.02) | 0.61 (0.23,1.61) | 0.80 (0.35,1.84) | 0.70 (0.35,1.39) | 0.78 (0.49,1.24) | 0.76 (0.46,1.26) | 0.88 (0.39,2.01) |
| **Other** | Any thrombocytopenia | 1.45 (1.12,1.88) | 2.53 (2.05,3.12) | 2.00 (1.67,2.39) | 1.79 (1.57,2.04) | 2.16 (1.86,2.52) | 1.98 (1.52,2.58) | 1.22 (0.94,1.58) | 2.07 (1.67,2.58) | 1.60 (1.33,1.93) | 1.24 (1.08,1.43) | 1.28 (1.10,1.49) | 1.04 (0.80,1.36) |
| | SAH and HS[8] | 0.91 (0.78,1.06) | 0.99 (0.86,1.16) | 0.99 (0.89,1.11) | 1.06 (0.98,1.15) | 1.11 (1.02,1.20) | 1.02 (0.89,1.18) | 0.89 (0.76,1.04) | 0.97 (0.83,1.13) | 0.97 (0.86,1.09) | 1.02 (0.94,1.10) | 1.04 (0.95,1.13) | 0.94 (0.81,1.08) |
| | Myocarditis | 1.77 (1.06,2.96) | 0.61 (0.27,1.37) | 1.36 (0.90,2.05) | 1.01 (0.77,1.34) | 1.12 (0.87,1.45) | 1.22 (0.77,1.95) | 1.63 (0.98,2.72) | 0.56 (0.25,1.26) | 1.26 (0.84,1.90) | 0.93 (0.70,1.22) | 0.99 (0.77,1.28) | 1.04 (0.66,1.65) |
| | Pericarditis | 0.99 (0.50,1.96) | 1.85 (1.11,3.09) | 1.50 (0.98,2.30) | 1.53 (1.18,1.98) | 1.14 (0.88,1.49) | 1.17 (0.71,1.94) | 0.94 (0.48,1.86) | 1.74 (1.04,2.91) | 1.37 (0.89,2.11) | 1.37 (1.05,1.78) | 0.96 (0.73,1.26) | 0.97 (0.60,1.59) |

Supplementary Table 9: Hazard ratios and 95% CIs for individual arterial and venous thrombotic events, and other cardiovascular events, by time since first dose vaccination with BNT-162b2.

| Outcome | | Age and Sex adjusted HR (95% CI) | | | | | | Fully Adjusted HR (95% CI) | | | | | |
|---|---|---|---|---|---|---|---|---|---|---|---|---|---|
| | | Week 1 | Week 2 | Weeks 3-4 | Weeks 5-12 | Weeks 13-24 | Weeks 25-26 | Week 1 | Week 2 | Weeks 3-4 | Weeks 5-12 | Weeks 13-24 | Weeks 25-26 |
| **Arterial** | Composite arterial | 0.80 (0.76,0.84) | 0.90 (0.86,0.95) | 0.96 (0.93,0.99) | 1.04 (1.01,1.06) | 1.13 (1.10,1.15) | 1.13 (1.09,1.18) | 0.73 (0.69,0.76) | 0.80 (0.77,0.84) | 0.83 (0.80,0.86) | 0.85 (0.83,0.87) | 0.90 (0.88,0.93) | 0.91 (0.87,0.94) |
| | AMI[2] | 0.88 (0.83,0.94) | 1.00 (0.94,1.06) | 1.01 (0.96,1.06) | 1.12 (1.09,1.16) | 1.22 (1.18,1.26) | 1.25 (1.18,1.31) | 0.76 (0.71,0.82) | 0.85 (0.79,0.90) | 0.83 (0.79,0.87) | 0.88 (0.85,0.91) | 0.93 (0.89,0.96) | 0.95 (0.89,1.00) |
| | Ischaemic stroke | 0.73 (0.68,0.77) | 0.83 (0.78,0.88) | 0.90 (0.86,0.95) | 0.95 (0.92,0.98) | 1.04 (1.01,1.08) | 1.04 (0.98,1.10) | 0.69 (0.65,0.74) | 0.78 (0.74,0.84) | 0.84 (0.80,0.88) | 0.85 (0.82,0.88) | 0.92 (0.88,0.96) | 0.91 (0.86,0.97) |
| **Venous** | Composite venous | 0.72 (0.65,0.79) | 0.86 (0.78,0.94) | 0.83 (0.78,0.89) | 0.89 (0.86,0.93) | 0.97 (0.93,1.01) | 0.90 (0.84,0.98) | 0.66 (0.60,0.73) | 0.79 (0.72,0.87) | 0.75 (0.70,0.81) | 0.79 (0.76,0.82) | 0.85 (0.81,0.88) | 0.78 (0.72,0.84) |
| | Lower limb DVT[6] | 0.75 (0.64,0.87) | 0.91 (0.78,1.05) | 0.81 (0.72,0.90) | 0.89 (0.84,0.95) | 0.92 (0.87,0.98) | 0.88 (0.78,0.99) | 0.71 (0.60,0.83) | 0.85 (0.73,0.98) | 0.75 (0.67,0.84) | 0.80 (0.75,0.86) | 0.82 (0.77,0.87) | 0.78 (0.69,0.88) |
| | Pulmonary embolism | 0.68 (0.60,0.78) | 0.80 (0.71,0.90) | 0.82 (0.75,0.90) | 0.88 (0.83,0.93) | 1.01 (0.95,1.06) | 0.93 (0.83,1.03) | 0.63 (0.55,0.72) | 0.73 (0.65,0.83) | 0.74 (0.68,0.81) | 0.77 (0.73,0.82) | 0.87 (0.82,0.93) | 0.80 (0.72,0.89) |
| | ICVT[7] | 0.17 (0.02,1.22) | 1.91 (1.03,3.54) | 1.21 (0.69,2.11) | 0.90 (0.63,1.28) | 1.29 (0.96,1.73) | 1.15 (0.60,2.20) | 0.16 (0.02,1.14) | 1.68 (0.89,3.14) | 1.07 (0.61,1.89) | 0.79 (0.55,1.14) | 1.12 (0.82,1.53) | 1.02 (0.53,1.99) |
| | Mesenteric thrombus | 0.62 (0.45,0.88) | 0.79 (0.59,1.07) | 0.82 (0.65,1.03) | 1.00 (0.86,1.16) | 1.22 (1.04,1.43) | 1.11 (0.85,1.45) | 0.55 (0.39,0.77) | 0.69 (0.51,0.94) | 0.67 (0.53,0.84) | 0.75 (0.64,0.89) | 0.87 (0.72,1.03) | 0.76 (0.57,1.00) |
| | Portal vein thrombosis | 0.89 (0.28,2.85) | 0.52 (0.13,2.15) | 0.82 (0.35,1.92) | 1.41 (0.94,2.11) | 1.55 (1.04,2.32) | 0.61 (0.21,1.74) | 0.60 (0.17,2.15) | 0.39 (0.09,1.71) | 0.59 (0.22,1.55) | 1.05 (0.64,1.71) | 1.17 (0.71,1.93) | 0.45 (0.13,1.50) |
| **Other** | Any thrombocytopenia | 1.61 (1.24,2.08) | 0.96 (0.69,1.36) | 1.47 (1.19,1.81) | 1.43 (1.24,1.64) | 1.66 (1.45,1.90) | 1.67 (1.30,2.14) | 1.25 (0.96,1.64) | 0.74 (0.52,1.04) | 1.09 (0.87,1.35) | 0.97 (0.83,1.13) | 1.08 (0.92,1.25) | 1.03 (0.79,1.34) |
| | SAH and HS[8] | 0.64 (0.53,0.78) | 0.66 (0.54,0.80) | 0.78 (0.68,0.89) | 0.88 (0.81,0.96) | 1.02 (0.94,1.11) | 1.02 (0.87,1.18) | 0.64 (0.53,0.78) | 0.65 (0.54,0.80) | 0.77 (0.68,0.88) | 0.88 (0.80,0.96) | 1.03 (0.94,1.13) | 1.04 (0.89,1.21) |
| | Myocarditis | 2.21 (1.38,3.55) | 1.52 (0.86,2.66) | 1.15 (0.72,1.81) | 1.47 (1.16,1.85) | 1.26 (1.00,1.58) | 1.96 (1.33,2.89) | 2.05 (1.28,3.29) | 1.41 (0.81,2.48) | 1.07 (0.67,1.70) | 1.34 (1.05,1.70) | 1.12 (0.89,1.41) | 1.66 (1.12,2.47) |
| | Pericarditis | 0.98 (0.46,2.07) | 0.85 (0.38,1.93) | 1.07 (0.63,1.82) | 1.87 (1.46,2.40) | 1.75 (1.38,2.21) | 0.89 (0.44,1.78) | 0.87 (0.41,1.86) | 0.77 (0.34,1.74) | 0.95 (0.56,1.63) | 1.61 (1.24,2.08) | 1.50 (1.17,1.92) | 0.74 (0.37,1.50) |

Supplementary Table 10: Hazard ratios and 95% CIs for individual arterial and venous thrombotic events, and other cardiovascular events, by time since first dose vaccination with mRNA-1273.

| Outcome | | Age and Sex adjusted HR (95% CI) | | | | | | Fully Adjusted HR (95% CI) | | | | | |
|---|---|---|---|---|---|---|---|---|---|---|---|---|---|
| | | Week 1 | Week 2 | Weeks 3-4 | Weeks 5-12 | Weeks 13-24 | Weeks 25-26 | Week 1 | Week 2 | Weeks 3-4 | Weeks 5-12 | Weeks 13-24 | Weeks 25-26 |
| **Arterial** | Composite arterial | 0.74 (0.41, 1.34) | 0.60 (0.31, 1.16) | 1.09 (0.77, 1.55) | 0.76 (0.61, 0.94) | 0.66 (0.54, 0.80) | 0.89 (0.58, 1.37) | 0.74 (0.41, 1.34) | 0.60 (0.31, 1.16) | 1.08 (0.76, 1.53) | 0.75 (0.60, 0.93) | 0.64 (0.52, 0.78) | 0.86 (0.56, 1.32) |
| | AMI[2] | 0.69 (0.31, 1.53) | 0.69 (0.31, 1.53) | 1.16 (0.74, 1.80) | 0.67 (0.50, 0.91) | 0.69 (0.54, 0.90) | 0.63 (0.33, 1.22) | 0.71 (0.32, 1.58) | 0.69 (0.31, 1.55) | 1.17 (0.75, 1.82) | 0.68 (0.50, 0.92) | 0.68 (0.53, 0.89) | 0.60 (0.31, 1.16) |
| | Ischaemic stroke | 0.96 (0.43, 2.14) | 0.48 (0.15, 1.49) | 0.89 (0.49, 1.62) | 0.82 (0.59, 1.13) | 0.56 (0.40, 0.79) | 1.28 (0.72, 2.26) | 0.94 (0.42, 2.10) | 0.48 (0.15, 1.49) | 0.89 (0.49, 1.62) | 0.80 (0.57, 1.12) | 0.53 (0.38, 0.76) | 1.18 (0.66, 2.09) |
| **Venous** | Composite venous | 0.91 (0.47, 1.74) | 0.90 (0.47, 1.73) | 0.69 (0.41, 1.18) | 0.69 (0.53, 0.90) | 0.63 (0.50, 0.79) | 1.05 (0.68, 1.64) | 0.90 (0.47, 1.73) | 0.90 (0.47, 1.73) | 0.69 (0.41, 1.17) | 0.68 (0.52, 0.89) | 0.62 (0.49, 0.78) | 1.03 (0.66, 1.60) |
| | Lower limb DVT[6] | 0.17 (0.02, 1.24) | 0.86 (0.36, 2.08) | 0.78 (0.40, 1.50) | 0.67 (0.47, 0.96) | 0.53 (0.38, 0.74) | 1.18 (0.67, 2.09) | 0.17 (0.02, 1.23) | 0.87 (0.36, 2.11) | 0.77 (0.40, 1.48) | 0.66 (0.47, 0.95) | 0.52 (0.37, 0.73) | 1.15 (0.65, 2.04) |
| | Pulmonary embolism | 1.69 (0.80, 3.57) | 0.96 (0.36, 2.57) | 0.59 (0.25, 1.43) | 0.73 (0.49, 1.07) | 0.73 (0.53, 1.00) | 0.89 (0.44, 1.80) | 1.62 (0.77, 3.42) | 0.91 (0.34, 2.43) | 0.57 (0.23, 1.36) | 0.69 (0.47, 1.02) | 0.70 (0.51, 0.96) | 0.84 (0.42, 1.70) |
| | ICVT[7]* | | | 1.40 (0.33, 5.99) | | | 0.59 (0.21, 1.67) | | | 1.50 (0.36, 6.23) | | | 0.57 (0.20, 1.63) |
| | Mesenteric thrombus* | | | 1.96 (0.47, 8.13) | | | 0.76 (0.28, 2.08) | | | 1.72 (0.42, 7.09) | | | 0.67 (0.24, 1.82) |
| | Portal vein thrombosis | | | | | | | | | | | | |
| **Other** | Any thrombocytopenia | | | 2.17 (1.01, 4.66) | | | 1.22 (0.75, 1.98) | | | 1.99 (0.92, 4.32) | | | 1.12 (0.69, 1.83) |
| | SAH and HS[8] | | | 0.80 (0.33, 1.95) | | | 0.80 (0.53, 1.20) | | | 0.83 (0.34, 2.03) | | | 0.82 (0.54, 1.25) |
| | Myocarditis | 5.57 (1.70,18.26) | 1.63 (0.22,11.86) | 0.87 (0.12, 6.42) | 5.64 (3.61, 8.81) | 1.95 (1.12, 3.40) | 2.09 (0.49, 8.88) | 4.64 (1.40,15.31) | 1.52 (0.21,10.99) | 0.87 (0.12, 6.45) | 5.35 (3.37, 8.48) | 1.78 (1.00, 3.16) | 2.05 (0.49, 8.66) |
| | Pericarditis | 1.75 (0.24,12.93) | 1.91 (0.26,13.99) | 2.07 (0.49, 8.79) | 1.97 (0.87, 4.44) | 1.83 (0.89, 3.79) | 2.34 (0.56, 9.84) | 2.01 (0.26,15.31) | 1.85 (0.25,13.87) | 2.11 (0.49, 9.06) | 1.94 (0.88, 4.29) | 1.95 (0.95, 4.01) | 2.16 (0.52, 9.01) |

*Time periods collapsed into weeks 0-4 and weeks 5-26.

Models could not be run for portal vein thrombosis.

Supplementary Table 11: Hazard ratios and 95% CIs for individual arterial and venous thrombotic events, and other cardiovascular events, by time since second dose vaccination for those on a primary course of ChAdOx1

| Outcome | | Age and Sex adjusted HR (95% CI) | | | | | | Fully Adjusted HR (95% CI) | | | | | |
|---|---|---|---|---|---|---|---|---|---|---|---|---|---|
| | | Week 1 | Week 2 | Weeks 3-4 | Weeks 5-12 | Weeks 13-24 | Weeks 25-26 | Week 1 | Week 2 | Weeks 3-4 | Weeks 5-12 | Weeks 13-24 | Weeks 25-26 |
| **Arterial** | Composite arterial | 0.73 (0.70,0.76) | 0.84 (0.81,0.88) | 0.81 (0.78,0.84) | 0.78 (0.76,0.81) | 0.78 (0.75,0.81) | 0.81 (0.77,0.85) | 0.72 (0.69,0.75) | 0.83 (0.80,0.87) | 0.79 (0.77,0.82) | 0.76 (0.74,0.78) | 0.73 (0.70,0.76) | 0.73 (0.70,0.77) |
| | AMI[2] | 0.76 (0.71,0.80) | 0.87 (0.82,0.92) | 0.86 (0.82,0.89) | 0.83 (0.80,0.87) | 0.84 (0.80,0.89) | 0.88 (0.82,0.95) | 0.73 (0.69,0.78) | 0.84 (0.79,0.89) | 0.82 (0.78,0.85) | 0.77 (0.74,0.80) | 0.73 (0.70,0.77) | 0.73 (0.68,0.79) |
| | Ischaemic stroke | 0.71 (0.67,0.75) | 0.82 (0.77,0.87) | 0.77 (0.74,0.81) | 0.75 (0.72,0.78) | 0.75 (0.70,0.79) | 0.76 (0.70,0.82) | 0.71 (0.67,0.76) | 0.83 (0.78,0.88) | 0.78 (0.75,0.82) | 0.76 (0.73,0.80) | 0.76 (0.72,0.80) | 0.77 (0.71,0.83) |
| **Venous** | Composite venous | 0.74 (0.67,0.80) | 0.86 (0.79,0.94) | 0.82 (0.77,0.88) | 0.75 (0.70,0.80) | 0.75 (0.69,0.81) | 0.80 (0.72,0.89) | 0.72 (0.66,0.79) | 0.84 (0.78,0.92) | 0.80 (0.75,0.85) | 0.71 (0.67,0.75) | 0.68 (0.63,0.73) | 0.69 (0.63,0.77) |
| | Lower limb DVT[6] | 0.72 (0.64,0.83) | 0.81 (0.72,0.92) | 0.80 (0.72,0.88) | 0.71 (0.64,0.77) | 0.68 (0.61,0.77) | 0.72 (0.61,0.84) | 0.71 (0.63,0.81) | 0.80 (0.70,0.90) | 0.77 (0.70,0.86) | 0.67 (0.61,0.73) | 0.62 (0.55,0.69) | 0.62 (0.54,0.73) |
| | Pulmonary embolism | 0.76 (0.67,0.85) | 0.90 (0.81,1.01) | 0.83 (0.76,0.91) | 0.78 (0.72,0.85) | 0.79 (0.71,0.89) | 0.85 (0.73,0.98) | 0.75 (0.66,0.84) | 0.88 (0.79,0.99) | 0.81 (0.74,0.89) | 0.74 (0.68,0.80) | 0.71 (0.64,0.79) | 0.73 (0.64,0.84) |
| | ICVT[7] | 0.30 (0.11,0.84) | 0.96 (0.51,1.80) | 0.93 (0.56,1.56) | 0.67 (0.40,1.12) | 0.74 (0.37,1.48) | 0.90 (0.33,2.44) | 0.29 (0.11,0.82) | 0.94 (0.50,1.76) | 0.88 (0.53,1.48) | 0.61 (0.36,1.04) | 0.63 (0.31,1.28) | 0.73 (0.27,1.96) |
| | Mesenteric thrombus | 0.64 (0.47,0.87) | 1.05 (0.81,1.36) | 0.77 (0.61,0.97) | 0.77 (0.62,0.96) | 0.76 (0.58,1.01) | 0.76 (0.52,1.09) | 0.62 (0.45,0.84) | 1.01 (0.78,1.31) | 0.73 (0.58,0.92) | 0.70 (0.57,0.87) | 0.64 (0.49,0.83) | 0.60 (0.42,0.85) |
| | Portal vein thrombosis | 0.66 (0.25,1.71) | 0.39 (0.11,1.37) | 0.92 (0.46,1.85) | 1.26 (0.67,2.38) | 1.44 (0.55,3.75) | 2.50 (0.70,8.87) | 0.49 (0.18,1.34) | 0.31 (0.09,1.09) | 0.67 (0.32,1.40) | 0.72 (0.36,1.43) | 0.52 (0.19,1.42) | 0.78 (0.22,2.74) |
| **Other** | Any thrombocytopenia | 0.89 (0.69,1.14) | 0.76 (0.57,1.01) | 0.95 (0.76,1.18) | 1.01 (0.81,1.25) | 1.43 (1.05,1.96) | 1.83 (1.18,2.82) | 0.73 (0.57,0.94) | 0.60 (0.46,0.79) | 0.71 (0.57,0.87) | 0.62 (0.52,0.75) | 0.66 (0.51,0.86) | 0.67 (0.46,0.96) |
| | SAH and HS[8] | 0.61 (0.51,0.72) | 0.76 (0.64,0.89) | 0.75 (0.66,0.85) | 0.68 (0.60,0.76) | 0.60 (0.52,0.69) | 0.59 (0.49,0.72) | 0.62 (0.52,0.74) | 0.77 (0.66,0.91) | 0.78 (0.68,0.88) | 0.71 (0.63,0.80) | 0.64 (0.56,0.74) | 0.64 (0.53,0.77) |
| | Myocarditis | 0.49 (0.23,1.07) | 0.75 (0.40,1.40) | 0.64 (0.38,1.07) | 0.75 (0.51,1.12) | 0.68 (0.40,1.13) | 0.73 (0.36,1.48) | 0.48 (0.22,1.04) | 0.75 (0.40,1.40) | 0.63 (0.38,1.05) | 0.72 (0.50,1.05) | 0.64 (0.40,1.02) | 0.67 (0.35,1.29) |
| | Pericarditis | 1.34 (0.84,2.16) | 0.80 (0.44,1.45) | 0.69 (0.43,1.10) | 0.50 (0.33,0.75) | 0.49 (0.29,0.81) | 0.59 (0.28,1.21) | 1.21 (0.76,1.95) | 0.68 (0.38,1.24) | 0.58 (0.36,0.91) | 0.39 (0.26,0.57) | 0.32 (0.19,0.51) | 0.33 (0.17,0.65) |

Supplementary Table 12: Hazard ratios and 95% CIs for individual arterial and venous thrombotic events, and other cardiovascular events, by time since second dose vaccination for those on a primary course of BNT-162b2.

| Outcome | | Age and Sex adjusted HR (95% CI) | | | | | | Fully Adjusted HR (95% CI) | | | | | |
|---|---|---|---|---|---|---|---|---|---|---|---|---|---|
| | | Week 1 | Week 2 | Weeks 3-4 | Weeks 5-12 | Weeks 13-24 | Weeks 25-26 | Week 1 | Week 2 | Weeks 3-4 | Weeks 5-12 | Weeks 13-24 | Weeks 25-26 |
| **Arterial** | Composite arterial | 0.71 (0.68,0.75) | 0.80 (0.77,0.84) | 0.81 (0.78,0.84) | 0.80 (0.78,0.83) | 0.80 (0.77,0.83) | 0.80 (0.76,0.84) | 0.73 (0.69,0.76) | 0.82 (0.78,0.85) | 0.82 (0.79,0.85) | 0.81 (0.78,0.83) | 0.80 (0.77,0.83) | 0.80 (0.76,0.85) |
| | AMI[2] | 0.70 (0.65,0.75) | 0.85 (0.79,0.90) | 0.83 (0.79,0.88) | 0.82 (0.79,0.86) | 0.85 (0.81,0.89) | 0.86 (0.80,0.93) | 0.71 (0.66,0.76) | 0.85 (0.80,0.91) | 0.84 (0.79,0.88) | 0.81 (0.78,0.85) | 0.82 (0.78,0.87) | 0.83 (0.77,0.90) |
| | Ischaemic stroke | 0.73 (0.68,0.78) | 0.76 (0.71,0.81) | 0.78 (0.74,0.82) | 0.79 (0.75,0.82) | 0.76 (0.72,0.80) | 0.75 (0.70,0.81) | 0.74 (0.69,0.79) | 0.77 (0.72,0.82) | 0.79 (0.75,0.83) | 0.80 (0.76,0.84) | 0.77 (0.72,0.81) | 0.76 (0.71,0.83) |
| **Venous** | Composite venous | 0.73 (0.66,0.81) | 0.82 (0.74,0.91) | 0.77 (0.71,0.84) | 0.79 (0.74,0.84) | 0.83 (0.77,0.89) | 0.82 (0.74,0.91) | 0.73 (0.66,0.81) | 0.82 (0.74,0.90) | 0.77 (0.71,0.83) | 0.76 (0.72,0.81) | 0.77 (0.72,0.83) | 0.74 (0.67,0.83) |
| | Lower limb DVT[6] | 0.74 (0.63,0.87) | 0.84 (0.72,0.98) | 0.83 (0.73,0.94) | 0.81 (0.74,0.89) | 0.82 (0.73,0.91) | 0.77 (0.65,0.91) | 0.75 (0.63,0.88) | 0.85 (0.73,0.99) | 0.84 (0.74,0.95) | 0.82 (0.74,0.90) | 0.81 (0.73,0.91) | 0.77 (0.65,0.91) |
| | Pulmonary embolism | 0.72 (0.63,0.83) | 0.79 (0.69,0.91) | 0.74 (0.66,0.82) | 0.76 (0.70,0.83) | 0.83 (0.75,0.92) | 0.84 (0.73,0.97) | 0.73 (0.63,0.84) | 0.80 (0.69,0.91) | 0.74 (0.66,0.82) | 0.74 (0.68,0.81) | 0.79 (0.71,0.87) | 0.78 (0.68,0.90) |
| | ICVT[7] | 0.47 (0.17,1.30) | 1.16 (0.55,2.44) | 0.71 (0.36,1.37) | 0.74 (0.47,1.16) | 0.88 (0.53,1.47) | 0.34 (0.11,1.06) | 0.51 (0.18,1.43) | 1.28 (0.61,2.70) | 0.76 (0.38,1.50) | 0.77 (0.49,1.21) | 0.87 (0.52,1.45) | 0.32 (0.10,0.99) |
| | Mesenteric thrombus | 0.55 (0.38,0.80) | 0.71 (0.52,0.99) | 0.86 (0.67,1.11) | 0.66 (0.53,0.83) | 0.71 (0.54,0.92) | 0.70 (0.49,1.02) | 0.54 (0.38,0.79) | 0.70 (0.51,0.97) | 0.84 (0.65,1.08) | 0.63 (0.50,0.79) | 0.64 (0.49,0.83) | 0.63 (0.43,0.91) |
| | Portal vein thrombosis | 0.68 (0.23,2.01) | 0.33 (0.07,1.49) | 0.64 (0.23,1.76) | 0.93 (0.48,1.81) | 0.89 (0.41,1.91) | 0.70 (0.22,2.22) | 0.57 (0.17,1.97) | 0.28 (0.06,1.28) | 0.61 (0.21,1.81) | 0.79 (0.36,1.72) | 0.60 (0.22,1.70) | 0.48 (0.11,2.06) |
| **Other** | Any thrombocytopenia | 0.84 (0.62,1.13) | 0.77 (0.56,1.05) | 1.00 (0.80,1.25) | 0.89 (0.74,1.06) | 1.18 (0.95,1.47) | 1.39 (0.99,1.96) | 0.78 (0.58,1.06) | 0.72 (0.52,0.98) | 0.92 (0.73,1.16) | 0.74 (0.62,0.89) | 0.87 (0.70,1.09) | 0.94 (0.67,1.33) |
| | SAH and HS[8] | 0.69 (0.56,0.85) | 0.83 (0.68,1.00) | 0.95 (0.82,1.10) | 0.86 (0.76,0.98) | 0.81 (0.69,0.94) | 0.73 (0.59,0.91) | 0.70 (0.57,0.86) | 0.85 (0.70,1.02) | 0.97 (0.84,1.13) | 0.89 (0.78,1.01) | 0.84 (0.73,0.98) | 0.77 (0.62,0.96) |
| | Myocarditis | 3.21 (2.10,4.92) | 1.60 (0.93,2.77) | 0.97 (0.58,1.61) | 1.07 (0.77,1.49) | 0.92 (0.65,1.31) | 1.32 (0.71,2.43) | 3.14 (2.04,4.85) | 1.63 (0.94,2.82) | 0.98 (0.59,1.63) | 1.11 (0.79,1.56) | 0.95 (0.66,1.38) | 1.42 (0.76,2.65) |
| | Pericarditis | 0.91 (0.43,1.90) | 1.44 (0.80,2.61) | 2.37 (1.59,3.54) | 1.38 (0.98,1.93) | 1.18 (0.79,1.77) | 1.65 (0.84,3.23) | 0.86 (0.41,1.81) | 1.45 (0.80,2.63) | 2.42 (1.62,3.62) | 1.34 (0.96,1.88) | 1.16 (0.77,1.72) | 1.61 (0.83,3.13) |

Supplementary Table 13: Hazard ratios and 95% CIs for individual arterial and venous thrombotic events, and other cardiovascular events, by time since second dose vaccination for those on a primary course of mRNA-1273.

| Outcome | | Age and Sex adjusted HR (95% CI) | | | | | | Fully Adjusted HR (95% CI) | | | | | |
|---|---|---|---|---|---|---|---|---|---|---|---|---|---|
| | | Week 1 | Week 2 | Weeks 3-4 | Weeks 5-12 | Weeks 13-24 | Weeks 25-26 | Week 1 | Week 2 | Weeks 3-4 | Weeks 5-12 | Weeks 13-24 | Weeks 25-26 |
| **Arterial** | Composite arterial | 0.56 (0.26, 1.21) | 1.38 (0.78, 2.42) | 0.47 (0.24, 0.91) | 0.64 (0.43, 0.94) | 0.74 (0.47, 1.17) | 0.35 (0.14, 0.92) | 0.78 (0.36, 1.67) | 1.60 (0.90, 2.85) | 0.62 (0.32, 1.23) | 0.81 (0.52, 1.27) | 1.09 (0.64, 1.83) | 0.59 (0.22, 1.62) |
| | AMI[2] | 0.98 (0.38, 2.54) | 2.08 (1.02, 4.25) | 0.45 (0.17, 1.19) | 0.69 (0.37, 1.29) | 0.52 (0.27, 0.98) | 0.36 (0.11, 1.16) | 1.05 (0.40, 2.75) | 2.39 (1.11, 5.17) | 0.59 (0.23, 1.51) | 0.86 (0.44, 1.69) | 0.75 (0.34, 1.66) | 0.53 (0.14, 2.07) |
| | Ischaemic stroke | 0.22 (0.05, 1.01) | 0.76 (0.25, 2.34) | 0.50 (0.18, 1.39) | 0.66 (0.35, 1.24) | 1.01 (0.49, 2.11) | 0.12 (0.01, 0.97) | 0.39 (0.08, 1.89) | 1.13 (0.38, 3.35) | 0.56 (0.19, 1.67) | 0.69 (0.35, 1.34) | 1.11 (0.55, 2.22) | 0.13 (0.02, 0.83) |
| **Venous** | Composite venous | 0.65 (0.28, 1.52) | 0.21 (0.05, 0.88) | 0.81 (0.44, 1.49) | 0.61 (0.39, 0.95) | 0.57 (0.36, 0.90) | 0.25 (0.06, 1.05) | 0.74 (0.31, 1.77) | 0.24 (0.06, 0.99) | 0.85 (0.45, 1.59) | 0.72 (0.45, 1.16) | 0.81 (0.46, 1.45) | 0.41 (0.08, 2.00) |
| | Lower limb DVT[6] | 0.66 (0.23, 1.89) | 0.16 (0.02, 1.23) | 0.46 (0.18, 1.20) | 0.38 (0.20, 0.71) | 0.40 (0.20, 0.77) | 0.20 (0.02, 1.67) | 0.95 (0.32, 2.78) | 0.20 (0.03, 1.56) | 0.56 (0.20, 1.53) | 0.49 (0.24, 0.99) | 0.52 (0.25, 1.08) | 0.30 (0.04, 2.38) |
| | Pulmonary embolism | 0.64 (0.15, 2.82) | 0.33 (0.04, 2.52) | 1.48 (0.64, 3.43) | 0.94 (0.47, 1.88) | 0.80 (0.39, 1.63) | 0.32 (0.04, 2.72) | 0.65 (0.14, 3.02) | 0.27 (0.03, 2.45) | 1.54 (0.68, 3.51) | 1.01 (0.50, 2.07) | 0.96 (0.46, 1.97) | 0.36 (0.04, 3.01) |
| | ICVT[7] | | | | | | | | | | | | |
| | Mesenteric thrombus | | | | | | | | | | | | |
| | Portal vein thrombosis | | | | | | | | | | | | |
| **Other** | Any thrombocytopenia* | | | 1.17 (0.31, 4.37) | | | 0.92 (0.15, 5.45) | | | 1.08 (0.24, 4.84) | | | 0.91 (0.11, 7.52) |
| | SAH and HS[8]* | | | 0.36 (0.06, 2.28) | | | 1.02 (0.31, 3.33) | | | 0.25 (0.04, 1.63) | | | 0.70 (0.24, 2.10) |
| | Myocarditis* | | | 7.62 (2.82,20.58) | | | 0.47 (0.14, 1.50) | | | 10.8 (3.79, 30.83) | | | 0.65 (0.20, 2.11) |
| | Pericarditis* | | | 1.93 (0.58, 6.44) | | | 1.06 (0.27, 4.11) | | | 1.40 (0.30, 6.64) | | | 0.54 (0.10, 2.98) |

*Time periods collapsed into weeks 0-4 and weeks 5-26.
Models could not be run for ICVT, mesenteric thrombus, portal vein thrombosis.

Supplementary Table 14: Hazard ratios and 95% CIs for individual arterial and venous thrombotic events, and other cardiovascular events, by time since booster vaccination with mRNA-1273 for those on a primary course of ChAdOx1.

| **Outcome** | | **Age and Sex adjusted HR (95% CI)** | | | | | **Fully Adjusted HR (95% CI)** | | | | |
|---|---|---|---|---|---|---|---|---|---|---|---|
| | | Week 1 | Week 2 | Weeks 3-4 | Weeks 5-12 | Weeks 13-24 | Week 1 | Week 2 | Weeks 3-4 | Weeks 5-12 | Weeks 13-24 |
| **Arterial** | Composite arterial* | | 0.64 (0.60,0.68) | | 0.53 (0.49,0.57) | | | 0.77 (0.73, 0.82) | | 0.67 (0.62, 0.72) | |
| | AMI[2]* | | 0.67 (0.62,0.73) | | 0.58 (0.52,0.64) | | | 0.79 (0.73, 0.86) | | 0.71 (0.64, 0.79) | |
| | Ischaemic stroke* | | 0.60 (0.54,0.66) | | 0.45 (0.40,0.51) | | | 0.75 (0.68, 0.82) | | 0.61 (0.53, 0.69) | |
| **Venous** | Composite venous* | | 0.54 (0.48,0.61) | | 0.50 (0.43,0.59) | | | 0.58 (0.51, 0.66) | | 0.55 (0.47, 0.65) | |
| | Lower limb DVT[6]* | | 0.60 (0.50,0.72) | | 0.54 (0.42,0.68) | | | 0.64 (0.53, 0.77) | | 0.58 (0.46, 0.74) | |
| | Pulmonary embolism* | | 0.48 (0.40,0.57) | | 0.48 (0.39,0.59) | | | 0.52 (0.44, 0.62) | | 0.54 (0.44, 0.66) | |
| | ICVT[7]* | | 0.86 (0.32,2.30) | | 0.82 (0.25,2.72) | | | 0.92 (0.35, 2.43) | | 0.92 (0.29, 2.93) | |
| | Mesenteric thrombus* | | 0.45 (0.28,0.71) | | 0.36 (0.19,0.66) | | | 0.52 (0.33, 0.82) | | 0.41 (0.22, 0.77) | |
| | Portal vein thrombosis | | | | | | | | | | |
| **Other** | Any thrombocytopenia* | | 0.79 (0.49,1.27) | | 0.56 (0.27,1.18) | | | 0.90 (0.56, 1.44) | | 0.63 (0.30, 1.32) | |
| | SAH and HS[8] | 0.31 (0.17,0.58) | 0.53 (0.33,0.86) | 0.56 (0.39,0.81) | 0.52 (0.36,0.74) | NA (NA, NA) | | | | | |
| | Myocarditis* | | 0.67 (0.31,1.46) | | 0.72 (0.24,2.16) | | | 0.72 (0.33, 1.56) | | 0.79 (0.26, 2.36) | |
| | Pericarditis* | | 0.63 (0.24,1.60) | | 0.12 (0.01,0.95) | | | 0.63 (0.25, 1.62) | | 0.12 (0.01, 0.98) | |

*Time periods collapsed into weeks 0-4 and weeks 5-24.
Models could not be run for portal vein thrombosis and part of SAH and HS.
NA (NA-NA) Could not obtain estimate for this time interval

Supplementary Table 15: Hazard ratios and 95% CIs for individual arterial and venous thrombotic events, and other cardiovascular events, by time since booster vaccination with BNT-162b2 for those on a primary course of ChAdOx1.

| Outcome | | Age and Sex adjusted HR (95% CI) | | | | | Fully Adjusted HR (95% CI) | | | | |
|---|---|---|---|---|---|---|---|---|---|---|---|
| | | Week 1 | Week 2 | Weeks 3-4 | Weeks 5-12 | Weeks 13-24 | Weeks 1 | Week 2 | Week 3-4 | Weeks 5-12 | Weeks 13-24 |
| **Arterial** | Composite arterial | 0.77 (0.73,0.80) | 0.79 (0.75,0.83) | 0.79 (0.76,0.82) | 0.73 (0.70,0.75) | 0.64 (0.60,0.69) | 0.79 (0.75, 0.83) | 0.82 (0.78, 0.86) | 0.83 (0.80, 0.86) | 0.78 (0.75, 0.81) | 0.71 (0.66, 0.76) |
| | AMI[2] | 0.76 (0.71,0.81) | 0.81 (0.76,0.87) | 0.79 (0.75,0.83) | 0.73 (0.70,0.77) | 0.62 (0.55,0.69) | 0.76 (0.71, 0.82) | 0.82 (0.76, 0.87) | 0.80 (0.76, 0.84) | 0.75 (0.72, 0.79) | 0.63 (0.57, 0.71) |
| | Ischaemic stroke | 0.78 (0.73,0.84) | 0.78 (0.73,0.84) | 0.80 (0.76,0.85) | 0.74 (0.70,0.78) | 0.69 (0.62,0.76) | 0.82 (0.76, 0.88) | 0.83 (0.77, 0.89) | 0.86 (0.82, 0.91) | 0.84 (0.80, 0.88) | 0.81 (0.73, 0.90) |
| **Venous** | Composite venous | 0.69 (0.62,0.77) | 0.77 (0.69,0.85) | 0.75 (0.69,0.81) | 0.68 (0.63,0.73) | 0.68 (0.58,0.80) | 0.67 (0.60, 0.75) | 0.74 (0.67, 0.83) | 0.73 (0.67, 0.79) | 0.64 (0.60, 0.69) | 0.63 (0.54, 0.74) |
| | Lower limb DVT[6] | 0.73 (0.61,0.86) | 0.90 (0.77,1.05) | 0.83 (0.74,0.94) | 0.69 (0.62,0.78) | 0.84 (0.66,1.07) | 0.71 (0.60, 0.84) | 0.87 (0.75, 1.02) | 0.81 (0.71, 0.91) | 0.66 (0.59, 0.74) | 0.76 (0.60, 0.97) |
| | Pulmonary embolism | 0.65 (0.57,0.75) | 0.68 (0.59,0.78) | 0.68 (0.61,0.76) | 0.64 (0.58,0.70) | 0.56 (0.46,0.70) | 0.64 (0.55, 0.73) | 0.66 (0.58, 0.76) | 0.66 (0.60, 0.74) | 0.61 (0.56, 0.67) | 0.53 (0.43, 0.65) |
| | ICVT[7]* | | 0.75 (0.38,1.46) | | 0.86 (0.38,1.98) | | | 0.72 (0.37,1.40) | | 0.76 (0.35,1.68) | |
| | Mesenteric thrombus | 0.56 (0.38,0.81) | 0.63 (0.44,0.90) | 0.53 (0.39,0.71) | 0.60 (0.47,0.76) | 0.75 (0.48,1.17) | 0.56 (0.38, 0.82) | 0.64 (0.45, 0.92) | 0.55 (0.41, 0.74) | 0.65 (0.51, 0.82) | 0.84 (0.53, 1.31) |
| | Portal vein thrombosis | 0.68 (0.20,2.30) | 0.22 (0.03,1.69) | 0.57 (0.21,1.53) | 0.69 (0.32,1.50) | 0.79 (0.14,4.41) | 0.64 (0.18, 2.32) | 0.16 (0.02, 1.42) | 0.42 (0.14, 1.26) | 0.50 (0.21, 1.18) | 0.58 (0.08, 4.19) |
| **Other** | Any thrombocytopenia | 1.49 (1.08,2.06) | 1.42 (1.01,2.00) | 1.47 (1.11,1.94) | 1.56 (1.19,2.05) | 3.76 (2.19,6.47) | 1.26 (0.92, 1.73) | 1.17 (0.83, 1.64) | 1.16 (0.88, 1.53) | 1.09 (0.84, 1.41) | 2.16 (1.26, 3.69) |
| | SAH and HS[8] | 0.65 (0.53,0.81) | 0.71 (0.58,0.87) | 0.66 (0.57,0.78) | 0.73 (0.64,0.83) | 0.73 (0.55,0.98) | 0.68 (0.55, 0.84) | 0.74 (0.60, 0.91) | 0.70 (0.60, 0.82) | 0.80 (0.70, 0.91) | 0.82 (0.61, 1.09) |
| | Myocarditis | 1.42 (0.74,2.73) | 1.05 (0.49,2.24) | 0.97 (0.53,1.79) | 0.75 (0.43,1.33) | 0.38 (0.05,3.04) | 1.32 (0.68, 2.56) | 0.99 (0.46, 2.11) | 0.90 (0.49, 1.65) | 0.65 (0.37, 1.15) | 0.30 (0.04, 2.36) |
| | Pericarditis | 0.56 (0.20,1.53) | 1.39 (0.69,2.81) | 0.41 (0.17,0.96) | 1.01 (0.58,1.76) | 0.35 (0.04,2.81) | 0.52 (0.19, 1.43) | 1.26 (0.62, 2.55) | 0.37 (0.16, 0.87) | 0.82 (0.47, 1.42) | 0.25 (0.03, 1.96) |

*Time periods collapsed into weeks 0-4 and weeks 5-24.

Supplementary Table 16: Hazard ratios and 95% CIs for individual arterial and venous thrombotic events, and other cardiovascular events, by time since booster vaccination with mRNA-1273 or BNT-162b2 for those on a primary course of ChAdOx1.

| **Outcome** | | **Age and Sex adjusted HR (95% CI)** | | | | | **Fully Adjusted HR (95% CI)** | | | | |
|---|---|---|---|---|---|---|---|---|---|---|---|
| | | Week 1 | Week 2 | Weeks 3-4 | Weeks 5-12 | Weeks 13-24 | Week 1 | Week 2 | Weeks 3-4 | Weeks 5-12 | Weeks 13-24 |
| **Arterial** | Composite arterial | 0.75 (0.72,0.79) | 0.78 (0.74,0.82) | 0.78 (0.75,0.81) | 0.73 (0.70,0.75) | 0.66 (0.62,0.71) | 0.79 (0.75,0.83) | 0.82 (0.79,0.86) | 0.83 (0.80,0.86) | 0.78 (0.76,0.81) | 0.73 (0.67,0.78) |
| | AMI[2] | 0.74 (0.70,0.79) | 0.78 (0.74,0.84) | 0.78 (0.74,0.82) | 0.73 (0.70,0.76) | 0.62 (0.56,0.69) | 0.77 (0.72,0.82) | 0.81 (0.76,0.86) | 0.81 (0.77,0.85) | 0.75 (0.72,0.79) | 0.64 (0.58,0.71) |
| | Ischaemic stroke | 0.77 (0.72,0.82) | 0.77 (0.72,0.83) | 0.78 (0.74,0.82) | 0.73 (0.70,0.77) | 0.71 (0.64,0.79) | 0.82 (0.76,0.87) | 0.83 (0.78,0.89) | 0.84 (0.80,0.89) | 0.82 (0.78,0.86) | 0.81 (0.73,0.90) |
| **Venous** | Composite venous | 0.66 (0.60,0.73) | 0.76 (0.69,0.83) | 0.71 (0.66,0.77) | 0.67 (0.63,0.72) | 0.67 (0.58,0.79) | 0.66 (0.59,0.72) | 0.75 (0.68,0.82) | 0.70 (0.65,0.76) | 0.64 (0.60,0.69) | 0.63 (0.54,0.74) |
| | Lower limb DVT[6] | 0.69 (0.59,0.80) | 0.89 (0.77,1.03) | 0.81 (0.72,0.92) | 0.71 (0.64,0.80) | 0.89 (0.70,1.13) | 0.68 (0.58,0.80) | 0.89 (0.77,1.03) | 0.81 (0.72,0.91) | 0.70 (0.63,0.78) | 0.84 (0.66,1.06) |
| | Pulmonary embolism | 0.63 (0.55,0.72) | 0.66 (0.58,0.75) | 0.64 (0.58,0.71) | 0.64 (0.58,0.70) | 0.57 (0.47,0.71) | 0.63 (0.55,0.71) | 0.66 (0.58,0.74) | 0.64 (0.57,0.70) | 0.62 (0.56,0.68) | 0.55 (0.44,0.67) |
| | ICVT[7]* | | 0.78 (0.43,1.42) | | | 0.83 (0.39,1.78) | | | 0.76 (0.41,1.38) | | 0.78 (0.36,1.68) |
| | Mesenteric thrombus | 0.51 (0.35,0.74) | 0.55 (0.38,0.79) | 0.58 (0.44,0.75) | 0.59 (0.46,0.75) | 0.83 (0.53,1.31) | 0.52 (0.36,0.75) | 0.56 (0.39,0.80) | 0.60 (0.46,0.79) | 0.61 (0.48,0.77) | 0.85 (0.54,1.34) |
| | Portal vein thrombosis | 0.62 (0.21,1.80) | 0.45 (0.14,1.48) | 0.58 (0.26,1.31) | 0.44 (0.20,0.98) | 0.45 (0.09,2.34) | 0.61 (0.20,1.91) | 0.45 (0.13,1.59) | 0.68 (0.28,1.63) | 0.42 (0.19,0.94) | 0.27 (0.04,1.76) |
| **Other** | Any thrombocytopenia | 1.33 (0.97,1.81) | 1.30 (0.94,1.81) | 1.44 (1.10,1.88) | 1.53 (1.17,2.00) | 3.87 (2.27,6.59) | 1.14 (0.84,1.55) | 1.09 (0.79,1.50) | 1.14 (0.88,1.48) | 1.05 (0.81,1.36) | 2.17 (1.27,3.71) |
| | SAH and HS[8] | 0.59 (0.49,0.73) | 0.68 (0.56,0.82) | 0.65 (0.56,0.76) | 0.73 (0.64,0.83) | 0.80 (0.60,1.07) | 0.64 (0.52,0.78) | 0.73 (0.61,0.89) | 0.71 (0.61,0.83) | 0.82 (0.72,0.94) | 0.93 (0.70,1.24) |
| | Myocarditis | 1.38 (0.77,2.46) | 1.04 (0.53,2.04) | 0.83 (0.45,1.52) | 0.80 (0.46,1.39) | 0.47 (0.06,3.75) | 1.34 (0.76,2.37) | 1.01 (0.52,1.97) | 0.78 (0.43,1.43) | 0.70 (0.41,1.21) | 0.39 (0.05,3.07) |
| | Pericarditis | 0.41 (0.15,1.13) | 1.20 (0.61,2.36) | 0.59 (0.30,1.14) | 0.88 (0.50,1.55) | 0.33 (0.04,2.63) | 0.40 (0.14,1.10) | 1.17 (0.60,2.28) | 0.55 (0.29,1.07) | 0.79 (0.46,1.37) | 0.26 (0.03,1.98) |

*Time periods collapsed into weeks 0-4 and weeks 5-24.

Supplementary Table 17: Hazard ratios and 95% CIs for individual arterial and venous thrombotic events, and other cardiovascular events, by time since booster vaccination with BNT-162b2 for those on a primary course of BNT-162b2.

| | | Age and Sex adjusted HR (95% CI) | | | | | Fully Adjusted HR (95% CI) | | | | |
|---|---|---|---|---|---|---|---|---|---|---|---|
| **Outcome** | | Week 1 | Week 2 | Weeks 3-4 | Weeks 5-12 | Weeks 13-24 | Week 1 | Week 2 | Weeks 3-4 | Weeks 5-12 | Weeks 13-24 |
| **Arterial** | Composite arterial | 0.66 (0.63, 0.70) | 0.74 (0.71, 0.78) | 0.75 (0.72, 0.78) | 0.70 (0.68, 0.73) | 0.63 (0.60, 0.67) | 0.69 (0.65, 0.73) | 0.78 (0.74, 0.83) | 0.80 (0.77, 0.83) | 0.78 (0.75, 0.81) | 0.73 (0.69, 0.77) |
| | AMI[2] | 0.66 (0.61, 0.71) | 0.76 (0.70, 0.81) | 0.79 (0.75, 0.84) | 0.72 (0.68, 0.75) | 0.63 (0.58, 0.68) | 0.68 (0.63, 0.74) | 0.79 (0.73, 0.85) | 0.84 (0.79, 0.89) | 0.78 (0.75, 0.83) | 0.71 (0.66, 0.77) |
| | Ischaemic stroke | 0.67 (0.62, 0.72) | 0.74 (0.69, 0.79) | 0.71 (0.67, 0.75) | 0.70 (0.66, 0.73) | 0.65 (0.60, 0.70) | 0.71 (0.65, 0.76) | 0.79 (0.73, 0.85) | 0.78 (0.73, 0.82) | 0.80 (0.76, 0.84) | 0.78 (0.72, 0.85) |
| **Venous** | Composite venous | 0.63 (0.56, 0.71) | 0.66 (0.59, 0.74) | 0.64 (0.59, 0.70) | 0.61 (0.56, 0.65) | 0.54 (0.48, 0.61) | 0.63 (0.56, 0.71) | 0.66 (0.59, 0.74) | 0.65 (0.59, 0.71) | 0.62 (0.57, 0.66) | 0.56 (0.49, 0.63) |
| | Lower limb DVT[6] | 0.71 (0.59, 0.86) | 0.77 (0.64, 0.91) | 0.68 (0.59, 0.78) | 0.63 (0.56, 0.71) | 0.54 (0.44, 0.66) | 0.71 (0.59, 0.86) | 0.76 (0.64, 0.91) | 0.68 (0.59, 0.78) | 0.63 (0.56, 0.71) | 0.55 (0.45, 0.67) |
| | Pulmonary embolism | 0.56 (0.48, 0.66) | 0.59 (0.51, 0.69) | 0.62 (0.55, 0.70) | 0.58 (0.53, 0.64) | 0.54 (0.46, 0.63) | 0.56 (0.48, 0.66) | 0.59 (0.51, 0.69) | 0.62 (0.55, 0.70) | 0.59 (0.53, 0.65) | 0.55 (0.47, 0.65) |
| | ICVT[7]* | 0.50 (0.12, 2.11) | 0.56 (0.13, 2.34) | 0.74 (0.31, 1.78) | 0.76 (0.41, 1.40) | 0.83 (0.32, 2.13) | 0.45 (0.11, 1.89) | 0.54 (0.13, 2.30) | 0.68 (0.28, 1.66) | 0.70 (0.37, 1.31) | 0.76 (0.29, 1.99) |
| | Mesenteric thrombus | 0.48 (0.33, 0.71) | 0.45 (0.30, 0.67) | 0.66 (0.51, 0.85) | 0.54 (0.43, 0.68) | 0.41 (0.28, 0.60) | 0.49 (0.33, 0.73) | 0.46 (0.31, 0.68) | 0.68 (0.53, 0.88) | 0.56 (0.45, 0.71) | 0.43 (0.29, 0.64) |
| | Portal vein thrombosis | | 0.25 (0.09, 0.72) | | 0.51 (0.23, 1.14) | | | 0.27 (0.09, 0.80) | | 0.48 (0.19, 1.20) | |
| **Other** | Any thrombocytopenia | 0.84 (0.57, 1.24) | 0.56 (0.34, 0.91) | 1.12 (0.84, 1.49) | 0.86 (0.67, 1.11) | 1.20 (0.80, 1.81) | 0.80 (0.54, 1.18) | 0.52 (0.32, 0.85) | 1.04 (0.78, 1.37) | 0.77 (0.60, 0.99) | 1.06 (0.71, 1.60) |
| | SAH and HS[8] | 0.71 (0.58, 0.88) | 0.83 (0.68, 1.02) | 0.85 (0.73, 0.99) | 0.73 (0.63, 0.84) | 0.63 (0.49, 0.79) | 0.74 (0.60, 0.91) | 0.87 (0.72, 1.07) | 0.90 (0.77, 1.05) | 0.80 (0.69, 0.92) | 0.71 (0.56, 0.91) |
| | Myocarditis | 1.83 (1.01, 3.32) | 1.05 (0.49, 2.25) | 1.24 (0.69, 2.23) | 0.88 (0.53, 1.48) | 0.34 (0.08, 1.50) | 1.79 (0.98, 3.25) | 1.03 (0.48, 2.22) | 1.23 (0.68, 2.21) | 0.86 (0.51, 1.44) | 0.34 (0.08, 1.51) |
| | Pericarditis | 0.98 (0.39, 2.47) | 1.99 (0.97, 4.10) | 1.17 (0.59, 2.31) | 1.32 (0.78, 2.24) | 1.33 (0.42, 4.23) | 0.97 (0.38, 2.44) | 1.99 (0.96, 4.09) | 1.16 (0.58, 2.31) | 1.30 (0.76, 2.21) | 1.30 (0.41, 4.18) |

*Time periods collapsed into weeks 0-4 and weeks 5-24.

Supplementary Table 18: Hazard ratios and 95% CIs for individual arterial and venous thrombotic events, and other cardiovascular events, by time since booster vaccination with mRNA-1273 for those on a primary course of BNT-162b2.

| | | Age and Sex adjusted HR (95% CI) | | | | | Fully Adjusted HR (95% CI) | | | | |
|---|---|---|---|---|---|---|---|---|---|---|---|
| **Outcome** | | Week 1 | Week 2 | Weeks 3-4 | Weeks 5-12 | Weeks 13-24 | Week 1 | Week 2 | Weeks 3-4 | Weeks 5-12 | Weeks 13-24 |
| **Arterial** | Composite arterial | 0.63 (0.52, 0.78) | 0.91 (0.77, 1.09) | 0.82 (0.71, 0.94) | 0.77 (0.69, 0.87) | 0.88 (0.28, 2.79) | 0.67 (0.55, 0.82) | 0.97 (0.81, 1.15) | 0.88 (0.77, 1.01) | 0.85 (0.75, 0.96) | 1.21 (0.38, 3.86) |
| | AMI[2] | 0.86 (0.75, 0.98) | | | 0.80 (0.68, 0.94) | | 0.90 (0.79, 1.04) | | | 0.86 (0.73, 1.02) | |
| | Ischaemic stroke | 0.46 (0.33, 0.65) | 0.90 (0.70, 1.16) | 0.75 (0.61, 0.93) | 0.70 (0.59, 0.84) | NA (NA, NA) | 0.48 (0.34, 0.68) | 0.95 (0.74, 1.22) | 0.80 (0.65, 0.98) | 0.77 (0.64, 0.93) | 2.29 (0.67, 7.82) |
| **Venous** | Composite venous | 0.60 (0.49, 0.74) | | | 0.56 (0.44, 0.72) | | 0.62 (0.50, 0.76) | | | 0.58 (0.45, 0.74) | |
| | Lower limb DVT[6] | 0.53 (0.39, 0.74) | | | 0.48 (0.31, 0.72) | | 0.52 (0.38, 0.73) | | | 0.46 (0.30, 0.69) | |
| | Pulmonary embolism | 0.66 (0.50, 0.87) | | | 0.67 (0.49, 0.92) | | 0.66 (0.50, 0.87) | | | 0.67 (0.49, 0.92) | |
| | ICVT[7]* | 0.28 (0.04, 1.86) | | | 0.40 (0.05, 3.22) | | 0.28 (0.04, 1.87) | | | 0.40 (0.05, 3.20) | |
| | Mesenteric thrombus | 0.69 (0.36, 1.33) | | | 0.68 (0.31, 1.45) | | 0.69 (0.35, 1.33) | | | 0.66 (0.30, 1.44) | |
| | Portal vein thrombosis | 0.65 (0.08, 5.17) | | | 0.81 (0.08, 8.10) | | 0.66 (0.08, 5.19) | | | 0.82 (0.08, 7.94) | |
| **Other** | Any thrombocytopenia | 1.04 (0.59, 1.84) | | | 0.13 (0.02, 0.98) | | 0.98 (0.56, 1.72) | | | 0.11 (0.01, 0.80) | |
| | SAH and HS[8] | 0.71 (0.46, 1.09) | | | 0.64 (0.36, 1.14) | | 0.76 (0.49, 1.18) | | | 0.70 (0.40, 1.25) | |
| | Myocarditis | 0.92 (0.36, 2.34) | | | 0.55 (0.08, 4.00) | | 0.87 (0.34, 2.21) | | | 0.52 (0.07, 3.77) | |
| | Pericarditis | 2.23 (0.71, 7.00) | | | 1.06 (0.09, 12.46) | | 2.33 (0.74, 7.35) | | | 1.08 (0.09, 12.70) | |

*Time periods collapsed into weeks 0-4 and weeks 5-24.

Supplementary Table 19: Hazard ratios and 95% CIs for individual arterial and venous thrombotic events, and other cardiovascular events, by time since booster vaccination with mRNA-1273 or BNT-162b2 for those on a primary course of BNT-162b2.

| | | Age and Sex adjusted HR (95% CI) | | | | | Fully Adjusted HR (95% CI) | | | | |
|---|---|---|---|---|---|---|---|---|---|---|---|
| **Outcome** | | Week 1 | Week 2 | Weeks 3-4 | Weeks 5-12 | Weeks 13-24 | Week 1 | Week 2 | Weeks 3-4 | Weeks 5-12 | Weeks 13-24 |
| **Arterial** | Composite arterial | 0.66 (0.62, 0.69) | 0.75 (0.71, 0.79) | 0.75 (0.72, 0.78) | 0.70 (0.68, 0.73) | 0.63 (0.59, 0.66) | 0.69 (0.65, 0.73) | 0.79 (0.75, 0.83) | 0.80 (0.77, 0.84) | 0.78 (0.75, 0.81) | 0.73 (0.68, 0.77) |
| | AMI[2] | 0.66 (0.62, 0.72) | 0.76 (0.71, 0.82) | 0.79 (0.75, 0.84) | 0.71 (0.68, 0.75) | 0.62 (0.58, 0.68) | 0.69 (0.64, 0.74) | 0.79 (0.74, 0.85) | 0.83 (0.79, 0.88) | 0.77 (0.73, 0.81) | 0.70 (0.64, 0.76) |
| | Ischaemic stroke | 0.65 (0.61, 0.70) | 0.75 (0.69, 0.80) | 0.71 (0.67, 0.75) | 0.69 (0.66, 0.73) | 0.65 (0.60, 0.70) | 0.69 (0.64, 0.74) | 0.80 (0.74, 0.85) | 0.77 (0.73, 0.82) | 0.79 (0.75, 0.83) | 0.77 (0.71, 0.84) |
| **Venous** | Composite venous | 0.62 (0.56, 0.70) | 0.65 (0.58, 0.73) | 0.64 (0.58, 0.69) | 0.61 (0.56, 0.65) | 0.54 (0.48, 0.61) | 0.63 (0.56, 0.70) | 0.66 (0.59, 0.73) | 0.64 (0.59, 0.70) | 0.61 (0.57, 0.66) | 0.56 (0.49, 0.63) |
| | Lower limb DVT[6] | 0.72 (0.60, 0.86) | 0.75 (0.63, 0.89) | 0.66 (0.58, 0.76) | 0.64 (0.57, 0.72) | 0.57 (0.47, 0.69) | 0.72 (0.61, 0.86) | 0.75 (0.63, 0.89) | 0.66 (0.58, 0.76) | 0.64 (0.57, 0.72) | 0.59 (0.48, 0.72) |
| | Pulmonary embolism | 0.56 (0.48, 0.65) | 0.59 (0.51, 0.69) | 0.63 (0.56, 0.70) | 0.59 (0.53, 0.65) | 0.53 (0.46, 0.62) | 0.56 (0.48, 0.65) | 0.60 (0.51, 0.69) | 0.63 (0.56, 0.71) | 0.59 (0.54, 0.65) | 0.54 (0.46, 0.64) |
| | ICVT[7]* | 0.43 (0.10, 1.81) | 0.65 (0.20, 2.14) | 0.63 (0.26, 1.55) | 0.78 (0.42, 1.47) | 0.93 (0.35, 2.47) | 0.39 (0.09, 1.63) | 0.61 (0.19, 1.98) | 0.61 (0.25, 1.48) | 0.69 (0.37, 1.29) | 0.81 (0.30, 2.20) |
| | Mesenteric thrombus | 0.46 (0.32, 0.68) | 0.51 (0.36, 0.73) | 0.64 (0.50, 0.82) | 0.54 (0.43, 0.68) | 0.42 (0.29, 0.61) | 0.47 (0.32, 0.70) | 0.53 (0.37, 0.76) | 0.66 (0.51, 0.85) | 0.57 (0.45, 0.72) | 0.45 (0.31, 0.66) |
| | Portal vein thrombosis | | 0.28 (0.11, 0.74) | | 0.51 (0.23, 1.12) | | | 0.24 (0.08, 0.68) | | 0.40 (0.17, 0.94) | |
| **Other** | Any thrombocytopenia | 0.90 (0.63, 1.29) | 0.65 (0.42, 1.01) | 1.08 (0.82, 1.43) | 0.85 (0.66, 1.09) | 1.28 (0.85, 1.92) | 0.84 (0.58, 1.20) | 0.59 (0.38, 0.92) | 0.97 (0.73, 1.27) | 0.71 (0.55, 0.91) | 1.02 (0.68, 1.54) |
| | SAH and HS[8] | 0.73 (0.59, 0.89) | 0.82 (0.67, 1.00) | 0.87 (0.75, 1.01) | 0.75 (0.65, 0.86) | 0.66 (0.52, 0.84) | 0.75 (0.61, 0.92) | 0.85 (0.70, 1.04) | 0.91 (0.78, 1.07) | 0.81 (0.70, 0.93) | 0.73 (0.58, 0.92) |
| | Myocarditis | 1.87 (1.10, 3.18) | 0.99 (0.48, 2.04) | 1.07 (0.59, 1.94) | 0.88 (0.53, 1.48) | 0.37 (0.08, 1.64) | 1.83 (1.07, 3.11) | 0.96 (0.46, 1.98) | 1.05 (0.58, 1.91) | 0.85 (0.51, 1.43) | 0.36 (0.08, 1.59) |
| | Pericarditis | 0.88 (0.35, 2.18) | 2.13 (1.09, 4.16) | 1.29 (0.68, 2.43) | 1.31 (0.78, 2.21) | 1.45 (0.47, 4.51) | 0.87 (0.35, 2.17) | 2.14 (1.10, 4.17) | 1.29 (0.68, 2.43) | 1.33 (0.79, 2.24) | 1.50 (0.48, 4.69) |

*Time periods collapsed into weeks 0-4 and weeks 5-24.

Supplementary Table 20: Hazard ratios and 95% CIs for individual arterial and venous thrombotic events, and other cardiovascular events, by time since booster vaccination with mRNA-1273 for those on a primary course of mRNA-1273 or BNT-162b2.

| Outcome | | Age and Sex adjusted HR (95% CI) | | | | | Fully Adjusted HR (95% CI) | | | | |
|---|---|---|---|---|---|---|---|---|---|---|---|
| | | Week 1 | Week 2 | Weeks 3-4 | Weeks 5-12 | Weeks 13-24 | Week 1 | Week 2 | Weeks 3-4 | Weeks 5-12 | Weeks 13-24 |
| **Arterial** | Composite arterial | 0.63 (0.51, 0.77) | 0.92 (0.78, 1.10) | 0.82 (0.71, 0.94) | 0.79 (0.70, 0.89) | 0.96 (0.30, 3.04) | 0.66 (0.54, 0.81) | 0.98 (0.83, 1.17) | 0.88 (0.76, 1.01) | 0.86 (0.76, 0.97) | 1.04 (0.32, 3.38) |
| | AMI[2]* | | 0.84 (0.73, 0.96) | | 0.81 (0.69, 0.95) | | | 0.89 (0.78, 1.02) | | 0.86 (0.73, 1.02) | |
| | Ischaemic stroke | 0.46 (0.33, 0.64) | 0.92 (0.72, 1.18) | 0.77 (0.63, 0.94) | 0.73 (0.61, 0.87) | NA (NA, NA) | 0.49 (0.35, 0.68) | 0.98 (0.77, 1.26) | 0.83 (0.68, 1.02) | 0.81 (0.67, 0.97) | 1.80 (0.53, 6.14) |
| **Venous** | Composite venous* | | 0.62 (0.51, 0.76) | | 0.60 (0.47, 0.76) | | | 0.61 (0.50, 0.75) | | 0.58 (0.45, 0.74) | |
| | Lower limb DVT[6]* | | 0.53 (0.38, 0.73) | | 0.52 (0.34, 0.78) | | | 0.53 (0.38, 0.73) | | 0.51 (0.34, 0.76) | |
| | Pulmonary embolism* | | 0.67 (0.52, 0.88) | | 0.67 (0.49, 0.92) | | | 0.70 (0.53, 0.91) | | 0.70 (0.51, 0.96) | |
| | ICVT[7]* | | 0.28 (0.04, 2.05) | | 0.36 (0.05, 2.81) | | | 0.26 (0.04, 1.79) | | 0.27 (0.04, 1.94) | |
| | Mesenteric thrombus* | | 0.66 (0.34, 1.28) | | 0.67 (0.32, 1.43) | | | 0.68 (0.35, 1.32) | | 0.68 (0.31, 1.46) | |
| | Portal vein thrombosis* | | 0.59 (0.08, 4.67) | | 0.66 (0.08, 5.28) | | | 0.58 (0.07, 4.65) | | 0.64 (0.08, 5.08) | |
| **Other** | Any thrombocytopenia* | | 1.11 (0.64, 1.95) | | 0.14 (0.02, 1.02) | | | 1.05 (0.59, 1.85) | | 0.12 (0.02, 0.90) | |
| | SAH and HS[8]* | | 0.67 (0.44, 1.03) | | 0.65 (0.36, 1.16) | | | 0.72 (0.47, 1.11) | | 0.71 (0.40, 1.27) | |
| | Myocarditis* | | 0.94 (0.40, 2.19) | | 0.47 (0.06, 3.61) | | | 0.90 (0.39, 2.11) | | 0.46 (0.06, 3.50) | |
| | Pericarditis* | | 2.02 (0.68, 6.03) | | 0.98 (0.10,10.16) | | | 2.04 (0.68, 6.07) | | 1.00 (0.10,10.31) | |

*Time periods collapsed into weeks 0-4 and weeks 5-24.

Supplementary Table 21: Hazard ratios and 95% CIs for individual arterial and venous thrombotic events, and other cardiovascular events, by time since booster vaccination with BNT-162b2 for those on a primary course of mRNA-1273 or BNT-162b2.

| **Outcome** | | **Age and Sex adjusted HR (95% CI)** | | | | | **Fully Adjusted HR (95% CI)** | | | | |
|---|---|---|---|---|---|---|---|---|---|---|---|
| | | Week 1 | Week 2 | Weeks 3-4 | Weeks 5-12 | Weeks 13-24 | Week 1 | Week 2 | Weeks 3-4 | Weeks 5-12 | Weeks 13-24 |
| **Arterial** | Composite arterial | 0.66 (0.63,0.70) | 0.75 (0.71,0.79) | 0.76 (0.73, 0.79) | 0.71 (0.69,0.74) | 0.64 (0.60,0.68) | 0.70 (0.66,0.74) | 0.79 (0.75,0.84) | 0.81 (0.78,0.85) | 0.80 (0.77,0.83) | 0.75 (0.70,0.79) |
| | AMI[2] | 0.66 (0.61,0.72) | 0.76 (0.71,0.82) | 0.80 (0.76,0.85) | 0.73 (0.69,0.77) | 0.63 (0.59,0.69) | 0.69 (0.64,0.74) | 0.80 (0.74,0.86) | 0.85 (0.80,0.90) | 0.79 (0.75,0.83) | 0.71 (0.65,0.77) |
| | Ischaemic stroke | 0.67 (0.62,0.72) | 0.74 (0.69,0.79) | 0.71 (0.67,0.75) | 0.70 (0.67,0.74) | 0.65 (0.60,0.71) | 0.71 (0.65,0.76) | 0.79 (0.73,0.85) | 0.77 (0.73,0.82) | 0.80 (0.75,0.84) | 0.77 (0.71,0.84) |
| **Venous** | Composite venous | 0.63 (0.56,0.71) | 0.67 (0.60,0.75) | 0.65 (0.59,0.71) | 0.61 (0.57,0.66) | 0.55 (0.49,0.62) | 0.63 (0.56,0.71) | 0.67 (0.59,0.75) | 0.64 (0.59,0.70) | 0.61 (0.57,0.66) | 0.56 (0.49,0.63) |
| | Lower limb DVT[6] | 0.72 (0.60,0.86) | 0.78 (0.65,0.93) | 0.68 (0.59,0.79) | 0.65 (0.58,0.73) | 0.57 (0.46,0.69) | 0.72 (0.60,0.86) | 0.78 (0.65,0.93) | 0.68 (0.59,0.79) | 0.65 (0.58,0.73) | 0.58 (0.48,0.71) |
| | Pulmonary embolism | 0.57 (0.49,0.67) | 0.60 (0.52,0.70) | 0.63 (0.56,0.71) | 0.59 (0.54,0.65) | 0.55 (0.47,0.65) | 0.58 (0.49,0.67) | 0.61 (0.52,0.71) | 0.64 (0.57,0.71) | 0.60 (0.54,0.66) | 0.56 (0.48,0.66) |
| | ICVT[7] | 0.50 (0.12,2.09) | 0.52 (0.12,2.16) | 0.69 (0.29,1.67) | 0.75 (0.41,1.38) | 0.85 (0.34,2.17) | 0.53 (0.13,2.19) | 0.51 (0.12,2.15) | 0.70 (0.29,1.68) | 0.72 (0.39,1.34) | 0.88 (0.34,2.26) |
| | Mesenteric thrombus | 0.48 (0.33,0.71) | 0.45 (0.31,0.67) | 0.67 (0.52,0.87) | 0.56 (0.45,0.70) | 0.43 (0.29,0.63) | 0.49 (0.33,0.72) | 0.46 (0.31,0.69) | 0.69 (0.53,0.89) | 0.58 (0.46,0.73) | 0.45 (0.31,0.67) |
| | Portal vein thrombosis* | 0.23 (0.08,0.68) | | | 0.45 (0.20,1.00) | | 0.21 (0.07,0.69) | | | 0.38 (0.17,0.87) | |
| **Other** | Any thrombocytopenia | 0.87 (0.59,1.28) | 0.57 (0.35,0.93) | 1.16 (0.87,1.53) | 0.91 (0.71,1.17) | 1.31 (0.87,1.98) | 0.77 (0.52,1.13) | 0.50 (0.30,0.81) | 1.00 (0.75,1.32) | 0.74 (0.57,0.95) | 1.05 (0.69,1.59) |
| | SAH and HS[8] | 0.72 (0.58,0.88) | 0.84 (0.69,1.02) | 0.85 (0.73,1.00) | 0.73 (0.64,0.85) | 0.63 (0.50,0.80) | 0.74 (0.60,0.92) | 0.88 (0.72,1.07) | 0.90 (0.77,1.05) | 0.80 (0.69,0.92) | 0.71 (0.56,0.90) |
| | Myocarditis | 1.88 (1.03,3.44) | 1.06 (0.49,2.30) | 1.32 (0.73,2.39) | 1.04 (0.62,1.73) | 0.43 (0.10,1.92) | 1.93 (1.06,3.53) | 1.11 (0.51,2.40) | 1.38 (0.76,2.48) | 1.06 (0.63,1.78) | 0.45 (0.10,1.99) |
| | Pericarditis | 0.98 (0.39,2.44) | 1.92 (0.94,3.93) | 1.14 (0.58,2.24) | 1.32 (0.79,2.20) | 1.39 (0.45,4.32) | 0.96 (0.38,2.39) | 1.83 (0.90,3.75) | 1.10 (0.56,2.18) | 1.28 (0.76,2.15) | 1.33 (0.43,4.14) |

*Time periods collapsed into weeks 0-4 and weeks 5-24.

Supplementary Table 22: Hazard ratios and 95% CIs for individual arterial and venous thrombotic events, and other cardiovascular events, by time since booster vaccination with mRNA-1273 or BNT-162b2 for those on a primary course of mRNA-1273 or BNT-162b2.

| **Outcome** | | **Age and Sex adjusted HR (95% CI)** | | | | | **Fully Adjusted HR (95% CI)** | | | | |
|---|---|---|---|---|---|---|---|---|---|---|---|
| | | Week 1 | Week 2 | Weeks 3-4 | Weeks 5-12 | Weeks 13-24 | Week 1 | Week 2 | Weeks 3-4 | Weeks 5-12 | Weeks 13-24 |
| **Arterial** | Composite arterial | 0.66 (0.63,0.70) | 0.76 (0.72,0.80) | 0.76 (0.73,0.79) | 0.71 (0.69,0.74) | 0.64 (0.60,0.67) | 0.69 (0.66,0.73) | 0.80 (0.76,0.84) | 0.81 (0.78,0.84) | 0.79 (0.76,0.81) | 0.73 (0.69,0.77) |
| | AMI[2] | 0.67 (0.62,0.72) | 0.77 (0.72,0.83) | 0.80 (0.76,0.85) | 0.72 (0.69,0.76) | 0.63 (0.58,0.68) | 0.70 (0.65,0.75) | 0.81 (0.75,0.87) | 0.85 (0.81,0.90) | 0.80 (0.76,0.84) | 0.71 (0.66,0.77) |
| | Ischaemic stroke | 0.66 (0.61,0.71) | 0.75 (0.70,0.81) | 0.72 (0.68,0.76) | 0.70 (0.67,0.74) | 0.65 (0.60,0.71) | 0.70 (0.65,0.75) | 0.80 (0.75,0.86) | 0.78 (0.74,0.83) | 0.80 (0.76,0.84) | 0.78 (0.72,0.84) |
| **Venous** | Composite venous | 0.63 (0.56,0.71) | 0.66 (0.59,0.74) | 0.64 (0.59,0.70) | 0.62 (0.57,0.66) | 0.55 (0.49,0.63) | 0.63 (0.56,0.71) | 0.66 (0.59,0.74) | 0.64 (0.59,0.70) | 0.62 (0.57,0.67) | 0.57 (0.50,0.64) |
| | Lower limb DVT[6] | 0.72 (0.60,0.86) | 0.76 (0.64,0.90) | 0.67 (0.58,0.76) | 0.65 (0.58,0.73) | 0.57 (0.47,0.70) | 0.72 (0.60,0.85) | 0.76 (0.64,0.90) | 0.66 (0.58,0.76) | 0.65 (0.58,0.73) | 0.58 (0.48,0.71) |
| | Pulmonary embolism | 0.57 (0.49,0.66) | 0.60 (0.52,0.69) | 0.63 (0.56,0.70) | 0.59 (0.53,0.64) | 0.53 (0.46,0.62) | 0.56 (0.48,0.66) | 0.60 (0.52,0.69) | 0.63 (0.56,0.70) | 0.58 (0.53,0.64) | 0.54 (0.46,0.63) |
| | ICVT[7] | 0.39 (0.09,1.64) | 0.61 (0.18,2.04) | 0.56 (0.23,1.37) | 0.71 (0.39,1.31) | 0.88 (0.34,2.28) | 0.42 (0.10,1.76) | 0.61 (0.19,2.01) | 0.60 (0.25,1.46) | 0.77 (0.42,1.42) | 0.91 (0.35,2.34) |
| | Mesenteric thrombus | 0.46 (0.32,0.68) | 0.51 (0.36,0.74) | 0.65 (0.51,0.84) | 0.56 (0.44,0.70) | 0.43 (0.30,0.63) | 0.48 (0.33,0.71) | 0.54 (0.37,0.77) | 0.69 (0.54,0.89) | 0.61 (0.48,0.76) | 0.49 (0.33,0.71) |
| | Portal vein thrombosis* | 0.25 (0.10,0.65) | | | 0.44 (0.20,0.94) | | 0.25 (0.10,0.66) | | | 0.39 (0.17,0.88) | |
| **Other** | Any thrombocytopenia | 0.93 (0.64,1.34) | 0.67 (0.43,1.04) | 1.14 (0.87,1.51) | 0.91 (0.70,1.16) | 1.39 (0.93,2.09) | 0.85 (0.59,1.22) | 0.60 (0.39,0.93) | 1.02 (0.77,1.35) | 0.76 (0.59,0.97) | 1.10 (0.73,1.66) |
| | SAH and HS[8] | 0.72 (0.58,0.88) | 0.81 (0.67,0.99) | 0.86 (0.74,1.00) | 0.74 (0.64,0.86) | 0.66 (0.52,0.83) | 0.75 (0.61,0.93) | 0.85 (0.70,1.04) | 0.92 (0.79,1.07) | 0.82 (0.71,0.95) | 0.75 (0.60,0.96) |
| | Myocarditis | 1.96 (1.18,3.25) | 0.98 (0.47,2.04) | 1.12 (0.62,2.01) | 0.98 (0.59,1.62) | 0.45 (0.10,1.97) | 1.93 (1.16,3.21) | 0.96 (0.46,2.00) | 1.07 (0.59,1.96) | 0.95 (0.57,1.58) | 0.41 (0.09,1.80) |
| | Pericarditis | 0.85 (0.34,2.10) | 2.04 (1.06,3.94) | 1.23 (0.66,2.32) | 1.29 (0.78,2.15) | 1.50 (0.49,4.61) | 0.88 (0.35,2.20) | 2.05 (1.07,3.93) | 1.29 (0.69,2.42) | 1.37 (0.82,2.28) | 1.68 (0.54,5.21) |

*Time periods collapsed into weeks 0-4 and weeks 5-24.

Supplementary Table 23: Hazard ratios and 95% CIs for individual arterial and venous thrombotic events, and other cardiovascular events, by time since booster vaccination with mRNA-1273 for those on a primary course of ChAdOx1 or mRNA-1273 or BNT-162b2.

| Outcome | | Age and Sex adjusted HR (95% CI) | | | | | Fully Adjusted HR (95% CI) | | | | |
|---|---|---|---|---|---|---|---|---|---|---|---|
| | | Week 1 | Week 2 | Weeks 3-4 | Weeks 5-12 | Weeks 13-24 | Week 1 | Week 2 | Weeks 3-4 | Weeks 5-12 | Weeks 13-24 |
| **Arterial** | Composite arterial | 0.65 (0.59,0.71) | 0.72 (0.66,0.79) | 0.67 (0.62,0.72) | 0.59 (0.56,0.64) | 0.43 (0.14,1.35) | 0.75 (0.68, 0.82) | 0.84 (0.77, 0.92) | 0.79 (0.74, 0.85) | 0.73 (0.68, 0.78) | 0.55 (0.18, 1.74) |
| | AMI[2]* | 0.71 (0.66,0.76) | | | 0.64 (0.59,0.70) | | 0.82 (0.76, 0.88) | | | 0.75 (0.69, 0.82) | |
| | Ischaemic stroke | 0.61 (0.53,0.70) | 0.73 (0.64,0.84) | 0.58 (0.52,0.65) | 0.52 (0.47,0.57) | 0.82 (0.26,2.59) | 0.71 (0.61, 0.82) | 0.86 (0.75, 0.99) | 0.69 (0.62, 0.78) | 0.65 (0.58, 0.72) | 0.88 (0.27, 2.90) |
| **Venous** | Composite venous* | 0.57 (0.52,0.64) | | | 0.54 (0.48,0.62) | | 0.61 (0.55, 0.68) | | | 0.58 (0.51, 0.66) | |
| | Lower limb DVT[6]* | 0.59 (0.51,0.70) | | | 0.55 (0.45,0.68) | | 0.62 (0.53, 0.73) | | | 0.59 (0.48, 0.72) | |
| | Pulmonary embolism* | 0.54 (0.47,0.62) | | | 0.54 (0.46,0.65) | | 0.58 (0.50, 0.67) | | | 0.59 (0.49, 0.70) | |
| | ICVT[7]* | 0.72 (0.30,1.68) | | | 0.69 (0.27,1.78) | | 0.74 (0.32, 1.75) | | | 0.70 (0.27, 1.80) | |
| | Mesenteric thrombus* | 0.48 (0.33,0.71) | | | 0.44 (0.27,0.71) | | 0.54 (0.36, 0.79) | | | 0.50 (0.31, 0.81) | |
| | Portal vein thrombosis | 0.76 (0.34,1.71) | | | 0.09 (0.01,0.76) | | 0.90 (0.40, 2.02) | | | 0.10 (0.01, 0.79) | |
| **Other** | Any thrombocytopenia* | 0.87 (0.61,1.25) | | | 0.44 (0.23,0.82) | | 0.93 (0.65, 1.32) | | | 0.45 (0.24, 0.84) | |
| | SAH and HS[8] | 0.40 (0.25,0.65) | 0.49 (0.31,0.76) | 0.63 (0.46,0.86) | 0.56 (0.42,0.76) | NA (NA, NA) | 0.45 (0.28, 0.73) | 0.55 (0.36, 0.87) | 0.74 (0.54, 1.00) | 0.68 (0.51, 0.92) | 4.55 (0.59,35.00) |
| | Myocarditis* | 0.84 (0.48,1.47) | | | 0.75 (0.30,1.88) | | 0.86 (0.49, 1.51) | | | 0.74 (0.29, 1.84) | |
| | Pericarditis* | 0.97 (0.48,1.93) | | | 0.30 (0.06,1.42) | | 1.01 (0.51, 2.02) | | | 0.32 (0.07, 1.52) | |

*Time periods collapsed into weeks 0-4 and weeks 5-24.
NA (NA-NA) Could not obtain estimate for this time interval

Supplementary Table 24: Hazard ratios and 95% CIs for individual arterial and venous thrombotic events, and other cardiovascular events, by time since booster vaccination with BNT-162b2 for those on a primary course of ChAdOx1 or mRNA-1273 or BNT-162b2.

| Outcome | | Age and Sex adjusted HR (95% CI) | | | | | Fully Adjusted HR (95% CI) | | | | |
|---|---|---|---|---|---|---|---|---|---|---|---|
| | | Week 1 | Week 2 | Weeks 3-4 | Weeks 5-12 | Weeks 13-24 | Week 1 | Week 2 | Weeks 3-4 | Weeks 5-12 | Weeks 13-24 |
| **Arterial** | Composite arterial | 0.73 (0.70,0.75) | 0.79 (0.76,0.81) | 0.79 (0.77,0.81) | 0.74 (0.72,0.76) | 0.65 (0.63,0.68) | 0.75 (0.72,0.78) | 0.82 (0.79,0.85) | 0.83 (0.81,0.86) | 0.80 (0.78,0.82) | 0.75 (0.72,0.78) |
| | AMI[2] | 0.73 (0.69,0.77) | 0.81 (0.77,0.85) | 0.82 (0.79,0.85) | 0.75 (0.73,0.78) | 0.66 (0.62,0.70) | 0.74 (0.70,0.77) | 0.82 (0.78,0.86) | 0.83 (0.80,0.87) | 0.78 (0.76,0.81) | 0.71 (0.67,0.75) |
| | Ischaemic stroke | 0.73 (0.69,0.77) | 0.77 (0.73,0.81) | 0.77 (0.74,0.80) | 0.73 (0.70,0.75) | 0.66 (0.62,0.70) | 0.77 (0.73,0.81) | 0.82 (0.78,0.86) | 0.83 (0.80,0.87) | 0.84 (0.81,0.87) | 0.82 (0.77,0.87) |
| **Venous** | Composite venous | 0.67 (0.62,0.73) | 0.73 (0.68,0.79) | 0.72 (0.68,0.76) | 0.66 (0.63,0.69) | 0.60 (0.54,0.66) | 0.66 (0.61,0.72) | 0.72 (0.67,0.78) | 0.71 (0.67,0.75) | 0.65 (0.62,0.68) | 0.60 (0.54,0.66) |
| | Lower limb DVT[6] | 0.73 (0.65,0.83) | 0.86 (0.77,0.96) | 0.79 (0.72,0.86) | 0.70 (0.65,0.76) | 0.67 (0.57,0.77) | 0.72 (0.64,0.82) | 0.85 (0.76,0.96) | 0.78 (0.71,0.86) | 0.70 (0.64,0.75) | 0.67 (0.58,0.78) |
| | Pulmonary embolism | 0.63 (0.57,0.70) | 0.66 (0.59,0.73) | 0.67 (0.62,0.73) | 0.63 (0.59,0.67) | 0.56 (0.49,0.63) | 0.62 (0.56,0.69) | 0.65 (0.59,0.72) | 0.66 (0.61,0.72) | 0.62 (0.58,0.66) | 0.55 (0.49,0.62) |
| | ICVT[7] | 0.49 (0.18,1.35) | 0.78 (0.34,1.77) | 0.75 (0.40,1.42) | 0.83 (0.51,1.33) | 0.89 (0.37,2.17) | 0.47 (0.17,1.29) | 0.73 (0.32,1.65) | 0.68 (0.36,1.29) | 0.71 (0.43,1.15) | 0.72 (0.29,1.75) |
| | Mesenteric thrombus | 0.52 (0.40,0.68) | 0.54 (0.42,0.71) | 0.63 (0.52,0.76) | 0.63 (0.54,0.74) | 0.56 (0.42,0.74) | 0.52 (0.40,0.68) | 0.55 (0.42,0.71) | 0.64 (0.53,0.77) | 0.65 (0.55,0.76) | 0.60 (0.45,0.79) |
| | Portal vein thrombosis | 0.57 (0.23,1.40) | 0.12 (0.02,0.86) | 0.39 (0.17,0.87) | 0.57 (0.33,0.97) | 0.29 (0.08,1.06) | 0.57 (0.22,1.47) | 0.10 (0.01,0.76) | 0.39 (0.17,0.92) | 0.62 (0.34,1.14) | 0.37 (0.09,1.52) |
| **Other** | Any thrombocytopenia | 1.16 (0.91,1.49) | 0.98 (0.74,1.28) | 1.30 (1.07,1.58) | 1.18 (0.99,1.41) | 2.00 (1.46,2.75) | 0.99 (0.78,1.27) | 0.82 (0.62,1.07) | 1.06 (0.87,1.28) | 0.88 (0.74,1.05) | 1.40 (1.02,1.92) |
| | SAH and HS[8] | 0.69 (0.60,0.80) | 0.78 (0.68,0.90) | 0.77 (0.69,0.86) | 0.75 (0.68,0.82) | 0.65 (0.55,0.78) | 0.72 (0.62,0.84) | 0.82 (0.72,0.95) | 0.82 (0.74,0.92) | 0.83 (0.75,0.91) | 0.76 (0.64,0.91) |
| | Myocarditis | 1.73 (1.12,2.69) | 1.10 (0.64,1.89) | 1.18 (0.77,1.79) | 0.94 (0.65,1.36) | 0.42 (0.13,1.39) | 1.65 (1.07,2.57) | 1.06 (0.62,1.82) | 1.11 (0.73,1.69) | 0.85 (0.59,1.24) | 0.37 (0.11,1.24) |
| | Pericarditis | 0.79 (0.40,1.55) | 1.75 (1.07,2.87) | 0.77 (0.45,1.30) | 1.27 (0.88,1.83) | 0.87 (0.34,2.20) | 0.78 (0.40,1.54) | 1.73 (1.05,2.83) | 0.76 (0.44,1.28) | 1.22 (0.85,1.76) | 0.85 (0.34,2.14) |

Supplementary Table 25: Hazard ratios and 95% CIs for individual arterial and venous thrombotic events, and other cardiovascular events, by time since booster vaccination with mRNA-1273 or BNT-162b2 for those on a primary course of ChAdOx1 or mRNA-1273 or BNT-162b2.

| Outcome | | Age and Sex adjusted HR (95% CI) | | | | | Fully Adjusted HR (95% CI) | | | | |
|---|---|---|---|---|---|---|---|---|---|---|---|
| | | Week 1 | Week 2 | Weeks 3-4 | Weeks 5-12 | Weeks 13-24 | Week 1 | Week 2 | Weeks 3-4 | Weeks 5-12 | Weeks 13-24 |
| **Arterial thrombotic events** | Composite arterial | 0.72 (0.69,0.74) | 0.78 (0.75,0.81) | 0.78 (0.76,0.80) | 0.73 (0.72,0.75) | 0.66 (0.63,0.69) | 0.75 (0.72,0.77) | 0.82 (0.79,0.84) | 0.82 (0.80,0.85) | 0.79 (0.77,0.81) | 0.74 (0.71,0.77) |
| | AMI[2] | 0.72 (0.69,0.76) | 0.79 (0.75,0.83) | 0.80 (0.78,0.83) | 0.75 (0.72,0.77) | 0.66 (0.62,0.70) | 0.74 (0.71,0.78) | 0.82 (0.78,0.85) | 0.83 (0.80,0.86) | 0.78 (0.75,0.80) | 0.70 (0.66,0.74) |
| | Ischaemic stroke | 0.72 (0.68,0.75) | 0.77 (0.73,0.81) | 0.75 (0.73,0.78) | 0.73 (0.70,0.75) | 0.68 (0.64,0.72) | 0.76 (0.73,0.80) | 0.82 (0.78,0.86) | 0.82 (0.79,0.85) | 0.82 (0.79,0.85) | 0.80 (0.75,0.85) |
| **Venous thromboembolic events** | Composite venous | 0.66 (0.61,0.71) | 0.73 (0.68,0.78) | 0.70 (0.66,0.74) | 0.67 (0.64,0.70) | 0.61 (0.56,0.67) | 0.66 (0.61,0.71) | 0.72 (0.67,0.78) | 0.69 (0.66,0.73) | 0.65 (0.62,0.69) | 0.60 (0.55,0.66) |
| | Lower limb DVT[6] | 0.70 (0.63,0.79) | 0.84 (0.76,0.94) | 0.77 (0.70,0.84) | 0.70 (0.65,0.76) | 0.68 (0.59,0.79) | 0.70 (0.62,0.79) | 0.84 (0.75,0.94) | 0.76 (0.70,0.83) | 0.69 (0.64,0.75) | 0.67 (0.58,0.78) |
| | Pulmonary embolism | 0.62 (0.56,0.68) | 0.65 (0.59,0.71) | 0.65 (0.61,0.70) | 0.63 (0.59,0.67) | 0.56 (0.50,0.63) | 0.62 (0.56,0.68) | 0.65 (0.59,0.71) | 0.65 (0.61,0.70) | 0.62 (0.58,0.66) | 0.56 (0.50,0.63) |
| | ICVT[7] | 0.59 (0.25,1.35) | 0.92 (0.47,1.83) | 0.62 (0.33,1.16) | 0.81 (0.51,1.29) | 0.86 (0.35,2.08) | 0.54 (0.23,1.24) | 0.84 (0.42,1.66) | 0.55 (0.29,1.02) | 0.66 (0.42,1.06) | 0.68 (0.28,1.67) |
| | Mesenteric thrombus | 0.50 (0.38,0.65) | 0.55 (0.43,0.71) | 0.65 (0.54,0.78) | 0.63 (0.54,0.74) | 0.58 (0.44,0.76) | 0.51 (0.39,0.66) | 0.56 (0.44,0.73) | 0.67 (0.56,0.80) | 0.65 (0.56,0.77) | 0.62 (0.46,0.82) |
| | Portal vein thrombosis | 0.55 (0.24,1.27) | 0.27 (0.08,0.87) | 0.50 (0.26,0.97) | 0.53 (0.32,0.90) | 0.31 (0.08,1.15) | 0.51 (0.21,1.23) | 0.27 (0.08,0.88) | 0.47 (0.23,0.93) | 0.43 (0.25,0.73) | 0.24 (0.06,1.01) |
| **Other cardiovascular events** | Any thrombocytopenia | 1.13 (0.89,1.42) | 0.98 (0.76,1.27) | 1.29 (1.06,1.55) | 1.19 (1.00,1.41) | 2.21 (1.61,3.03) | 1.01 (0.80,1.28) | 0.86 (0.67,1.11) | 1.11 (0.92,1.33) | 0.93 (0.78,1.10) | 1.57 (1.15,2.15) |
| | SAH and HS[8] | 0.66 (0.57,0.76) | 0.75 (0.65,0.86) | 0.76 (0.68,0.84) | 0.74 (0.68,0.82) | 0.68 (0.57,0.80) | 0.69 (0.59,0.79) | 0.79 (0.69,0.90) | 0.80 (0.72,0.89) | 0.81 (0.74,0.89) | 0.77 (0.64,0.91) |
| | Myocarditis | 1.71 (1.17,2.50) | 1.04 (0.64,1.70) | 0.96 (0.63,1.46) | 0.91 (0.63,1.30) | 0.41 (0.12,1.36) | 1.69 (1.15,2.47) | 1.05 (0.64,1.71) | 0.95 (0.63,1.45) | 0.87 (0.60,1.24) | 0.40 (0.12,1.34) |
| | Pericarditis | 0.64 (0.32,1.26) | 1.69 (1.05,2.70) | 0.94 (0.60,1.50) | 1.27 (0.88,1.84) | 1.02 (0.40,2.59) | 0.61 (0.31,1.20) | 1.60 (1.00,2.54) | 0.88 (0.56,1.39) | 1.11 (0.77,1.59) | 0.83 (0.33,2.10) |

Supplementary Table 26: Subgroup-specific maximally-adjusted hazard ratios and 95% CIs for composite arterial and composite venous events by time since first dose vaccination.

| Vaccination course | Event | Stratum | Week 1 | Week 2 | Weeks 3-4 | Weeks 5-12 | Weeks 13-24 | Weeks 25-26 |
|---|---|---|---|---|---|---|---|---|
| Dose 1: BNT-162b2 | Composite arterial | Age group: <40 | 0.97 (0.72,1.30) | 0.94 (0.70,1.27) | 0.90 (0.72,1.13) | 1.16 (1.02,1.32) | 1.00 (0.87,1.15) | 0.93 (0.72,1.21) |
| | | Age group: 40-59 | 0.87 (0.78,0.99) | 0.99 (0.88,1.11) | 1.00 (0.92,1.09) | 0.91 (0.85,0.96) | 0.94 (0.88,1.00) | 0.99 (0.89,1.10) |
| | | Age group: 60-79 | 0.75 (0.70,0.81) | 0.84 (0.79,0.90) | 0.86 (0.81,0.90) | 0.88 (0.84,0.92) | 0.94 (0.90,0.98) | 0.96 (0.90,1.03) |
| | | Age group: >=80 | 0.62 (0.58,0.67) | 0.67 (0.62,0.72) | 0.72 (0.68,0.77) | 0.77 (0.73,0.80) | 0.85 (0.81,0.90) | 0.89 (0.82,0.96) |
| | | Ethnicity: White | 0.72 (0.68,0.75) | 0.79 (0.76,0.83) | 0.81 (0.78,0.84) | 0.82 (0.80,0.84) | 0.86 (0.83,0.88) | 0.85 (0.81,0.88) |
| | | Ethnicity: Black or Black British | 0.51 (0.34,0.78) | 0.71 (0.50,1.02) | 0.95 (0.76,1.20) | 0.92 (0.79,1.07) | 0.84 (0.73,0.97) | 0.83 (0.62,1.10) |
| | | Ethnicity: Asian or Asian British | 0.79 (0.65,0.96) | 0.97 (0.81,1.16) | 0.99 (0.87,1.13) | 0.99 (0.90,1.08) | 1.07 (0.98,1.17) | 1.25 (1.07,1.47) |
| | | Ethnicity: Other Ethnic Groups | 0.79 (0.49,1.27) | 0.54 (0.30,0.95) | 0.92 (0.66,1.28) | 0.96 (0.77,1.21) | 1.14 (0.92,1.42) | 1.14 (0.77,1.68) |
| | | Ethnicity: Mixed | 0.90 (0.51,1.56) | 1.08 (0.63,1.84) | 1.02 (0.68,1.53) | 0.90 (0.69,1.17) | 0.80 (0.61,1.06) | 0.92 (0.56,1.54) |
| | | Ethnicity: Unknown | 1.01 (0.63,1.63) | 0.82 (0.47,1.42) | 1.20 (0.86,1.69) | 1.40 (1.12,1.75) | 1.99 (1.59,2.49) | 1.73 (1.17,2.56) |
| | | No prior history of COVID-19 | 0.73 (0.70,0.77) | 0.82 (0.78,0.86) | 0.84 (0.81,0.87) | 0.86 (0.84,0.88) | 0.91 (0.89,0.94) | 0.92 (0.88,0.96) |
| | | Prior history of COVID-19 | 0.63 (0.48,0.83) | 0.53 (0.39,0.73) | 0.77 (0.63,0.94) | 0.78 (0.68,0.90) | 0.89 (0.75,1.04) | 0.79 (0.60,1.05) |
| | | No prior history of event | 0.70 (0.66,0.75) | 0.80 (0.76,0.86) | 0.86 (0.82,0.90) | 0.89 (0.86,0.92) | 0.94 (0.91,0.97) | 0.97 (0.92,1.02) |
| | | Prior history of event | 0.76 (0.71,0.81) | 0.82 (0.77,0.87) | 0.83 (0.79,0.87) | 0.84 (0.81,0.87) | 0.95 (0.92,0.99) | 0.99 (0.93,1.05) |
| | | Sex: Female | 0.68 (0.63,0.73) | 0.74 (0.69,0.80) | 0.79 (0.75,0.83) | 0.82 (0.79,0.85) | 0.83 (0.80,0.87) | 0.83 (0.78,0.89) |
| | | Sex: Male | 0.78 (0.73,0.82) | 0.87 (0.82,0.92) | 0.89 (0.85,0.93) | 0.91 (0.88,0.94) | 0.99 (0.96,1.02) | 1.01 (0.96,1.06) |
| | Composite venous | Age group: <40 | 0.66 (0.50,0.88) | 0.85 (0.66,1.09) | 0.75 (0.62,0.91) | 0.70 (0.62,0.78) | 0.65 (0.58,0.72) | 0.53 (0.42,0.66) |
| | | Age group: 40-59 | 0.77 (0.60,0.97) | 0.94 (0.76,1.17) | 0.91 (0.77,1.07) | 0.92 (0.84,1.02) | 0.92 (0.84,1.01) | 0.84 (0.70,1.02) |
| | | Age group: 60-79 | 0.66 (0.57,0.78) | 0.74 (0.64,0.86) | 0.62 (0.55,0.70) | 0.68 (0.63,0.75) | 0.90 (0.82,0.98) | 0.87 (0.75,1.01) |
| | | Age group: >=80 | 0.50 (0.41,0.61) | 0.58 (0.48,0.71) | 0.62 (0.54,0.72) | 0.61 (0.54,0.69) | 0.84 (0.75,0.95) | 0.92 (0.77,1.11) |
| | | Ethnicity: White | 0.65 (0.59,0.72) | 0.76 (0.69,0.84) | 0.73 (0.68,0.79) | 0.76 (0.73,0.80) | 0.80 (0.77,0.84) | 0.75 (0.69,0.82) |
| | | Ethnicity: Black or Black British | 0.89 (0.49,1.63) | 1.13 (0.66,1.93) | 0.77 (0.48,1.22) | 0.92 (0.72,1.17) | 0.95 (0.77,1.18) | 0.89 (0.56,1.41) |
| | | Ethnicity: Asian or Asian British | 0.51 (0.25,1.04) | 1.02 (0.60,1.73) | 1.18 (0.83,1.69) | 0.96 (0.76,1.21) | 1.07 (0.86,1.34) | 0.56 (0.31,0.98) |
| | | Ethnicity: Other Ethnic Groups | 1.03 (0.40,2.64) | 1.42 (0.63,3.18) | 0.59 (0.24,1.47) | 0.89 (0.57,1.39) | 1.08 (0.74,1.57) | 1.10 (0.51,2.38) |
| | | Ethnicity: Mixed | 0.63 (0.20,1.99) | 0.88 (0.32,2.44) | 0.68 (0.30,1.57) | 0.62 (0.39,0.99) | 0.61 (0.39,0.93) | 0.55 (0.23,1.32) |
| | | Ethnicity: Unknown | 0.98 (0.35,2.71) | 0.55 (0.13,2.29) | 1.30 (0.64,2.64) | 1.42 (0.92,2.17) | 1.80 (1.21,2.69) | 1.48 (0.61,3.56) |
| | | No prior history of COVID-19 | 0.68 (0.62,0.76) | 0.80 (0.73,0.88) | 0.76 (0.71,0.82) | 0.80 (0.77,0.84) | 0.86 (0.82,0.90) | 0.79 (0.73,0.86) |
| | | Prior history of COVID-19 | 0.32 (0.15,0.68) | 0.63 (0.36,1.10) | 0.80 (0.55,1.17) | 0.66 (0.51,0.86) | 0.72 (0.55,0.95) | 0.62 (0.36,1.07) |
| | | No prior history of event | 0.66 (0.59,0.73) | 0.80 (0.73,0.89) | 0.76 (0.70,0.81) | 0.78 (0.74,0.82) | 0.84 (0.81,0.88) | 0.75 (0.69,0.82) |
| | | Prior history of event | 0.68 (0.53,0.88) | 0.69 (0.53,0.89) | 0.71 (0.59,0.85) | 0.77 (0.69,0.86) | 0.77 (0.69,0.86) | 0.87 (0.72,1.06) |
| | | Sex: Female | 0.71 (0.62,0.81) | 0.73 (0.64,0.84) | 0.73 (0.66,0.81) | 0.78 (0.74,0.83) | 0.82 (0.77,0.87) | 0.76 (0.68,0.84) |
| | | Sex: Male | 0.63 (0.54,0.74) | 0.88 (0.77,1.01) | 0.81 (0.73,0.89) | 0.82 (0.77,0.88) | 0.90 (0.84,0.96) | 0.82 (0.73,0.92) |
| Dose 1: ChAdOx1 | Composite arterial | Age group: <40 | 0.97 (0.67,1.39) | 1.11 (0.79,1.57) | 1.10 (0.84,1.42) | 1.24 (1.05,1.48) | 0.99 (0.83,1.18) | 1.08 (0.80,1.45) |
| | | Age group: 40-59 | 0.82 (0.76,0.89) | 1.04 (0.96,1.12) | 1.03 (0.97,1.09) | 0.98 (0.93,1.02) | 1.00 (0.95,1.05) | 1.01 (0.93,1.08) |
| | | Age group: 60-79 | 0.82 (0.77,0.86) | 0.90 (0.86,0.96) | 0.91 (0.87,0.95) | 0.94 (0.91,0.98) | 0.97 (0.94,1.01) | 1.00 (0.95,1.06) |
| | | Age group: >=80 | 0.73 (0.67,0.80) | 0.83 (0.76,0.91) | 0.85 (0.79,0.92) | 0.90 (0.85,0.95) | 0.97 (0.91,1.03) | 0.95 (0.87,1.04) |
| | | Ethnicity: White | 0.81 (0.77,0.84) | 0.94 (0.90,0.98) | 0.93 (0.90,0.96) | 0.94 (0.92,0.97) | 0.94 (0.91,0.96) | 0.92 (0.89,0.96) |
| | | Ethnicity: Black or Black British | 0.98 (0.76,1.27) | 1.13 (0.89,1.45) | 0.99 (0.81,1.20) | 1.08 (0.95,1.22) | 0.93 (0.82,1.06) | 0.82 (0.65,1.04) |
| | | Ethnicity: Asian or Asian British | 0.99 (0.84,1.16) | 1.09 (0.93,1.27) | 1.23 (1.09,1.38) | 1.24 (1.14,1.35) | 1.27 (1.16,1.39) | 1.19 (1.03,1.37) |
| | | Ethnicity: Other Ethnic Groups | 1.21 (0.85,1.70) | 1.17 (0.82,1.67) | 1.06 (0.80,1.41) | 1.22 (1.01,1.47) | 1.16 (0.95,1.41) | 1.31 (0.95,1.80) |
| | | Ethnicity: Mixed | 0.93 (0.58,1.51) | 0.96 (0.59,1.56) | 1.00 (0.70,1.45) | 1.00 (0.78,1.27) | 1.25 (0.98,1.59) | 1.38 (0.92,2.09) |
| | | Ethnicity: Unknown | 1.10 (0.74,1.64) | 0.95 (0.62,1.45) | 1.94 (1.51,2.49) | 1.94 (1.62,2.33) | 1.99 (1.65,2.40) | 2.49 (1.86,3.33) |
| | | No prior history of COVID-19 | 0.83 (0.80,0.86) | 0.97 (0.93,1.01) | 0.97 (0.94,1.00) | 1.00 (0.98,1.02) | 1.00 (0.97,1.02) | 0.99 (0.95,1.03) |
| | | Prior history of COVID-19 | 0.93 (0.78,1.13) | 0.83 (0.68,1.02) | 0.87 (0.74,1.02) | 0.83 (0.73,0.95) | 0.82 (0.70,0.95) | 0.87 (0.69,1.10) |
| | | No prior history of event | 0.85 (0.81,0.90) | 1.04 (0.99,1.10) | 1.05 (1.01,1.09) | 1.04 (1.01,1.07) | 1.04 (1.01,1.07) | 1.03 (0.99,1.08) |
| | | Prior history of event | 0.81 (0.76,0.86) | 0.87 (0.82,0.92) | 0.86 (0.82,0.90) | 0.93 (0.90,0.96) | 0.97 (0.93,1.01) | 0.99 (0.93,1.05) |
| | | Sex: Female | 0.78 (0.73,0.84) | 0.91 (0.86,0.97) | 0.90 (0.86,0.95) | 0.91 (0.88,0.95) | 0.90 (0.86,0.94) | 0.86 (0.81,0.92) |
| | | Sex: Male | 0.87 (0.83,0.92) | 1.00 (0.96,1.06) | 1.02 (0.98,1.06) | 1.06 (1.03,1.09) | 1.08 (1.05,1.12) | 1.11 (1.05,1.16) |
| | Composite venous | Age group: <40 | 1.17 (0.87,1.59) | 2.02 (1.59,2.58) | 1.33 (1.08,1.65) | 1.09 (0.95,1.24) | 0.90 (0.79,1.02) | 0.70 (0.54,0.90) |
| | | Age group: 40-59 | 0.64 (0.55,0.76) | 1.12 (0.99,1.28) | 1.14 (1.03,1.26) | 0.95 (0.89,1.03) | 0.94 (0.87,1.01) | 0.86 (0.75,0.97) |
| | | Age group: 60-79 | 0.76 (0.68,0.86) | 0.83 (0.74,0.92) | 0.74 (0.68,0.82) | 0.78 (0.72,0.84) | 0.83 (0.74,0.94) | 0.76 (0.68,0.86) |
| | | Age group: >=80 | 0.78 (0.64,0.97) | 0.70 (0.56,0.88) | 0.75 (0.63,0.90) | 0.81 (0.70,0.94) | 1.03 (0.90,1.19) | 1.06 (0.85,1.32) |
| | | Ethnicity: White | 0.80 (0.73,0.87) | 1.01 (0.93,1.09) | 0.97 (0.91,1.03) | 0.90 (0.86,0.94) | 0.92 (0.88,0.97) | 0.83 (0.77,0.89) |
| | | Ethnicity: Black or Black British | 0.93 (0.56,1.57) | 1.06 (0.65,1.73) | 1.26 (0.89,1.78) | 1.17 (0.94,1.44) | 0.87 (0.71,1.07) | 0.86 (0.58,1.27) |
| | | Ethnicity: Asian or Asian British | 0.83 (0.50,1.38) | 0.85 (0.51,1.43) | 0.87 (0.60,1.26) | 1.05 (0.84,1.31) | 1.03 (0.82,1.31) | 0.62 (0.38,1.03) |
| | | Ethnicity: Other Ethnic Groups | 0.69 (0.24,1.97) | 1.58 (0.75,3.33) | 0.85 (0.43,1.69) | 0.96 (0.66,1.41) | 0.91 (0.63,1.31) | 0.30 (0.10,0.84) |
| | | Ethnicity: Mixed | 0.40 (0.10,1.64) | 1.15 (0.50,2.66) | 0.29 (0.09,0.92) | 0.84 (0.56,1.27) | 0.72 (0.49,1.06) | 1.18 (0.64,2.19) |
| | | Ethnicity: Unknown | 1.39 (0.67,2.85) | 1.07 (0.46,2.50) | 1.91 (1.15,3.16) | 1.58 (1.12,2.22) | 2.11 (1.50,2.98) | 2.21 (1.24,3.93) |
| | | No prior history of COVID-19 | 0.80 (0.73,0.87) | 1.01 (0.94,1.09) | 0.97 (0.92,1.03) | 0.91 (0.88,0.95) | 0.93 (0.89,0.98) | 0.83 (0.78,0.90) |
| | | Prior history of COVID-19 | 0.89 (0.59,1.33) | 1.10 (0.75,1.61) | 1.09 (0.81,1.46) | 1.04 (0.84,1.30) | 1.04 (0.81,1.33) | 0.88 (0.57,1.36) |
| | | No prior history of event | 0.83 (0.76,0.90) | 1.02 (0.94,1.11) | 0.96 (0.91,1.03) | 0.93 (0.89,0.97) | 0.94 (0.90,0.99) | 0.82 (0.76,0.88) |
| | | Prior history of event | 0.64 (0.51,0.80) | 0.93 (0.77,1.12) | 0.99 (0.86,1.14) | 0.78 (0.71,0.87) | 0.83 (0.74,0.92) | 0.86 (0.73,1.03) |
| | | Sex: Female | 0.82 (0.73,0.92) | 1.01 (0.91,1.12) | 0.97 (0.89,1.05) | 0.88 (0.83,0.93) | 0.90 (0.85,0.96) | 0.83 (0.75,0.92) |
| | | Sex: Male | 0.81 (0.72,0.91) | 1.05 (0.95,1.17) | 1.03 (0.95,1.12) | 1.01 (0.95,1.07) | 1.04 (0.97,1.10) | 0.91 (0.82,1.00) |

Supplementary Table 27: Subgroup-specific maximally-adjusted hazard ratios and 95% CIs for composite arterial and composite venous events by time since COVID-19 dose 1&2 vaccination.

| Vaccination course | Event | Stratum | Week 1 | Week 2 | Weeks 3-4 | Weeks 5-12 | Weeks 13-24 | Weeks 25-26 |
|---|---|---|---|---|---|---|---|---|
| **Doses 1&2: BNT-162b2** | Composite arterial | Age group: <40 | 1.02 (0.73,1.42) | 1.22 (0.89,1.66) | 0.85 (0.65,1.13) | 1.03 (0.84,1.27) | 1.11 (0.86,1.43) | 1.33 (0.88,2.00) |
| | | Age group: 40-59 | 0.73 (0.64,0.83) | 0.82 (0.72,0.93) | 0.85 (0.77,0.94) | 0.78 (0.72,0.84) | 0.70 (0.64,0.77) | 0.63 (0.55,0.72) |
| | | Age group: 60-79 | 0.72 (0.67,0.78) | 0.82 (0.76,0.88) | 0.77 (0.73,0.83) | 0.70 (0.66,0.75) | 0.65 (0.60,0.70) | 0.62 (0.57,0.68) |
| | | Age group: >=80 | 0.72 (0.67,0.78) | 0.79 (0.73,0.85) | 0.84 (0.79,0.89) | 0.86 (0.82,0.90) | 0.83 (0.78,0.88) | 0.82 (0.75,0.89) |
| | | Ethnicity: White | 0.72 (0.69,0.76) | 0.81 (0.78,0.86) | 0.81 (0.78,0.85) | 0.81 (0.78,0.84) | 0.81 (0.78,0.84) | 0.81 (0.76,0.86) |
| | | Ethnicity: Black or Black British | 0.71 (0.48,1.05) | 0.99 (0.70,1.40) | 0.86 (0.65,1.14) | 0.73 (0.58,0.91) | 0.73 (0.57,0.92) | 0.71 (0.49,1.05) |
| | | Ethnicity: Asian or Asian British | 0.75 (0.62,0.92) | 0.84 (0.69,1.02) | 0.91 (0.78,1.05) | 0.82 (0.73,0.92) | 0.74 (0.65,0.85) | 0.75 (0.62,0.92) |
| | | Ethnicity: Other Ethnic Groups | 1.08 (0.70,1.66) | 0.94 (0.59,1.48) | 0.77 (0.52,1.13) | 0.80 (0.60,1.06) | 0.72 (0.52,0.99) | 0.82 (0.50,1.35) |
| | | Ethnicity: Mixed | 0.51 (0.25,1.04) | 0.27 (0.10,0.68) | 0.56 (0.33,0.95) | 0.71 (0.47,1.05) | 0.76 (0.50,1.16) | 0.86 (0.43,1.73) |
| | | Ethnicity: Unknown | 0.65 (0.39,1.08) | 0.59 (0.34,1.03) | 0.74 (0.50,1.10) | 0.84 (0.61,1.14) | 0.60 (0.41,0.86) | 0.42 (0.24,0.75) |
| | | No prior history of COVID-19 | 0.72 (0.69,0.76) | 0.81 (0.77,0.85) | 0.81 (0.78,0.84) | 0.80 (0.78,0.83) | 0.79 (0.76,0.82) | 0.79 (0.75,0.84) |
| | | Prior history of COVID-19 | 0.73 (0.57,0.93) | 0.93 (0.74,1.16) | 0.87 (0.72,1.05) | 0.88 (0.76,1.03) | 0.81 (0.67,0.96) | 0.81 (0.62,1.06) |
| | | No prior history of event | 0.70 (0.65,0.75) | 0.84 (0.79,0.90) | 0.83 (0.79,0.88) | 0.83 (0.79,0.86) | 0.80 (0.76,0.84) | 0.76 (0.71,0.81) |
| | | Prior history of event | 0.77 (0.72,0.82) | 0.80 (0.75,0.86) | 0.82 (0.78,0.86) | 0.78 (0.74,0.81) | 0.74 (0.70,0.78) | 0.78 (0.72,0.84) |
| | | Sex: Female | 0.69 (0.64,0.75) | 0.79 (0.73,0.85) | 0.81 (0.76,0.86) | 0.77 (0.73,0.81) | 0.74 (0.69,0.78) | 0.75 (0.69,0.81) |
| | | Sex: Male | 0.76 (0.71,0.81) | 0.85 (0.80,0.90) | 0.84 (0.80,0.88) | 0.86 (0.83,0.90) | 0.89 (0.85,0.94) | 0.90 (0.84,0.97) |
| | Composite venous | Age group: <40 | 0.76 (0.56,1.03) | 0.92 (0.69,1.22) | 0.60 (0.46,0.77) | 0.74 (0.63,0.87) | 0.78 (0.66,0.94) | 0.76 (0.54,1.07) |
| | | Age group: 40-59 | 0.79 (0.62,1.02) | 0.93 (0.73,1.17) | 0.79 (0.65,0.95) | 0.77 (0.67,0.88) | 0.71 (0.62,0.83) | 0.67 (0.54,0.85) |
| | | Age group: 60-79 | 0.74 (0.62,0.88) | 0.73 (0.61,0.87) | 0.68 (0.58,0.79) | 0.64 (0.55,0.75) | 0.64 (0.54,0.76) | 0.61 (0.49,0.75) |
| | | Age group: >=80 | 0.61 (0.50,0.75) | 0.72 (0.60,0.87) | 0.78 (0.67,0.92) | 0.75 (0.65,0.85) | 0.71 (0.61,0.83) | 0.69 (0.56,0.86) |
| | | Ethnicity: White | 0.73 (0.65,0.82) | 0.84 (0.75,0.93) | 0.77 (0.71,0.84) | 0.78 (0.73,0.83) | 0.80 (0.74,0.87) | 0.77 (0.69,0.87) |
| | | Ethnicity: Black or Black British | 0.48 (0.19,1.18) | 0.89 (0.46,1.73) | 0.85 (0.51,1.40) | 0.64 (0.45,0.93) | 0.70 (0.49,1.01) | 0.42 (0.19,0.92) |
| | | Ethnicity: Asian or Asian British | 0.79 (0.42,1.46) | 0.33 (0.14,0.77) | 0.76 (0.49,1.19) | 0.68 (0.50,0.94) | 0.63 (0.44,0.89) | 0.65 (0.36,1.19) |
| | | Ethnicity: Other Ethnic Groups | 1.03 (0.37,2.88) | 0.16 (0.02,1.31) | 0.37 (0.13,1.08) | 0.64 (0.35,1.18) | 0.45 (0.21,0.97) | 0.60 (0.19,1.96) |
| | | Ethnicity: Mixed | 1.11 (0.32,3.84) | 2.02 (0.73,5.62) | 0.86 (0.33,2.25) | 0.78 (0.36,1.69) | 0.72 (0.31,1.72) | 0.53 (0.11,2.68) |
| | | Ethnicity: Unknown | 0.72 (0.18,2.82) | 0.91 (0.28,2.91) | 1.17 (0.56,2.46) | 0.86 (0.45,1.62) | 0.62 (0.27,1.43) | 0.72 (0.21,2.44) |
| | | No prior history of COVID-19 | 0.73 (0.66,0.82) | 0.82 (0.74,0.91) | 0.77 (0.71,0.84) | 0.78 (0.73,0.84) | 0.80 (0.74,0.86) | 0.77 (0.68,0.86) |
| | | Prior history of COVID-19 | 0.69 (0.43,1.09) | 0.88 (0.58,1.34) | 0.84 (0.61,1.16) | 0.63 (0.49,0.81) | 0.66 (0.50,0.86) | 0.84 (0.55,1.28) |
| | | No prior history of event | 0.74 (0.66,0.83) | 0.80 (0.72,0.90) | 0.76 (0.70,0.83) | 0.77 (0.72,0.82) | 0.76 (0.70,0.82) | 0.71 (0.63,0.80) |
| | | Prior history of event | 0.68 (0.51,0.89) | 0.92 (0.72,1.17) | 0.81 (0.66,0.99) | 0.72 (0.61,0.85) | 0.74 (0.62,0.88) | 0.82 (0.64,1.05) |
| | | Sex: Female | 0.72 (0.62,0.83) | 0.81 (0.71,0.93) | 0.76 (0.68,0.85) | 0.75 (0.69,0.82) | 0.74 (0.67,0.82) | 0.71 (0.62,0.82) |
| | | Sex: Male | 0.76 (0.65,0.89) | 0.83 (0.72,0.97) | 0.78 (0.69,0.89) | 0.81 (0.74,0.90) | 0.88 (0.79,0.99) | 0.87 (0.73,1.03) |
| **Doses 1&2: ChAdOx1** | Composite arterial | Age group: <40 | 0.56 (0.37,0.86) | 0.89 (0.63,1.28) | 0.63 (0.46,0.87) | 0.65 (0.49,0.85) | 0.54 (0.38,0.76) | 0.68 (0.43,1.07) |
| | | Age group: 40-59 | 0.70 (0.64,0.77) | 0.82 (0.75,0.89) | 0.80 (0.74,0.85) | 0.81 (0.76,0.86) | 0.81 (0.75,0.88) | 0.83 (0.74,0.92) |
| | | Age group: 60-79 | 0.73 (0.69,0.77) | 0.83 (0.78,0.88) | 0.79 (0.75,0.83) | 0.72 (0.69,0.75) | 0.68 (0.64,0.72) | 0.66 (0.61,0.71) |
| | | Age group: >=80 | 0.70 (0.64,0.77) | 0.77 (0.70,0.84) | 0.70 (0.65,0.75) | 0.69 (0.64,0.73) | 0.66 (0.62,0.72) | 0.67 (0.60,0.74) |
| | | Ethnicity: White | 0.71 (0.68,0.75) | 0.84 (0.80,0.87) | 0.79 (0.77,0.82) | 0.76 (0.74,0.79) | 0.74 (0.71,0.77) | 0.74 (0.70,0.78) |
| | | Ethnicity: Black or Black British | 0.75 (0.57,0.99) | 0.67 (0.50,0.89) | 0.72 (0.58,0.89) | 0.72 (0.61,0.85) | 0.65 (0.53,0.79) | 0.58 (0.43,0.79) |
| | | Ethnicity: Asian or Asian British | 0.71 (0.60,0.83) | 0.80 (0.69,0.94) | 0.80 (0.71,0.91) | 0.70 (0.63,0.78) | 0.65 (0.57,0.73) | 0.66 (0.55,0.78) |
| | | Ethnicity: Other Ethnic Groups | 0.56 (0.36,0.86) | 0.81 (0.56,1.17) | 0.64 (0.47,0.87) | 0.63 (0.49,0.82) | 0.73 (0.52,1.01) | 0.77 (0.50,1.20) |
| | | Ethnicity: Mixed | 1.09 (0.68,1.77) | 0.98 (0.60,1.59) | 1.09 (0.76,1.58) | 0.97 (0.70,1.36) | 0.89 (0.58,1.36) | 1.02 (0.57,1.83) |
| | | Ethnicity: Unknown | 0.90 (0.64,1.27) | 0.85 (0.59,1.21) | 0.84 (0.63,1.12) | 0.75 (0.57,0.98) | 0.72 (0.51,1.02) | 0.69 (0.43,1.10) |
| | | No prior history of COVID-19 | 0.72 (0.69,0.75) | 0.83 (0.80,0.87) | 0.79 (0.76,0.82) | 0.76 (0.74,0.78) | 0.73 (0.70,0.76) | 0.73 (0.69,0.77) |
| | | Prior history of COVID-19 | 0.71 (0.61,0.83) | 0.77 (0.66,0.90) | 0.80 (0.72,0.90) | 0.68 (0.61,0.75) | 0.68 (0.60,0.77) | 0.68 (0.57,0.81) |
| | | No prior history of event | 0.68 (0.64,0.72) | 0.82 (0.78,0.87) | 0.83 (0.79,0.86) | 0.74 (0.72,0.77) | 0.68 (0.65,0.71) | 0.63 (0.60,0.68) |
| | | Prior history of event | 0.74 (0.70,0.79) | 0.81 (0.76,0.86) | 0.71 (0.68,0.75) | 0.72 (0.69,0.75) | 0.71 (0.68,0.75) | 0.76 (0.71,0.82) |
| | | Sex: Female | 0.70 (0.66,0.75) | 0.82 (0.77,0.88) | 0.77 (0.73,0.81) | 0.75 (0.71,0.79) | 0.73 (0.69,0.78) | 0.75 (0.69,0.81) |
| | | Sex: Male | 0.74 (0.70,0.78) | 0.85 (0.81,0.89) | 0.82 (0.79,0.86) | 0.79 (0.76,0.82) | 0.77 (0.73,0.81) | 0.77 (0.72,0.83) |
| | Composite venous | Age group: <40 | 0.60 (0.41,0.88) | 0.89 (0.64,1.22) | 0.59 (0.43,0.80) | 0.49 (0.39,0.61) | 0.48 (0.37,0.62) | 0.44 (0.30,0.66) |
| | | Age group: 40-59 | 0.71 (0.60,0.83) | 0.86 (0.74,1.01) | 0.85 (0.75,0.96) | 0.70 (0.63,0.79) | 0.73 (0.64,0.84) | 0.71 (0.59,0.85) |
| | | Age group: 60-79 | 0.69 (0.61,0.79) | 0.75 (0.66,0.85) | 0.68 (0.61,0.76) | 0.61 (0.55,0.67) | 0.52 (0.46,0.59) | 0.55 (0.47,0.64) |
| | | Age group: >=80 | 0.85 (0.68,1.06) | 0.89 (0.72,1.11) | 0.82 (0.68,0.99) | 0.68 (0.58,0.80) | 0.62 (0.52,0.74) | 0.60 (0.47,0.77) |
| | | Ethnicity: White | 0.73 (0.67,0.80) | 0.84 (0.77,0.92) | 0.79 (0.73,0.84) | 0.70 (0.66,0.75) | 0.67 (0.61,0.72) | 0.68 (0.61,0.76) |
| | | Ethnicity: Black or Black British | 0.60 (0.31,1.14) | 0.71 (0.39,1.30) | 0.89 (0.58,1.37) | 0.83 (0.59,1.15) | 0.75 (0.51,1.10) | 0.45 (0.24,0.83) |
| | | Ethnicity: Asian or Asian British | 0.56 (0.30,1.04) | 0.77 (0.44,1.34) | 0.95 (0.64,1.40) | 0.67 (0.48,0.95) | 0.55 (0.36,0.86) | 0.67 (0.36,1.25) |
| | | Ethnicity: Other Ethnic Groups | 0.26 (0.06,1.12) | 1.20 (0.55,2.60) | 0.85 (0.42,1.72) | 0.79 (0.43,1.45) | 0.72 (0.33,1.57) | 0.81 (0.24,2.75) |
| | | Ethnicity: Mixed | 1.37 (0.59,3.21) | 0.49 (0.14,1.69) | 0.56 (0.24,1.30) | 0.47 (0.22,1.02) | 0.54 (0.23,1.31) | 0.73 (0.22,2.48) |
| | | Ethnicity: Unknown | 0.73 (0.33,1.62) | 1.10 (0.55,2.19) | 0.94 (0.52,1.69) | 0.95 (0.56,1.64) | 0.74 (0.37,1.46) | 0.83 (0.34,2.03) |
| | | No prior history of COVID-19 | 0.71 (0.65,0.78) | 0.82 (0.75,0.90) | 0.80 (0.75,0.86) | 0.70 (0.66,0.74) | 0.65 (0.60,0.71) | 0.67 (0.60,0.74) |
| | | Prior history of COVID-19 | 0.90 (0.67,1.20) | 1.08 (0.82,1.43) | 0.71 (0.55,0.92) | 0.81 (0.66,0.99) | 0.85 (0.66,1.10) | 0.84 (0.58,1.21) |
| | | No prior history of event | 0.73 (0.67,0.80) | 0.83 (0.76,0.91) | 0.78 (0.73,0.84) | 0.70 (0.65,0.74) | 0.65 (0.60,0.70) | 0.64 (0.57,0.71) |
| | | Prior history of event | 0.65 (0.52,0.82) | 0.87 (0.71,1.06) | 0.79 (0.68,0.93) | 0.66 (0.58,0.76) | 0.63 (0.54,0.74) | 0.72 (0.58,0.91) |
| | | Sex: Female | 0.71 (0.63,0.81) | 0.80 (0.71,0.90) | 0.78 (0.71,0.86) | 0.68 (0.62,0.74) | 0.64 (0.58,0.71) | 0.65 (0.57,0.75) |
| | | Sex: Male | 0.74 (0.65,0.83) | 0.88 (0.79,0.99) | 0.81 (0.74,0.89) | 0.75 (0.69,0.81) | 0.72 (0.65,0.81) | 0.75 (0.65,0.87) |

Supplementary Table 28: Subgroup-specific maximally-adjusted hazard ratios and 95% CIs for composite arterial and composite venous events by time since booster vaccination with BNT-162b2 or mRNA-1273for those on a primary course of ChAdOx1.

| Vaccination course | Event | Stratum | Week 1 | Week 2 | Weeks 3-4 | Weeks 5-12 | Weeks 13-24 |
|---|---|---|---|---|---|---|---|
| | | | | **Weeks 1-4** | | | **Weeks 5-24** |
| **Doses 1&2: ChAdOx1; Booster: BNT-162b2** | Composite arterial | Age group: <40 | 0.88 (0.50, 1.57) | 0.52 (0.26, 1.07) | 0.53 (0.31, 0.90) | 0.50 (0.31, 0.80) | 1.01 (0.30, 3.36) |
| | | Age group: 40-59 | 0.70 (0.62, 0.79) | 0.79 (0.70, 0.89) | 0.83 (0.75, 0.91) | 0.82 (0.75, 0.90) | 0.85 (0.64, 1.12) |
| | | Age group: 60-79 | 0.79 (0.74, 0.84) | 0.81 (0.76, 0.86) | 0.80 (0.76, 0.84) | 0.73 (0.70, 0.77) | 0.65 (0.59, 0.72) |
| | | Age group: >=80 | 0.78 (0.70, 0.86) | 0.79 (0.71, 0.88) | 0.80 (0.73, 0.86) | 0.75 (0.70, 0.80) | 0.70 (0.62, 0.80) |
| | | Ethnicity: White | 0.78 (0.74, 0.83) | 0.82 (0.77, 0.86) | 0.82 (0.79, 0.85) | 0.78 (0.75, 0.80) | 0.69 (0.64, 0.75) |
| | | Ethnicity: Black or Black British | 0.96 (0.65, 1.41) | 0.90 (0.60, 1.35) | 0.91 (0.67, 1.24) | 0.82 (0.63, 1.07) | 1.12 (0.61, 2.05) |
| | | Ethnicity: Asian or Asian British | 0.78 (0.63, 0.97) | 0.89 (0.72, 1.09) | 0.87 (0.74, 1.02) | 0.75 (0.65, 0.85) | 0.80 (0.58, 1.11) |
| | | Ethnicity: Other Ethnic Groups | 0.81 (0.50, 1.32) | 0.80 (0.49, 1.30) | 0.78 (0.53, 1.14) | 0.67 (0.49, 0.92) | 0.51 (0.21, 1.26) |
| | | Ethnicity: Mixed | 0.63 (0.29, 1.38) | 0.46 (0.18, 1.15) | 1.00 (0.61, 1.63) | 0.70 (0.43, 1.14) | 0.56 (0.16, 1.99) |
| | | Ethnicity: Unknown | 0.57 (0.35, 0.94) | 0.50 (0.29, 0.87) | 0.69 (0.48, 0.99) | 0.70 (0.51, 0.97) | 0.72 (0.33, 1.57) |
| | | No prior history of COVID-19 | 0.78 (0.74,0.82) | 0.82 (0.78,0.86) | 0.83 (0.79,0.86) | 0.78 (0.75,0.81) | 0.71 (0.65,0.76) |
| | | Prior history of COVID-19 | 0.90 (0.75,1.08) | 0.81 (0.67,0.99) | 0.83 (0.72,0.96) | 0.76 (0.67,0.86) | 0.72 (0.54,0.95) |
| | | No prior history of event | 0.70 (0.66,0.75) | 0.78 (0.74,0.84) | 0.76 (0.72,0.80) | 0.71 (0.68,0.74) | 0.63 (0.57,0.69) |
| | | Prior history of event | 0.89 (0.82,0.95) | 0.84 (0.78,0.91) | 0.89 (0.84,0.95) | 0.85 (0.81,0.89) | 0.80 (0.72,0.90) |
| | | Sex: Female | 0.83 (0.77,0.90) | 0.80 (0.74,0.87) | 0.85 (0.80,0.90) | 0.80 (0.76,0.85) | 0.70 (0.62,0.79) |
| | | Sex: Male | 0.77 (0.72,0.82) | 0.85 (0.80,0.90) | 0.84 (0.80,0.88) | 0.81 (0.77,0.84) | 0.76 (0.69,0.84) |
| | Composite venous | Age group: <40 | 0.81 (0.46, 1.41) | 1.15 (0.71, 1.88) | 0.96 (0.65, 1.41) | 0.68 (0.47, 0.99) | 1.24 (0.48, 3.17) |
| | | Age group: 40-59 | 0.71 (0.57, 0.89) | 0.91 (0.75, 1.12) | 0.75 (0.63, 0.89) | 0.79 (0.68, 0.92) | 0.57 (0.33, 0.97) |
| | | Age group: 60-79 | 0.62 (0.53, 0.71) | 0.62 (0.54, 0.71) | 0.64 (0.58, 0.72) | 0.53 (0.48, 0.59) | 0.58 (0.47, 0.72) |
| | | Age group: >=80 | 0.55 (0.42, 0.73) | 0.59 (0.45, 0.77) | 0.56 (0.46, 0.69) | 0.53 (0.45, 0.62) | 0.46 (0.33, 0.65) |
| | | Ethnicity: White | 0.68 (0.61, 0.76) | 0.74 (0.67, 0.83) | 0.72 (0.67, 0.79) | 0.65 (0.60, 0.70) | 0.64 (0.55, 0.76) |
| | | Ethnicity: Black or Black British | | 0.69 (0.44, 1.09) | | | 0.49 (0.29, 0.82) |
| | | Ethnicity: Asian or Asian British | 0.37 (0.14, 1.00) | 1.15 (0.62, 2.16) | 0.72 (0.40, 1.28) | 0.75 (0.48, 1.20) | 0.51 (0.11, 2.45) |
| | | Ethnicity: Other Ethnic Groups | | 0.78 (0.33, 1.82) | | | 0.89 (0.36, 2.20) |
| | | Ethnicity: Mixed | 1.14 (0.28, 4.62) | 1.35 (0.35, 5.11) | 1.75 (0.80, 3.83) | 0.32 (0.12, 0.85) | 0.84 (0.14, 5.11) |
| | | Ethnicity: | 0.22 (0.05, 0.92) | 0.43 (0.15, 1.19) | 0.42 (0.20, 0.88) | 0.40 (0.21, 0.74) | 0.81 (0.26, 2.49) |
| | | No prior history of COVID-19 | 0.66 (0.59,0.74) | 0.72 (0.65,0.80) | 0.71 (0.66,0.77) | 0.64 (0.59,0.69) | 0.62 (0.52,0.73) |
| | | Prior history of COVID-19 | 0.84 (0.57,1.25) | 1.03 (0.72,1.48) | 0.87 (0.64,1.17) | 0.61 (0.47,0.81) | 0.66 (0.35,1.23) |
| | | No prior history of event | 0.67 (0.60,0.75) | 0.72 (0.64,0.80) | 0.72 (0.66,0.79) | 0.62 (0.57,0.67) | 0.59 (0.50,0.70) |
| | | Prior history of event | 0.63 (0.47,0.85) | 0.86 (0.66,1.12) | 0.68 (0.55,0.85) | 0.70 (0.58,0.83) | 0.74 (0.49,1.11) |
| | | Sex: Female | 0.74 (0.64,0.85) | 0.77 (0.67,0.89) | 0.76 (0.68,0.84) | 0.66 (0.60,0.73) | 0.54 (0.43,0.67) |
| | | Sex: Male | 0.62 (0.52,0.72) | 0.73 (0.63,0.85) | 0.71 (0.64,0.80) | 0.66 (0.59,0.73) | 0.81 (0.65,1.01) |
| **Doses 1&2: ChAdOx1; Booster: mRNA-1273** | Composite arterial | Age group: <40 | | 0.72 (0.40, 1.27) | | | 0.46 (0.19, 1.10) |
| | | Age group: 40-59 | | 0.84 (0.76, 0.94) | | | 0.73 (0.63,0.85) |
| | | Age group: 60-79 | | 0.72 (0.66, 0.79) | | | 0.60 (0.54, 0.67) |
| | | Age group: >=80 | | 0.80 (0.65, 0.99) | | | 0.88 (0.70, 1.10) |
| | | Ethnicity: White | | 0.76 (0.71, 0.81) | | | 0.66 (0.61, 0.72) |
| | | Ethnicity: Black or Black British | | 0.72 (0.42, 1.21) | | | 0.51 (0.24, 1.07) |
| | | Ethnicity: Asian or Asian British | | 0.74 (0.57, 0.96) | | | 0.81 (0.59, 1.11) |
| | | Ethnicity: Other Ethnic Groups | | 1.07 (0.64, 1.79) | | | 0.75 (0.37, 1.54) |
| | | Ethnicity: Mixed | | 0.57 (0.24, 1.34) | | | 0.17 (0.04, 0.78) |
| | | Ethnicity: Unknown | | 0.81 (0.54, 1.23) | | | 0.44 (0.24, 0.81) |
| | | No prior history of COVID-19 | | 0.76 (0.71, 0.81) | | | 0.67 (0.61, 0.72) |
| | | Prior history of COVID-19 | | 0.86 (0.68, 1.10) | | | 0.62 (0.45, 0.86) |
| | | No prior history of event | | 0.71 (0.66, 0.77) | | | 0.61 (0.56, 0.67) |
| | | Prior history of event | | 0.88 (0.79, 0.98) | | | 0.74 (0.64, 0.84) |
| | | Sex: Female | | 0.76 (0.68, 0.84) | | | 0.65 (0.56, 0.74) |
| | | Sex: Male | | 0.78 (0.72, 0.84) | | | 0.68 (0.62, 0.75) |
| | Composite venous | Age group: <40 | | 0.67 (0.41, 1.11) | | | 0.69 (0.36, 1.33) |
| | | Age group: 40-59 | | 0.63 (0.52, 0.77) | | | 0.64 (0.50, 0.82) |
| | | Age group: 60-79 | | 0.51 (0.42, 0.61) | | | 0.40 (0.32, 0.50) |
| | | Age group: >=80 | | 0.49 (0.27, 0.88) | | | 0.81 (0.46, 1.42) |
| | | Ethnicity: White | | 0.58 (0.51, 0.66) | | | 0.54 (0.46, 0.63) |
| | | Ethnicity: Black or Black British | | 0.61 (0.24, 1.58) | | | 0.52 (0.15, 1.80) |
| | | Ethnicity: Asian or Asian British | | 0.31 (0.10, 1.01) | | | 0.37 (0.09, 1.62) |
| | | Ethnicity: Other Ethnic Groups | | 0.97 (0.23, 4.03) | | | 2.05 (0.44, 9.59) |
| | | Ethnicity: Mixed | | 1.78 (0.54, 5.83) | | | 1.16 (0.19, 7.08) |
| | | Ethnicity: Unknown | | 0.35 (0.13, 0.94) | | | 0.26 (0.08, 0.78) |
| | | No prior history of COVID-19 | | 0.58 (0.51,0.65) | | | 0.56 (0.48,0.66) |
| | | Prior history of COVID-19 | | 0.75 (0.48,1.18) | | | 0.45 (0.22,0.91) |
| | | No prior history of event | | 0.55 (0.48,0.62) | | | 0.51 (0.43,0.61) |
| | | Prior history of event | | 0.83 (0.61,1.15) | | | 0.75 (0.50,1.12) |
| | | Sex: Female | | 0.54 (0.45,0.65) | | | 0.55 (0.44,0.69) |
| | | Sex: Male | | 0.64 (0.55,0.76) | | | 0.58 (0.47,0.72) |

Supplementary Table 29: Subgroup-specific maximally-adjusted hazard ratios and 95% CIs for composite arterial and composite venous events by time since booster with BNT-162b2 or mRNA-1273 for those on a primary course of BNT-162b2.

| Vaccination course | Event | Stratum | Week 1 | Week 2 | Weeks 3-4 | Weeks 5-12 | Weeks 13-24 |
|---|---|---|---|---|---|---|---|
| | | | | Weeks 1-4 | | Weeks 5-24 | |
| **Doses 1&2: BNT-162b2; Booster: BNT-162b2** | Composite arterial | Age group: <40 | 0.71 (0.40, 1.29) | 1.02 (0.60, 1.75) | 0.71 (0.45, 1.12) | 1.17 (0.79, 1.73) | 1.04 (0.44, 2.45) |
| | | Age group: 40-59 | 0.67 (0.56, 0.80) | 0.78 (0.66, 0.92) | 0.85 (0.75, 0.96) | 0.79 (0.71, 0.87) | 0.70 (0.56, 0.88) |
| | | Age group: 60-79 | 0.67 (0.62, 0.73) | 0.79 (0.73, 0.86) | 0.77 (0.72, 0.82) | 0.73 (0.69, 0.78) | 0.68 (0.61, 0.75) |
| | | Age group: >=80 | 0.65 (0.60, 0.70) | 0.69 (0.64, 0.75) | 0.71 (0.67, 0.76) | 0.68 (0.64, 0.72) | 0.63 (0.58, 0.68) |
| | | Ethnicity: White | 0.68 (0.64, 0.72) | 0.79 (0.74, 0.83) | 0.79 (0.76, 0.83) | 0.77 (0.74, 0.80) | 0.72 (0.68, 0.76) |
| | | Ethnicity: Black or Black British | 0.96 (0.63, 1.48) | 0.86 (0.55, 1.34) | 0.73 (0.51, 1.05) | 0.75 (0.58, 0.98) | 1.03 (0.64, 1.66) |
| | | Ethnicity: Asian or Asian British | 0.75 (0.59, 0.96) | 0.72 (0.56, 0.93) | 0.83 (0.69, 1.00) | 0.88 (0.76, 1.01) | 0.71 (0.54, 0.92) |
| | | Ethnicity: Other Ethnic Groups | 0.75 (0.42, 1.33) | 0.61 (0.33, 1.15) | 0.77 (0.49, 1.20) | 0.90 (0.64, 1.25) | 0.85 (0.45, 1.61) |
| | | Ethnicity: Mixed | 0.50 (0.15, 1.70) | 0.60 (0.22, 1.66) | 1.36 (0.76, 2.44) | 1.00 (0.57, 1.74) | 2.04 (0.90, 4.64) |
| | | Ethnicity: Unknown | 0.59 (0.32, 1.09) | 0.65 (0.36, 1.17) | 0.81 (0.53, 1.23) | 0.59 (0.41, 0.84) | 0.46 (0.24, 0.86) |
| | | No prior history of COVID-19 | 0.69 (0.65, 0.73) | 0.78 (0.74, 0.83) | 0.79 (0.76, 0.83) | 0.78 (0.75, 0.81) | 0.73 (0.69, 0.77) |
| | | Prior history of COVID-19 | 0.83 (0.64, 1.08) | 0.82 (0.63, 1.08) | 1.02 (0.84, 1.23) | 0.87 (0.74, 1.03) | 0.80 (0.59, 1.07) |
| | | No prior history of event | 0.62 (0.58, 0.67) | 0.72 (0.68, 0.78) | 0.76 (0.72, 0.80) | 0.73 (0.69, 0.76) | 0.65 (0.60, 0.70) |
| | | Prior history of event | 0.77 (0.71, 0.83) | 0.85 (0.78, 0.91) | 0.84 (0.79, 0.89) | 0.82 (0.78, 0.87) | 0.83 (0.76, 0.91) |
| | | Sex: Female | 0.67 (0.62, 0.73) | 0.76 (0.70, 0.82) | 0.81 (0.76, 0.86) | 0.76 (0.72, 0.80) | 0.71 (0.65, 0.78) |
| | | Sex: Male | 0.72 (0.67, 0.77) | 0.83 (0.77, 0.89) | 0.82 (0.78, 0.87) | 0.84 (0.80, 0.88) | 0.79 (0.73, 0.86) |
| | Composite venous | Age group: <40 | 0.61 (0.35, 1.09) | 1.01 (0.64, 1.59) | 0.90 (0.62, 1.30) | 0.99 (0.71, 1.37) | 0.74 (0.32, 1.75) |
| | | Age group: 40-59 | 0.85 (0.63, 1.14) | 0.65 (0.47, 0.91) | 0.72 (0.57, 0.92) | 0.66 (0.54, 0.79) | 0.60 (0.42, 0.86) |
| | | Age group: 60-79 | 0.61 (0.51, 0.73) | 0.56 (0.47, 0.68) | 0.54 (0.47, 0.62) | 0.46 (0.40, 0.52) | 0.48 (0.38, 0.59) |
| | | Age group: >=80 | 0.44 (0.36, 0.55) | 0.54 (0.44, 0.65) | 0.46 (0.40, 0.54) | 0.42 (0.37, 0.48) | 0.35 (0.29, 0.43) |
| | | Ethnicity: White | 0.63 (0.56, 0.72) | 0.65 (0.58, 0.73) | 0.65 (0.59, 0.71) | 0.60 (0.56, 0.65) | 0.55 (0.49, 0.63) |
| | | Ethnicity: Black or Black British | 0.45 (0.14, 1.44) | 0.49 (0.15, 1.60) | 0.57 (0.26, 1.24) | 0.81 (0.47, 1.40) | 0.41 (0.12, 1.39) |
| | | Ethnicity: Asian or Asian British | 0.45 (0.16, 1.22) | 0.44 (0.16, 1.22) | 0.60 (0.31, 1.16) | 0.55 (0.34, 0.89) | 0.52 (0.20, 1.35) |
| | | Ethnicity: Other Ethnic Groups | | 0.48 (0.16, 1.45) | | 0.94 (0.40, 2.22) | |
| | | Ethnicity: Mixed | 1.62 (0.30, 8.84) | 1.81 (0.34, 9.72) | 1.42 (0.34, 5.85) | 0.84 (0.24, 2.94) | 0.96 (0.09, 10.43) |
| | | Ethnicity: Unknown | 0.46 (0.14, 1.56) | 1.13 (0.45, 2.79) | 0.17 (0.04, 0.72) | 0.44 (0.21, 0.94) | 0.21 (0.04, 1.22) |
| | | No prior history of COVID-19 | 0.62 (0.55, 0.70) | 0.66 (0.58, 0.74) | 0.64 (0.58, 0.70) | 0.60 (0.56, 0.65) | 0.54 (0.48, 0.62) |
| | | Prior history of COVID-19 | 0.81 (0.48, 1.39) | 0.65 (0.36, 1.17) | 0.67 (0.43, 1.05) | 0.70 (0.51, 0.97) | 0.63 (0.33, 1.19) |
| | | No prior history of event | 0.60 (0.53, 0.69) | 0.65 (0.57, 0.74) | 0.62 (0.57, 0.69) | 0.59 (0.55, 0.64) | 0.53 (0.46, 0.60) |
| | | Prior history of event | 0.72 (0.54, 0.96) | 0.66 (0.49, 0.89) | 0.70 (0.56, 0.88) | 0.64 (0.53, 0.77) | 0.66 (0.47, 0.92) |
| | | Sex: Female | 0.58 (0.50, 0.69) | 0.60 (0.51, 0.70) | 0.60 (0.53, 0.67) | 0.58 (0.53, 0.64) | 0.57 (0.48, 0.68) |
| | | Sex: Male | 0.68 (0.57, 0.81) | 0.74 (0.62, 0.88) | 0.70 (0.61, 0.80) | 0.64 (0.58, 0.72) | 0.53 (0.44, 0.65) |
| **Doses 1&2: BNT-162b2; Booster: mRNA-1273** | Composite arterial | Age group: <40 | | 0.38 (0.21, 0.69) | | 0.19 (0.07, 0.53) | |
| | | Age group: 40-59 | 0.63 (0.42, 0.94) | 0.96 (0.68, 1.34) | 0.83 (0.63,1.09) | 0.79 (0.61, 1.01) | 13.78 (1.83,103.78) |
| | | Age group: 60-79 | | 0.70 (0.61, 0.82) | | 0.65 (0.55, 0.78) | |
| | | Age group: >=80 | 0.81 (0.54, 1.22) | 0.95 (0.65, 1.40) | 0.68 (0.49, 0.96) | 0.67 (0.51, 0.87) | 0.94 (0.23, 3.85) |
| | | Ethnicity: White | 0.68 (0.55, 0.84) | 0.93 (0.77, 1.12) | 0.88 (0.76, 1.02) | 0.85 (0.75, 0.96) | 1.19 (0.37, 3.80) |
| | | Ethnicity: Black or Black British | | 1.07 (0.50, 2.28) | | 1.33 (0.57, 3.10) | |
| | | Ethnicity: Asian or Asian British | | 0.83 (0.53, 1.32) | | 0.70 (0.38, 1.30) | |
| | | Ethnicity: Other Ethnic Groups | | 1.31 (0.53, 3.25) | | 1.68 (0.57, 4.90) | |
| | | Ethnicity: Mixed | | 2.13 (0.74, 6.14) | | 1.28 (0.26, 6.24) | |
| | | Ethnicity: Unknown | | 0.54 (0.19, 1.56) | | 0.11 (0.01, 0.96) | |
| | | No prior history of COVID-19 | 0.68 (0.55,0.83) | 0.96 (0.80,1.15) | 0.88 (0.76,1.01) | 0.85 (0.75,0.96) | 1.12 (0.35,3.59) |
| | | Prior history of COVID-19 | | 1.21 (0.79, 1.86) | | 0.96 (0.53, 1.75) | |
| | | No prior history of event | 0.59 (0.45, 0.78) | 0.86 (0.68, 1.09) | 0.82 (0.69, 0.98) | 0.72 (0.62, 0.85) | 1.21 (0.38, 3.91) |
| | | Prior history of event | | 0.83 (0.71, 0.96) | | 0.81 (0.68, 0.97) | |
| | | Sex: Female | 0.62 (0.44,0.87) | 0.58 (0.41,0.83) | 0.80 (0.63,1.00) | 0.67 (0.54,0.82) | 1.30 (0.30,5.51) |
| | | Sex: Male | 0.69 (0.54,0.89) | 1.20 (0.99,1.47) | 0.93 (0.78,1.10) | 0.97 (0.83,1.13) | 0.80 (0.11,5.80) |
| | Composite venous | Age group: <40 | | 0.94 (0.61, 1.45) | | 0.85 (0.37, 1.94) | |
| | | Age group: 40-59 | | 0.60 (0.39, 0.93) | | 0.57 (0.35, 0.93) | |
| | | Age group: 60-79 | | 0.39 (0.28, 0.56) | | 0.44 (0.30, 0.65) | |
| | | Age group: >=80 | | 0.52 (0.30, 0.90) | | 0.31 (0.15, 0.65) | |
| | | Ethnicity: White | | 0.60 (0.48, 0.75) | | 0.58 (0.45, 0.76) | |
| | | Ethnicity: Black or Black British | | 0.32 (0.04, 2.48) | | 0.77 (0.17, 3.42) | |
| | | Ethnicity: Asian or Asian British | | 0.83 (0.25, 2.78) | | 0.40 (0.06, 2.68) | |
| | | Ethnicity: Other Ethnic Groups | | 0.92 (0.18, 4.69) | | 0.58 (0.05, 6.45) | |
| | | Ethnicity: Mixed | | 0.61 (0.49, 0.75) | | 0.57 (0.44, 0.73) | |
| | | Ethnicity: Unknown | | 0.39 (0.12, 1.28) | | 0.21 (0.03, 1.47) | |
| | | No prior history of COVID-19 | | 0.61 (0.49, 0.76) | | 0.54 (0.42, 0.71) | |
| | | Prior history of COVID-19 | | 0.54 (0.31, 0.95) | | 0.54 (0.27, 1.08) | |
| | | No prior history of event | | 0.62 (0.47, 0.81) | | 0.67 (0.48, 0.93) | |
| | | Prior history of event | | 0.61 (0.44, 0.83) | | 0.48 (0.32, 0.71) | |
| | | Sex: Female | | 0.94 (0.61, 1.45) | | 0.85 (0.37, 1.94) | |
| | | Sex: Male | | 0.60 (0.39, 0.93) | | 0.57 (0.35, 0.93) | |

* Stratified analyses for primary course mRNA-1273 are not presented, nor are booster with mRNA-1273 only, due to the small number of individuals who received mRNA-1273.

Supplementary Table 30: Associations of COVID-19 vaccination brand and dose with myocarditis and pericarditis until the CDC's Announcement on 17 May 2021.

| Event | Vaccination course | Week 1 | Week 2 | Weeks 3-4 | Weeks 5-12 | Weeks 13-24 |
|---|---|---|---|---|---|---|
| | | | | | Week 5-24 | |
| **Myocarditis** | **Dose 1: ChAdOx1** | 1.42 (0.83, 2.45) | 0.53 (0.23, 1.21) | 1.13 (0.72, 1.76) | 0.77 (0.52, 1.12) | 0.69 (0.32, 1.46) |
| | **Dose 1: BNT-162b2** | 1.85 (0.92, 3.72) | 1.20 (0.52, 2.74) | 0.87 (0.43, 1.76) | 0.74 (0.47, 1.17) | 0.74 (0.38, 1.46) |
| | **Doses 1&2: ChAdOx1** | 0.55 (0.14, 2.20) | 0.64 (0.15, 2.73) | 0.76 (0.22, 2.55) | 1.15 (0.23, 5.77) | |
| | **Doses 1&2: BNT-162b2** | 0.95 (0.27, 3.38) | 1.41 (0.42, 4.76) | 0.60 (0.17, 2.08) | 1.21 (0.41, 3.54) | |
| **Pericarditis** | **Dose 1: ChAdOx1** | 0.88 (0.44, 1.74) | 1.57 (0.92, 2.68) | 1.21 (0.76, 1.93) | 1.08 (0.76, 1.54) | 0.65 (0.31, 1.34) |
| | **Dose 1: BNT-162b2** | 0.43 (0.11, 1.78) | 0.95 (0.34, 2.64) | 1.19 (0.62, 2.30) | 1.15 (0.75, 1.76) | 1.89 (1.13, 3.16) |
| | **Doses 1&2: ChAdOx1** | 1.18 (0.51, 2.70) | 1.19 (0.48, 2.93) | 0.57 (0.22, 1.47) | 0.41 (0.09, 1.82) | |
| | **Doses 1&2: BNT-162b2** | 0.31 (0.07, 1.33) | 1.23 (0.48, 3.12) | 1.35 (0.62, 2.97) | 1.34 (0.63, 2.89) | |

### Supplementary Figures

#### Supplementary Figure 1: Consort Flow Diagram for Population Inclusion/Exclusion Criteria*

*Counts rounded to the nearest 5. Number excluded at each stage calculated from the rounded counts. 14,550 excluded due to having a latest non-NULL LSOA recorded as non-English, although latest LSOAs assigned as NULL (due to having no latest non-NULL LSOA on record, or due to having conflicting latest LSOAs) were included.

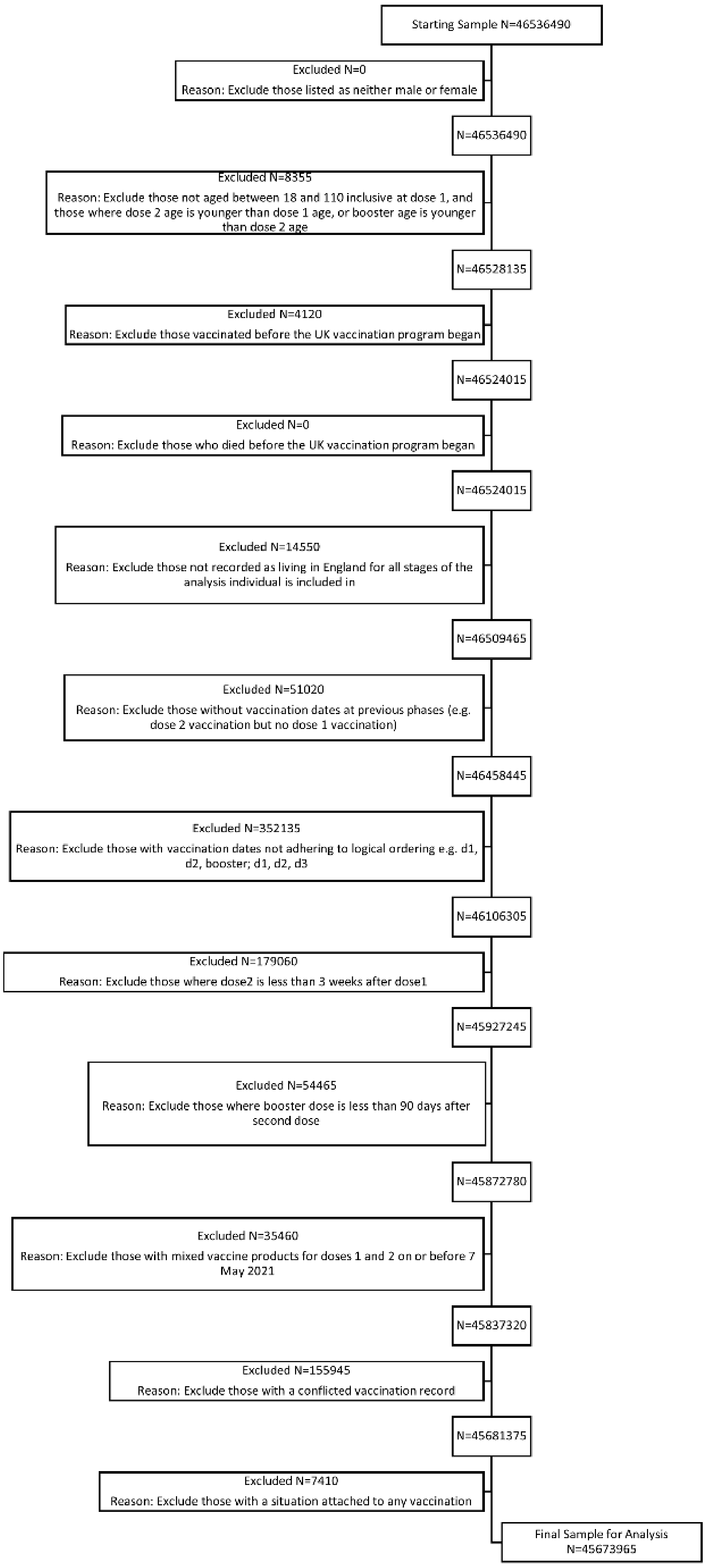

Supplementary Figure 2: Subgroup-specific maximally-adjusted hazard ratios for a composite arterial event by time since dose 1 vaccination

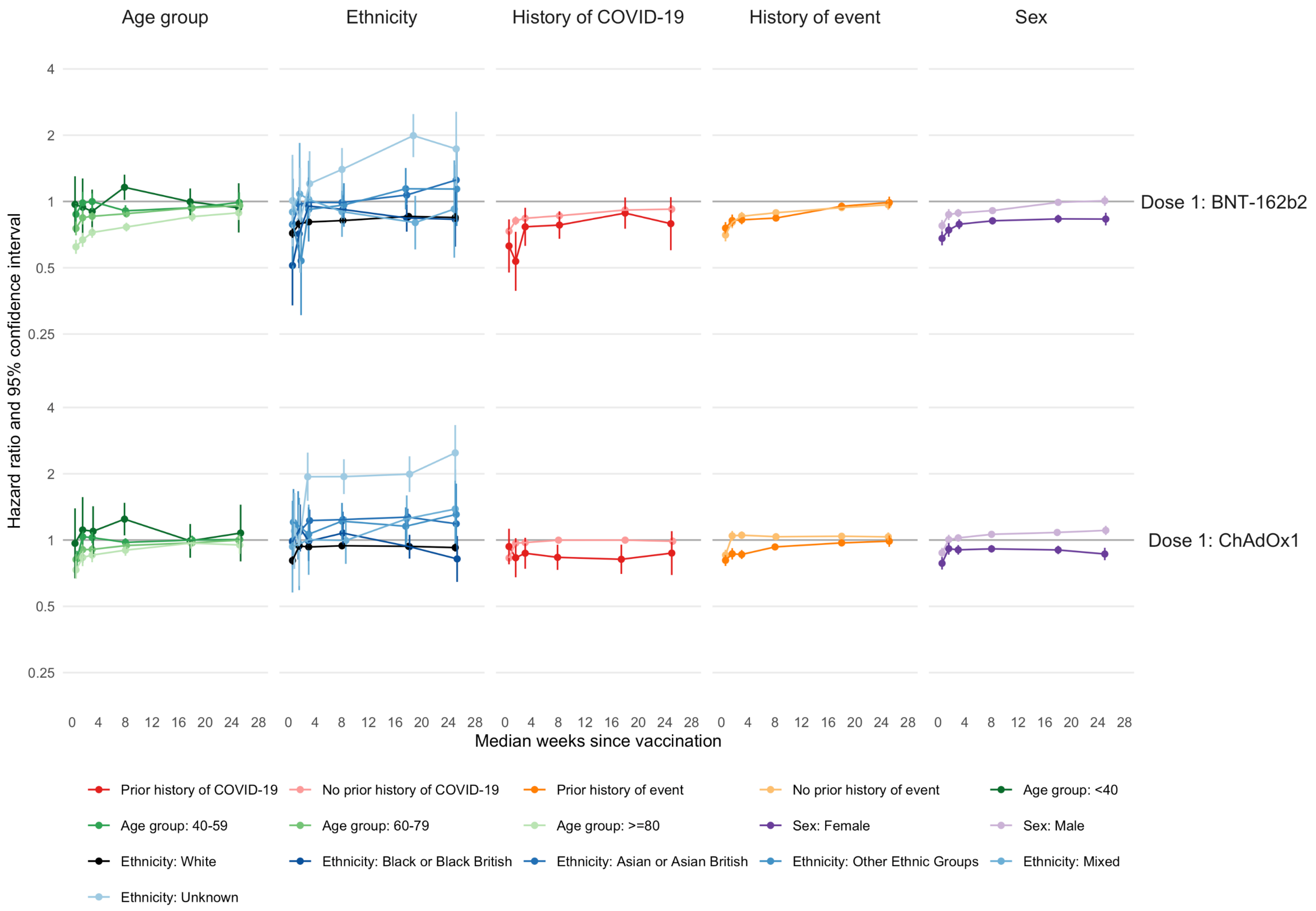

Supplementary Figure 3: Subgroup-specific maximally-adjusted hazard ratios for a composite venous event by time since dose 1 vaccination

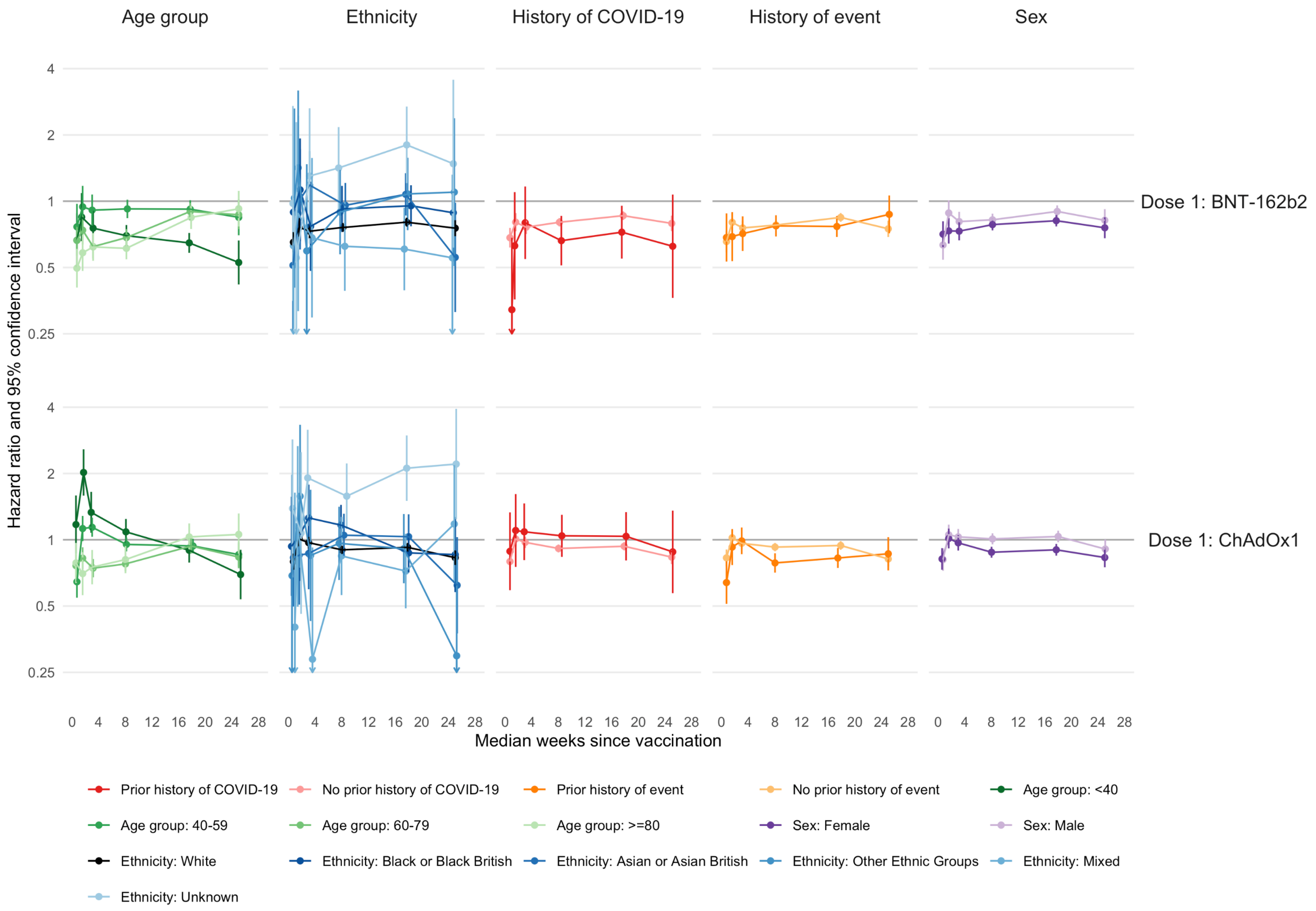

Supplementary Figure 4: Subgroup-specific maximally-adjusted hazard ratios for a composite arterial event by time since second dose vaccination

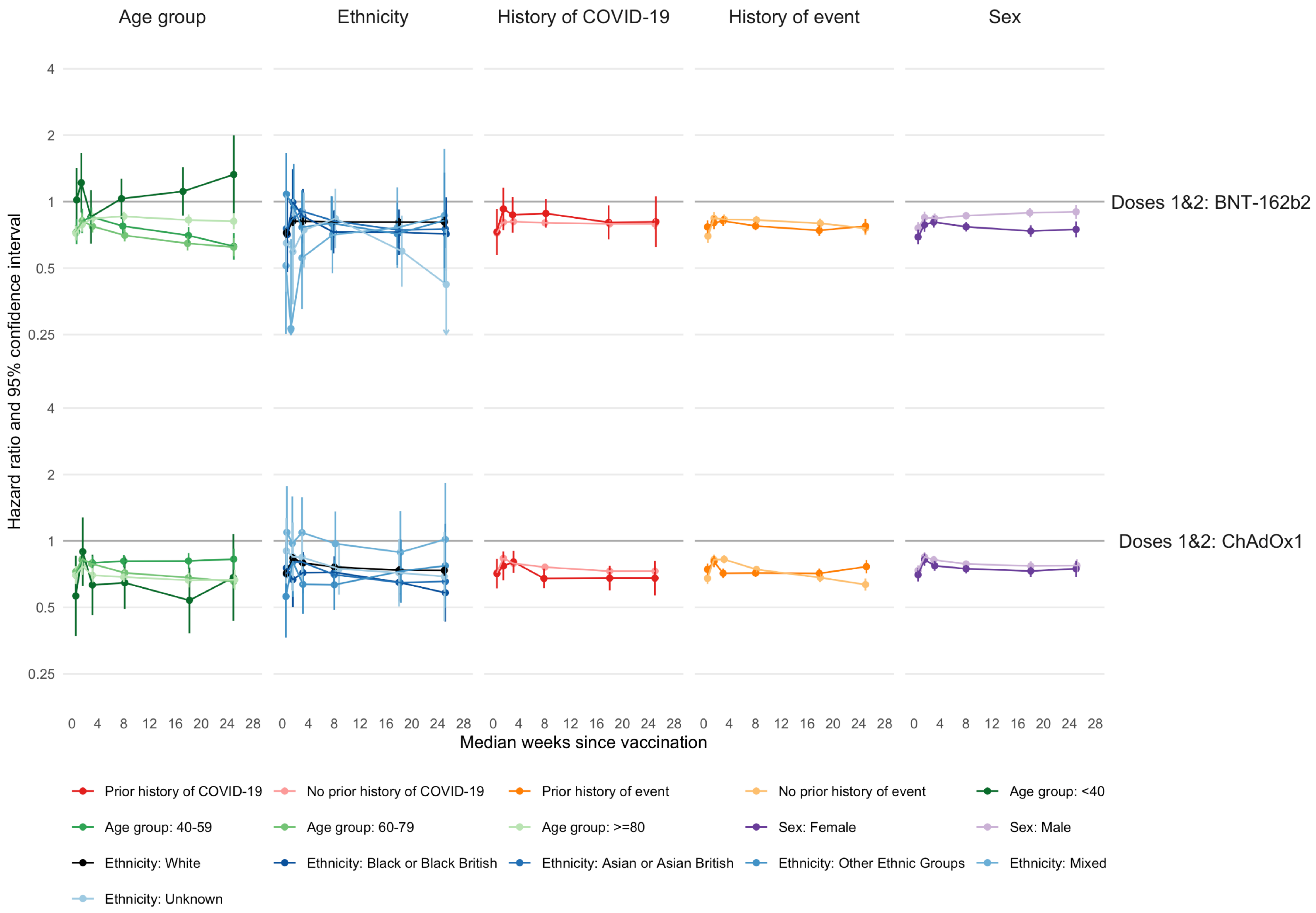

Supplementary Figure 5: Subgroup-specific maximally-adjusted hazard ratios for a composite venous event by time since second dose vaccination

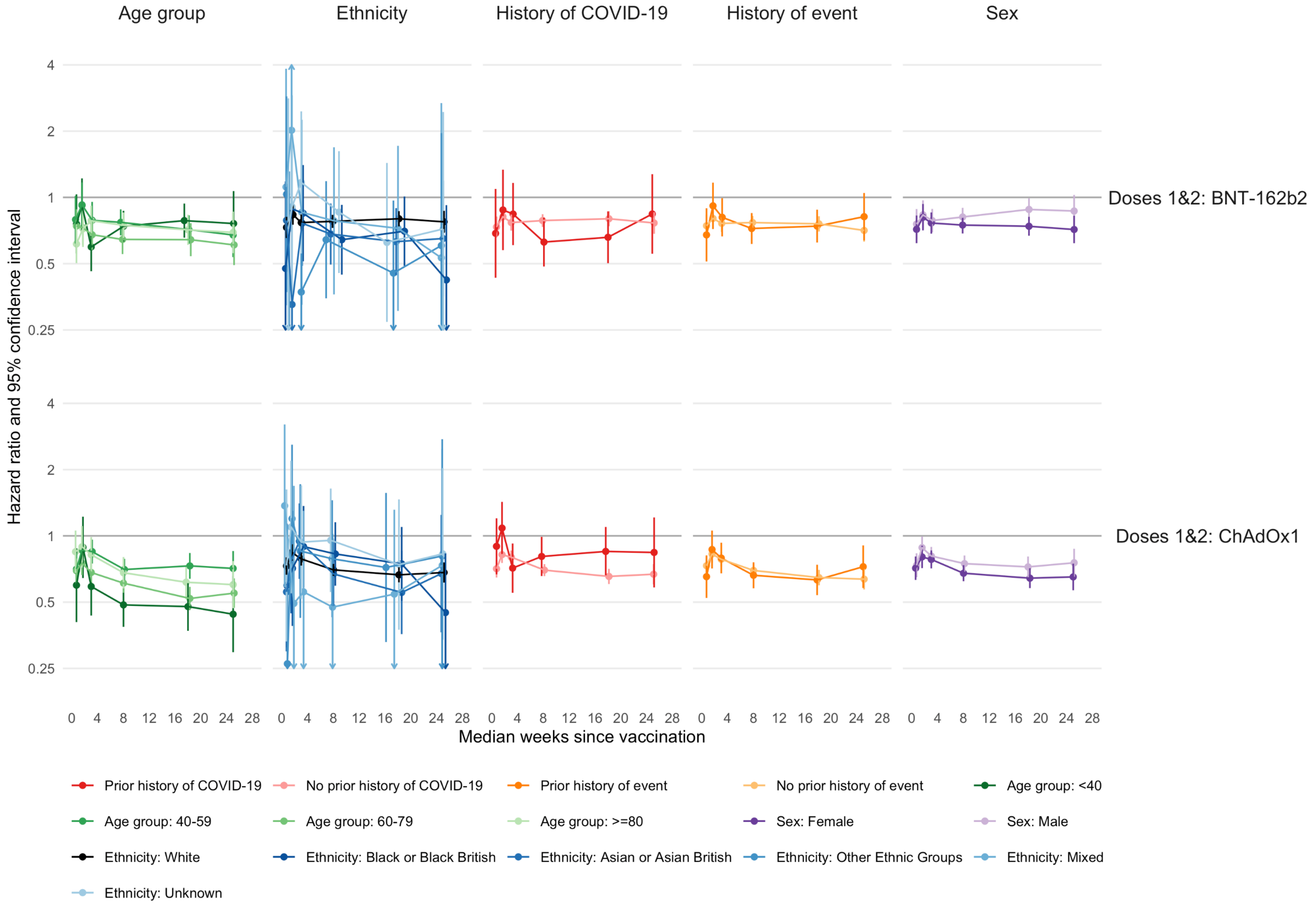

Supplementary Figure 6: Subgroup-specific maximally-adjusted hazard ratios for a composite arterial event by time since booster vaccination[9]

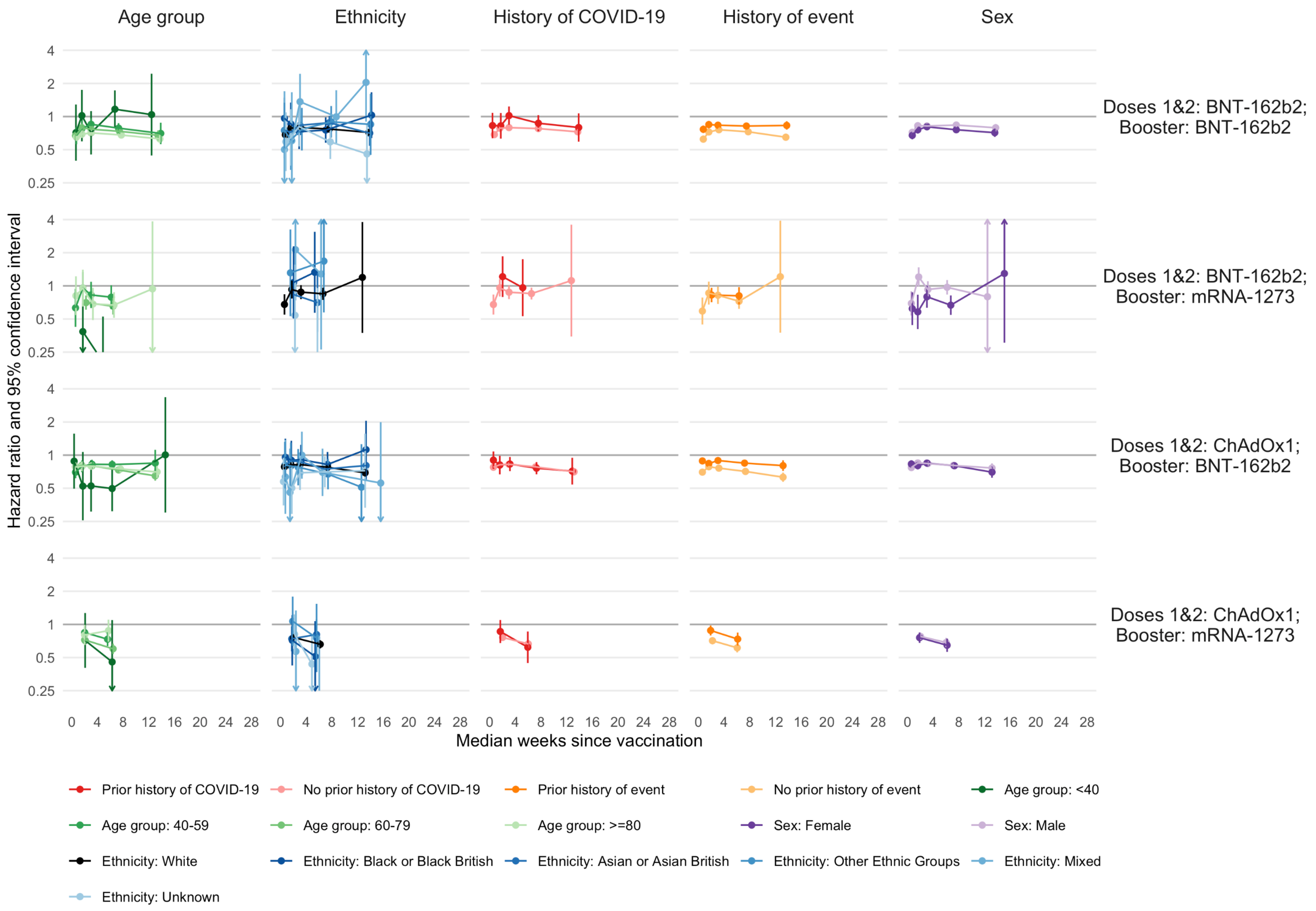

9 The aHR for weeks 12-24  for Age group: 40-59, Doses 1&2: BNT-162b2/mRNA-1273; Booster: mRNA-1273 could not be estimated due to small counts.

Supplementary Figure 7: Subgroup-specific maximally-adjusted hazard ratios for a composite venous event by time since booster vaccination

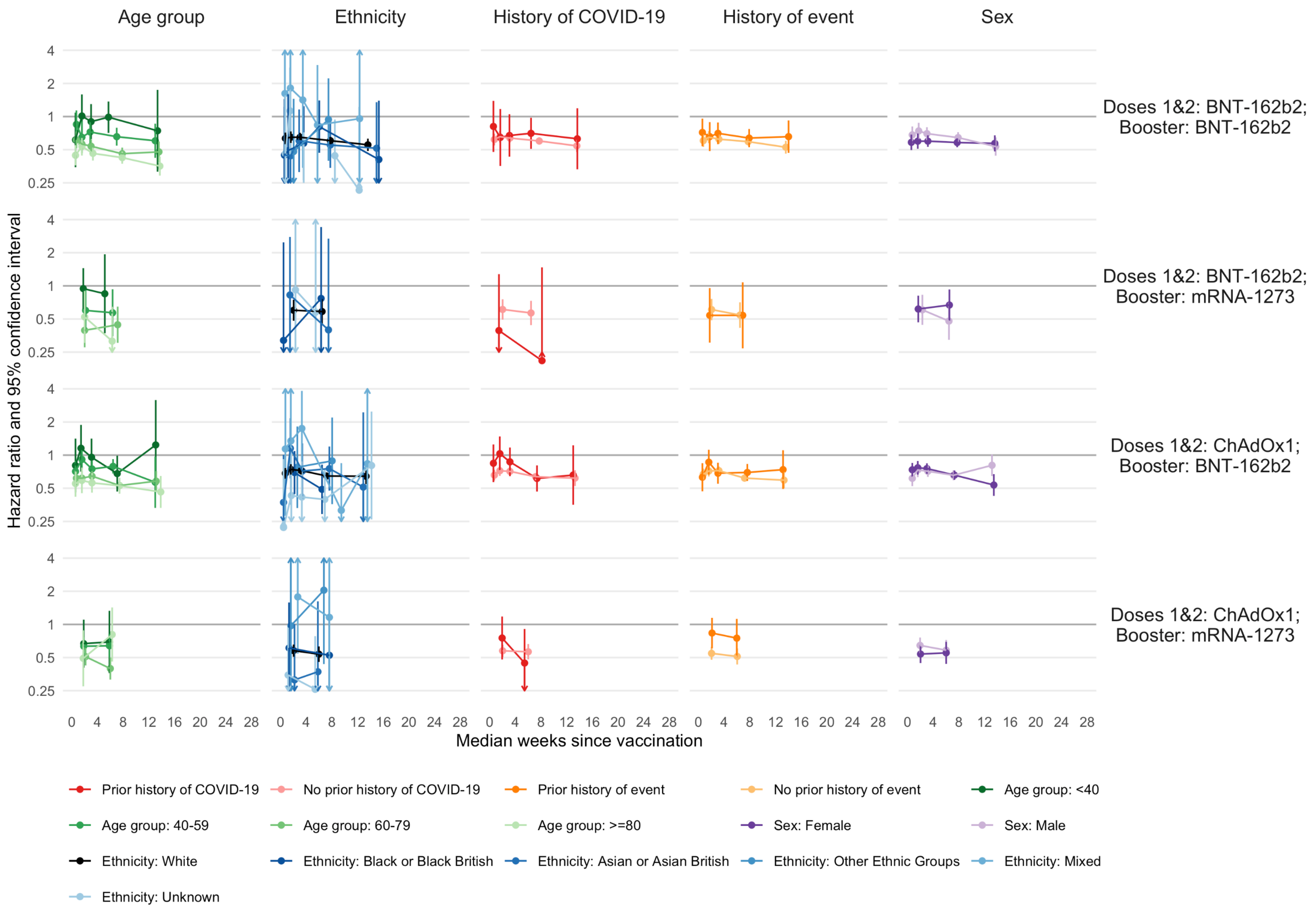

Supplementary Figure 8: Associations of COVID-19 vaccination brand and dose with myocarditis and pericarditis until the CDC's Announcement on 17 May 2021.

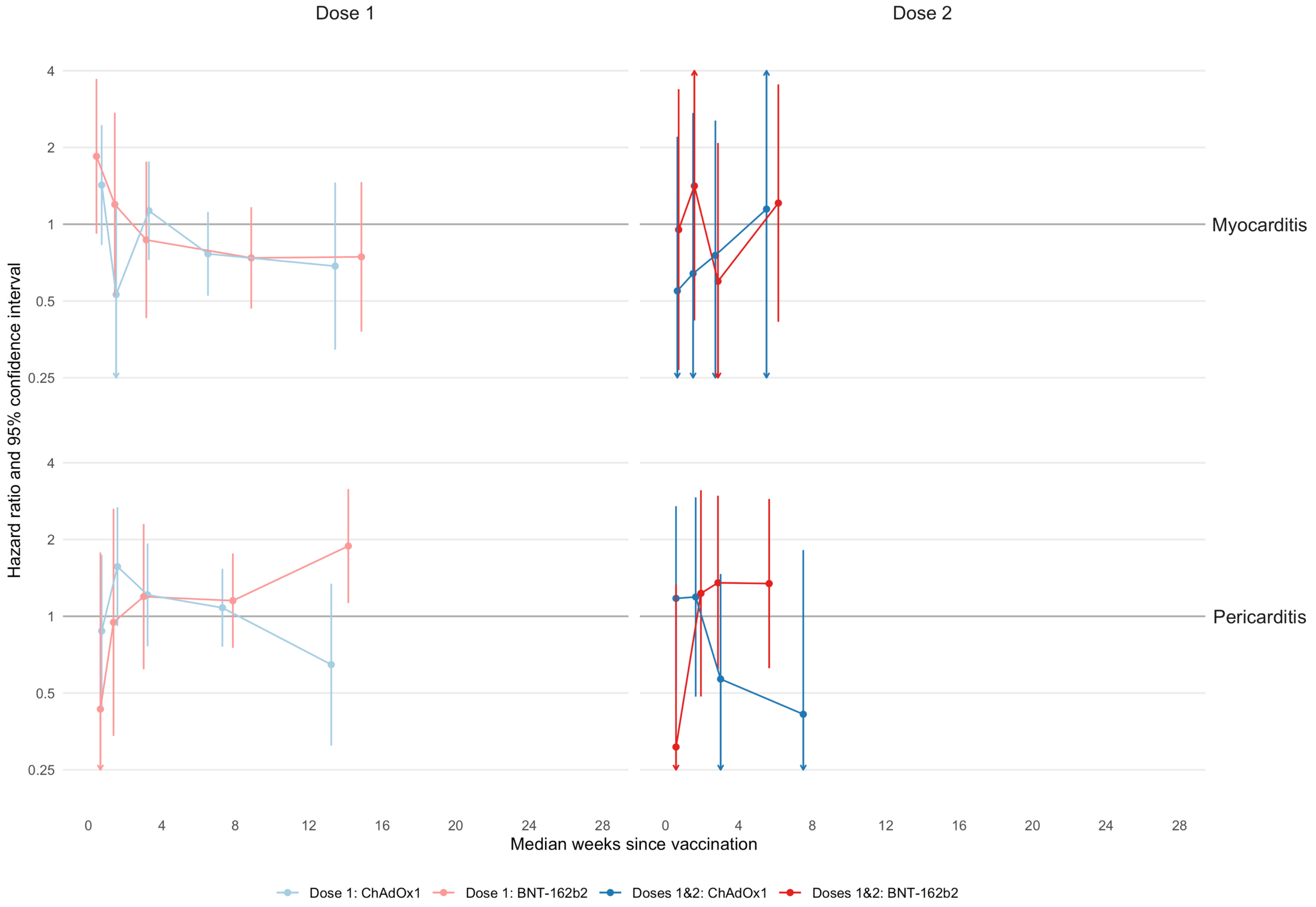
